# Projecting vaccine demand and impact for emerging zoonotic pathogens

**DOI:** 10.1101/2021.11.09.21266135

**Authors:** A Lerch, QA ten Bosch, M L’Azou Jackson, AA Bettis, M Bernuzzi, GAV Murphy, QM Tran, JH Huber, AS Siraj, GM Bron, M Elliott, CS Hartlage, K Koh, K Strimbu, M Walters, TA Perkins, SM Moore

**Affiliations:** Department of Biological Sciences and Eck Institute for Global Health, University of Notre Dame, Notre Dame, Indiana, USA; Quantitative Veterinary Epidemiology, Wageningen University and Research, Wageningen, Netherlands; Coalition for Epidemic Preparedness Innovations (CEPI), London, UK; Coalition for Epidemic Preparedness Innovations (CEPI), Oslo, Norway; Bill & Melinda Gates Foundation, Seattle, Washington, USA

**Author notes:** Corresponding authors: T Alex Perkins, Sean M Moore.

## Abstract

Despite large outbreaks in humans seeming improbable for a number of zoonotic pathogens, several pose a concern due to their epidemiological characteristics and evolutionary potential. To enable effective responses to these pathogens in the event that they undergo future emergence, the Coalition for Epidemic Preparedness Innovations is advancing the development of vaccines for several pathogens prioritized by the World Health Organization. A major challenge in this pursuit is anticipating demand for a vaccine stockpile to support outbreak response. We developed a modeling framework for outbreak response for emerging zoonoses under three reactive vaccination strategies. Annual vaccine regimen requirements for a population-wide strategy ranged from >670,000 (95% prediction interval: 0-3,630,000) regimens for Lassa virus to 1,190,000 (95% PrI: 0-8,480,000) regimens for Rift Valley fever virus, while the regimens required for ring vaccination or targeting healthcare workers (HCWs) were several orders of magnitude lower (between 1/25 and 1/700) than those required by a population-wide strategy. For each pathogen and vaccination strategy, reactive vaccination typically prevented fewer than 10% of cases, because of their presently low R0 values. Targeting HCWs had a higher per-regimen impact than population-wide vaccination. Our framework provides a flexible methodology for estimating vaccine stockpile needs and the geographic distribution of demand under a range of outbreak response scenarios.

## Introduction

Less than two years ago, SARS-CoV-2 was an unknown virus circulating in a zoonotic reservoir (Andersen et al. 2020). In the time since, it has caused a pandemic resulting in more than 4.6 million deaths (WHO 2020). Theoretical work (Antia et al. 2003) predicts that frequent small-scale outbreaks in humans may provide opportunities for selection of more transmissible variants that facilitate emergence from the original reservoir. Indeed, virological studies indicate that a sequence of mutations acquired in this manner may offer a plausible explanation for the emergence of SARS-CoV in 2003 (Sheahan et al. 2008). More frequent spillover and more human-to-human transmission ensuing from those spillovers are expected to increase the probability that adaptations such as these arise and facilitate more widespread emergence (Morse et al. 2012). Because of this evolutionary potential, even zoonotic pathogens with limited human-to-human transmission—as defined by a basic reproduction number, *R*0, below 1—are viewed as a concern. The status quo of investing in the development of diagnostics, therapeutics, and vaccines only in reaction to emerging disease threats has made the world dangerously vulnerable to pandemics (Røttingen et al. 2017; Excler et al. 2021).

To preempt future public health emergencies arising from emerging zoonotic diseases, the World Health Organization (WHO) developed a research and development blueprint for action to prevent epidemics (WHO 2016). This R&D Blueprint prioritizes and regularly updates a list of pathogens for development of diagnostics, therapeutics, and vaccines (https://www.who.int/activities/prioritizing-diseases-for-research-and-development-in-emergency-contexts). The Coalition for Epidemic Preparedness Innovations (CEPI) was launched in 2017 to accelerate the development of vaccines against emerging infectious diseases and to enable equitable access to these vaccines for people during outbreaks (Gouglas et al. 2019; Bernasconi et al. 2020; Huneycutt et al. 2020). The first call for proposals from CEPI was on developing vaccines for Lassa virus (LASV), MERS coronavirus (MERS-CoV), and Nipah virus (NiV). Soon after, it added Rift Valley fever virus (RVFV) and chikungunya virus (CHIKV) to its portfolio. As of early 2021, CEPI was supporting development of a total of 19 different vaccine candidates for these five diseases, in addition to other efforts related to Ebola, COVID-19, and “disease X” (CEPI 2018).

In anticipation of vaccine candidates for these diseases progressing through safety and efficacy trials and towards implementation, there is a need to understand future potential vaccine demand (Røttingen et al. 2017). Even though these vaccines are not yet available for public health use, understanding demand at an early stage is important to inform fundraising and planning efforts in support of the manufacturing and distribution infrastructure that will be required for their implementation (Excler et al. 2021). Following the development of a new vaccine, manufacturing capacities are typically the first limiting factor for vaccine supply, which raises allocation and prioritization decisions to protect people at higher risk of infection and clinical disease (Medlock and Galvani 2009; Bubar et al. 2021). Appropriate planning of vaccine stockpiles to support vaccine demand is important to minimize the extent to which difficult decisions about vaccine prioritization must be made once a vaccine becomes available for use. At the same time, overestimating vaccine stockpile needs could result in doses expiring and resources that could have gone to other needs being wasted.

To improve capabilities to plan vaccine stockpiles for emerging zoonotic pathogens, we developed a modeling framework to quantify the vaccine stockpile size needed to meet demand for outbreak response and applied it to LASV, MERS-CoV, NiV, and RVFV (Figure 1). Each of these pathogens is zoonotic, with the majority of human cases believed to result from spillover transmission from non-human hosts accompanied by self-limiting, human-to-human transmission (Linthicum, Britch, and Anyamba 2016; Cauchemez et al. 2016; Siddle et al. 2018; Nikolay, Salje, Hossain, Khan, Sazzad, Rahman, Daszak, Ströher, Pulliam, Kilpatrick, Nichol, et al. 2019). Our model is driven by geographically and seasonally realistic patterns of spillover for each pathogen, with each spillover event having the potential to spark an outbreak that we simulated stochastically with a branching process model. Outbreak response with reactive vaccination was triggered in our model whenever a threshold number of cases was exceeded within a certain space-time window. We quantified the number of vaccine regimens required (where the number of regimens equals the number of individuals vaccinated) under three different approaches to reactive vaccination: 1) population-wide within the same geographic area as the outbreak, 2) targeted on healthcare workers (HCWs) within that area, or 3) targeted on a ring of contacts around each index case. Using vaccines modeled after target product profiles for each pathogen (WHO 2017c, 2017b, 2017a, 2019), we also quantified the impact of reactive vaccination under a range of scenarios about deployment timing, coverage, per-exposure protection (PEP) from vaccination, and several epidemiological parameters.

**Figure 1.**
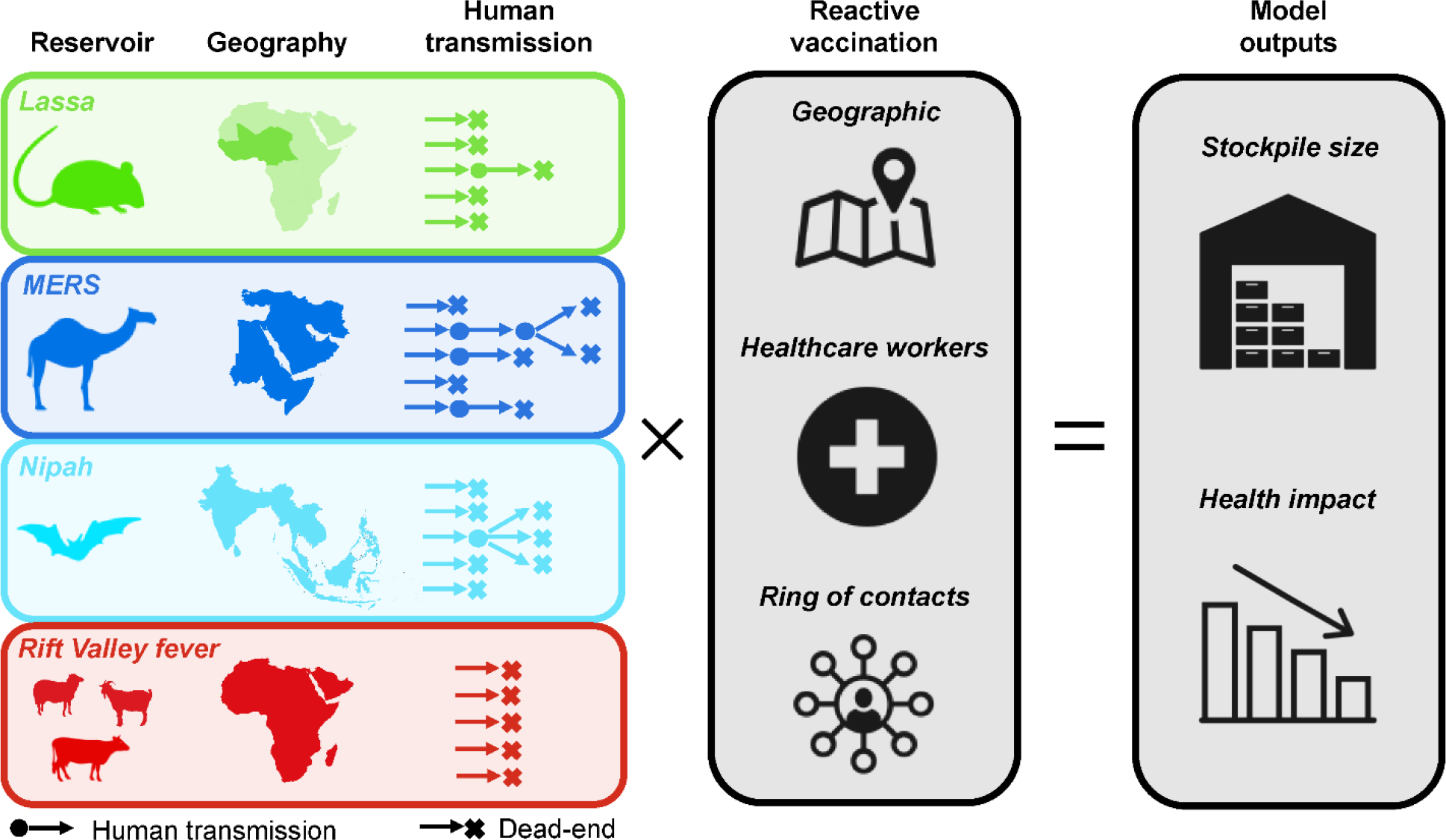
Overview of this study. We considered four emerging zoonoses prioritized by the WHO R&D Blueprint and CEPI. For each, we modeled spillover, human-to-human transmission, and reactive vaccination. We quantified the vaccine stockpile necessary to meet demands of reactive vaccination under three scenarios: vaccinating an entire population within the same geographic area as a detected outbreak, vaccinating healthcare workers within that geographic area, or vaccinating contacts associated with each spillover case. **Lassa fever** is caused by LASV, a virus that circulates in rodents in West Africa and has resulted in thousands of cases and deaths in recent years (Mylne et al. 2015; Roberts 2018). **Nipah** is caused by NiV, a virus that circulates in fruit bats that can be found throughout tropical and subtropical Asia (Yob et al. 2001; Luby et al. 2009), but documented spillover to humans has been mainly limited to India, Bangladesh, and Malaysia (Pulliam et al. 2012; Girish Kumar et al. 2019; Nikolay, Salje, Hossain, Khan, Sazzad, Rahman, Daszak, Ströher, Pulliam, Kilpatrick, Nichol, et al. 2019). **MERS** is caused by MERS-CoV, a coronavirus that probably originated in bats (Anthony et al. 2017), and is known to circulate among domestic camel populations in the Middle East and parts of eastern and northern Africa, resulting in spillover from camels to humans (Müller et al. 2015; Dudas et al. 2018; Hui et al. 2018). Human-to-human transmission has been reported in nosocomial settings for three of these pathogens (Fisher-Hoch et al. 1995; Park et al. 2016; Nikolay, Salje, Hossain, Khan, Sazzad, Rahman, Daszak, Ströher, Pulliam, Kilpatrick, Nichol, et al. 2019), although only MERS was reported in large hospital outbreaks (Assiri, McGeer, et al. 2013; Park et al. 2016). The evidence for community transmission of these viruses is more limited (Siddle et al. 2018; Hui et al. 2018; Nikolay, Salje, Hossain, Khan, Sazzad, Rahman, Daszak, Ströher, Pulliam, Kilpatrick, Nichol, et al. 2019). **Rift Valley fever** is caused by RVFV, a mosquito-transmitted virus infecting ruminant livestock species in Africa, the Arabian Peninsula, and the Indian Ocean islands (Pepin et al. 2010; Bron et al. 2021; Gerken et al. 2021). RVF outbreaks have been associated with heavy rainfall in eastern and southern Africa (Anyamba et al. 2009, 2010), but transmission can also occur outside of these epizootic events (Linthicum, Britch, and Anyamba 2016). Humans can be infected via direct contact with infected animals or via mosquito bite, but are believed to be dead-end hosts (Al-Hamdan et al. 2015).

## Methods

### Epidemiological data

For each of the pathogens, we collated epidemiological data through the end of 2020 from multiple sources, including WHO outbreak reports (e.g., (CSR n.d.)), ProMED reports (https://promedmail.org), country-level reports (https://www.moh.gov.sa, https://ncdc.gov.ng), and a literature search. A detailed overview of the source of epidemiological data for each pathogen can be found in the Supplementary Table S1.

**Table 1.** Overview of parameter estimates. Incubation period and infectious period are defined in units of days, and parameters for seasonality refer to week of the year. Numbers in parentheses for R0 represent the 95% confidence intervals.

| Parameter | LASV | MERS-CoV | NiV | RVFV |
| --- | --- | --- | --- | --- |
| Seasonality |  |  |  |  |
| - Peak (wk) | 31.1 | 23.3 | 27.3 | 23.2 |
| - SD (wk) | 6.2 | 13.6 | 6.4 | 12.7 |
| Incubation period |  |  |  |  |
| - Mean (d) | 12.05 | 5.56 | 9.87 | 2.88 |
| - SD (d) | 3.62 | 0.77 | 0.84 | 1.95 |
| Infectious period |  |  |  |  |
| - Mean (d) | 11.31 | 13.5 | 6.49 | 7<sup>1</sup> |
| - SD (d) | 8.29 | 2.61 | 0.26 | - |
| $R_0$ | | | | |
| - Mean | 0.063 (0.05, 0.08) | 0.58 (0.31, 0.99) | 0.325 (0.21, 0.52) | 0 (0.01) |
| - Dispersion | - | 1.42 | 0.048 | - |

<sup>1</sup> Fixed value used for sensitivity analysis only.

### Spillover simulation

Given extensive spatial heterogeneity of incidence, we collated epidemiological data at the first administrative level (adm1) in each country—e.g., province or state—within the study region for each pathogen. The primary epidemiological data used to inform spillover rates was the annual incidence of reported cases of each pathogen at the adm1 level (Table S1). Where possible, case data was categorized into cases of documented or suspected human-to-human transmission, documented or suspected spillover cases, and cases of unknown origin. The geographic coverage of our analysis for each pathogen was determined by the geographic distribution of spillover cases in the literature. All countries with at least one documented spillover case were included in our analysis. We excluded countries with imported cases but no spillover from a zoonotic source (e.g., South Korea for MERS-CoV).

Spillover rates were estimated using a generalized linear mixed model (GLMM) with a zero-inflated negative binomial distribution to capture overdispersion in the annual distribution of spillover cases within an adm1. Spillover cases were defined as documented spillover cases, suspected spillover cases, or cases of unknown origin; thereby excluding any cases of documented or suspected human-to-human transmission. Year, country, and adm1 were treated as random effects, with the adm1 variable nested within the country variable. Year was also included as a random effect for the zero-inflated portion of the model. Model fitting was conducted using the glmmTMB package in R (Brooks et al. 2017). This default model did not converge for NiV; therefore, for NiV we used the GLMM model without the random effect by year in the zero-inflated portion of the model to enable convergence. Then, for each pathogen, we simulated annual spillover cases for each year and adm1 by taking draws (1,000 replicates) from a zero-inflated negative binomial distribution using the estimated parameters from the appropriate GLMM fit. We randomly sampled 1,000 of these simulated spillovers from the last five years as inputs to the outbreak simulation model so that the simulated spillovers would reflect recent spillover rates.

To account for the seasonality of spillover, we fitted a beta distribution to the timing of spillover cases within a year (daily for MERS, weekly for Lassa fever, monthly for Nipah and RVF) and simulated the timing of each spillover case as a random draw from that distribution (Table 1). To account for spatial clustering of cases below the adm1 level, we associated each simulated case with a catchment area. We did so according to probabilities proportional to catchment area population. Catchment areas were defined by second administrative level (adm2) or hospitals aggregated within 10 km for first administrative (adm1) areas that did not have an adm2 level. These catchment areas, therefore, represent areas where individuals would be expected to seek care and have their diagnosis reported, and the aggregation of hospitals within a 10km area assumes that individuals who seek treatment for the relatively severe symptoms of these diseases do so at larger hospitals. Hospital location data for sub-Saharan Africa used in the analysis of LASV was obtained from (Maina et al. 2019), and hospital location data outside of sub-Saharan Africa was obtained from https://www.healthsites.io (Saameli et al. 2018). The primary set of findings we reported are based on a set of 1,570 catchment areas for LASV, 767 for MERS-CoV, 5,076 for NiV and 2,126 for RVFV, which differ because of the different geography of each pathogen. We examined the sensitivity of our results to the definition of a catchment area by rerunning the analyses with either adm1 catchment areas or all hospitals within an adm1 as distinct catchment areas. The results of these analyses are presented in the Supplement (SI Text).

### Outbreak simulation

To simulate incidence attributable to human-to-human transmission, we considered each spillover case as a potential index case for an outbreak. A schematic overview of both the spillover and outbreak simulation models, including outbreak response, is provided in Figure 2. Human-to-human transmission was simulated stochastically using a branching process model. For each primary case, a certain number of secondary cases was drawn either from a Poisson distribution (for Lassa fever and RVF) with λ = R0, or from a negative binomial distribution (for MERS and Nipah) with μ = R0 and a dispersion parameter, *k*. A Poisson distribution was used for Lassa fever and RVF, because both have an estimated R0<0.1 and no available estimate of overdispersion. We used a negative binomial distribution for MERS and Nipah, because secondary cases for these diseases are known to be overdispersed, with a majority of human-to-human transmission arising from a small minority of primary cases (Cauchemez et al. 2016; Nikolay, Salje, Hossain, Khan, Sazzad, Rahman, Daszak, Ströher, Pulliam, Kilpatrick, and Others 2019).

**Figure 2.**
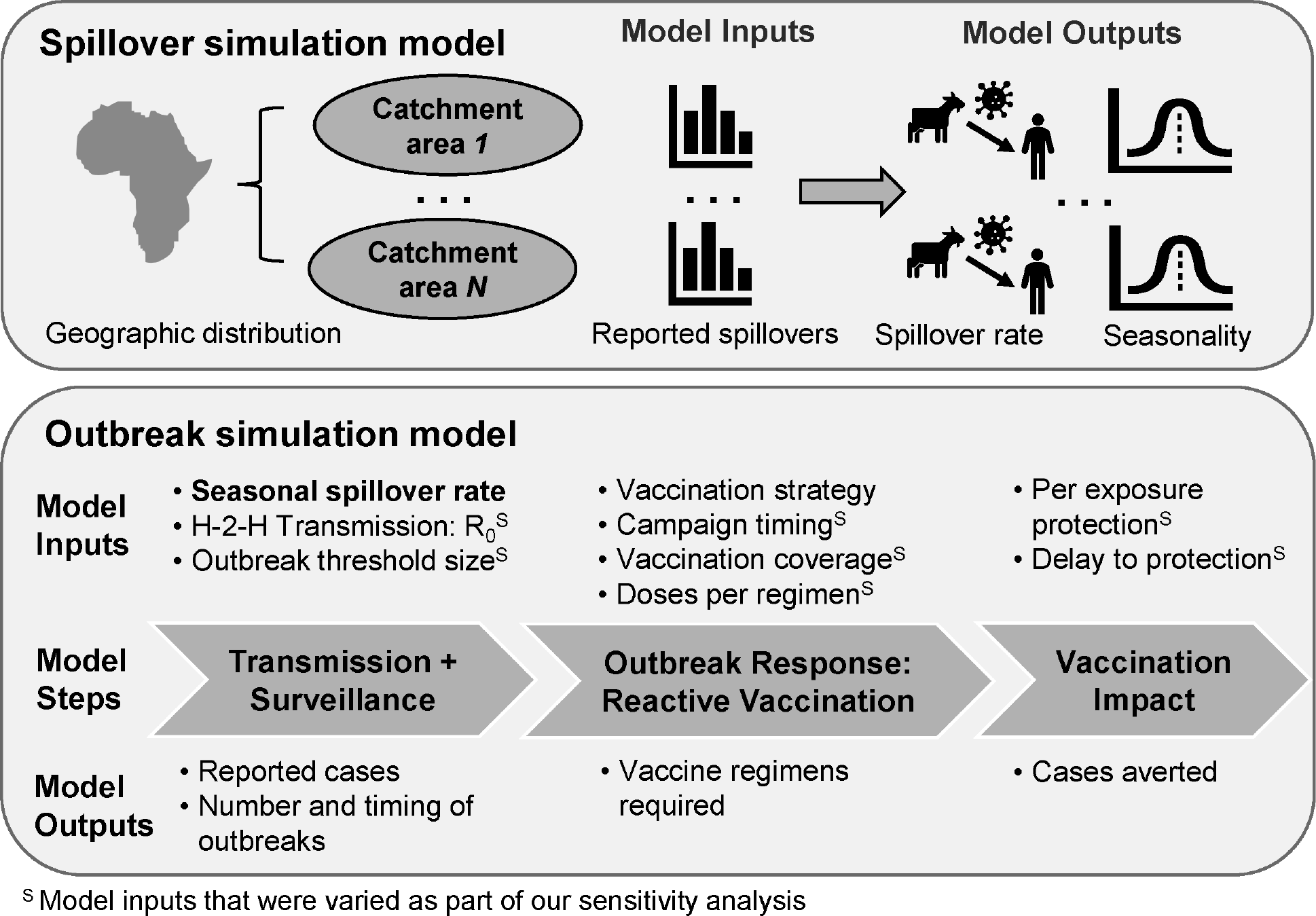
Schematic of the spillover simulation and outbreak simulation models. The spillover simulation model estimates the magnitude and timing (seasonality) of the spillover rate for each catchment area from the historical distribution of reported spillovers in the catchment area. These estimated spillover rates are input into our outbreak model for each catchment area (as identified by the bolded model input), which used a branching process model to simulate human-to-human transmission. An outbreak response was triggered within a catchment area if the number of reported cases exceeded a predetermined number within a 28d time window (outbreak threshold size). Outbreak model inputs with a superscript S were varied as part of our sensitivity analysis.

We estimated R0 and variability therein differently for each pathogen. For LASV, we estimated an R0 for nosocomial transmission by fitting a simple branching process model to observed outbreak sizes from (Lo Iacono et al. 2015) using the optimize function in R and assuming a Poisson offspring distribution (Farrington, Kanaan, and Gay 2003). The resulting estimate of R0 for LASV was 0.063 (95% confidence interval [CI]: 0.05 - 0.08) (Table 1). For MERS-CoV, we compiled estimates of R0 from multiple studies analyzing data from MERS outbreaks (Cauchemez et al. 2014; Breban, Riou, and Fontanet 2013; Poletto et al. 2014; Chowell et al. 2014; Cauchemez et al. 2016; Kucharski and Althaus 2015) and described variability in those estimates with a gamma distribution, which resulted in a median R0 of 0.583 (90% CI: 0.31 - 0.99). The dispersion parameter estimate, *k*=0.26, for MERS-CoV was obtained from (Kucharski and Althaus 2015). For NiV, we estimated R0 and its variability from detailed epidemiological investigations of Nipah outbreaks in Bangladesh that estimated person-to-person chains of NiV transmission (Nikolay 2019). Using data from these studies on the number of secondary infections per primary infection and the size of each transmission cluster, we obtained maximum-likelihood estimates of R0 (0.33, 95% CI: 0.21 - 0.52) and *k* (0.048, 95% CI: 0.031 - 0.074), which were consistent with a branching process with a negative binomial offspring distribution. For RVFV, we assumed R0=0, and considered R0=0.01 for sensitivity analysis only, as no human-to-human transmission has been definitively documented to date (Al-Hamdan et al. 2015).

The timing of incubation and infectious periods were then simulated subsequently based on gamma distributions of those periods that we estimated by fitting a model to reconcile variability in previously published estimates (Table 1). As no human-to-human transmission is known for RVFV, we assumed for the sensitivity analysis a fixed duration for the infectious period of 7 days that is consistent with the duration of detectable viremia after onset of symptoms (Bird et al. 2009). For all pathogens, the infection date of secondary cases was simulated as a draw from a uniform distribution over the infectious period of the primary case. Each secondary case was assigned to the same catchment area as the associated index case. A detailed overview of the source for each parameter of each pathogen can be found in the Supplementary Table S1.

### Vaccine campaign simulation

Three different reactive vaccination strategies were evaluated: 1) vaccinating a portion of the general population in a given catchment area; 2) specifically targeting the HCWs in that catchment area; or 3) adopting a ring vaccination strategy where the local population surrounding each index case are targeted for vaccination. These strategies were chosen as they represent three of the most frequently deployed outbreak response strategies. For each strategy, baseline vaccination campaign parameter values (and parameter ranges for the sensitivity analysis) were based on vaccine target product profiles for each pathogen (WHO 2017c, 2017b, 2017a, 2019), or chosen in consultation with CEPI and subject-matter experts for each pathogen (Table 2).

**Table 2.**
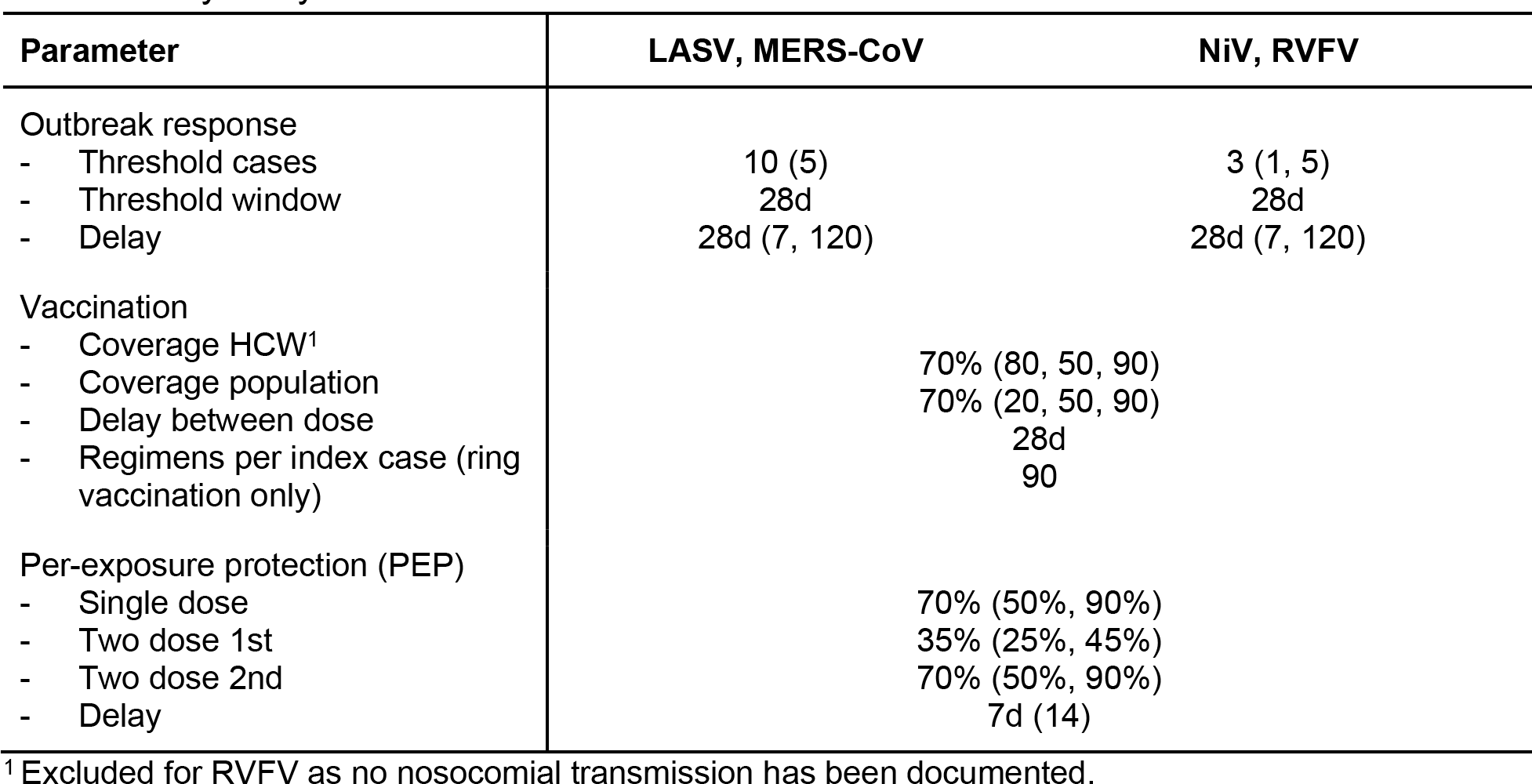
Overview of simulation scenarios. Parameter values for the baseline reactive vaccination scenario for each pathogen. Outbreak response threshold cases and threshold window refer to the number of cases that need to occur within a certain time window to trigger an outbreak response. Parameter values in parentheses are alternative values used as a part of the sensitivity analysis.

To estimate the impact of vaccination, we simulated each outbreak response relative to a counterfactual simulation in which there was no outbreak response. Vaccination impact was defined as the number of cases averted via vaccination and calculated by taking the difference between the number of cases in the vaccination and no-vaccination scenarios. In our baseline scenario, an outbreak response within a single catchment area was triggered once ten cases for Lassa fever and MERS or three cases for Nipah and RVF were detected within a four-week window (Table 2). These outbreak response thresholds were chosen through discussion with CEPI and pathogen experts, and do not necessarily match the different outbreak definitions currently used by WHO or individual countries. The vaccination start date was calculated by adding a delay to the outbreak response date. To simplify vaccine uptake in our model, we assumed that each target population was immunized on a single day. Multi-day vaccination campaigns would likely reduce the impact of outbreak response relative to our estimates, but this impact would be less severe than a comparable delay in protection following vaccination because at least a portion of the population would be protected at the beginning of the campaign. Therefore, our analysis of the sensitivity of vaccination impact to a delay in protection following vaccination could be considered an upper bound on the sensitivity to extending the vaccine administration period for a given round of vaccination. In the case of a 2-dose vaccine, an additional delay of 28 days was assumed between administration of the first and second doses.

For the general population vaccination strategy, HCWs were treated as part of the general population and were vaccinated with the same probability as the general population. For the HCW vaccination strategy, non-HCWs were not vaccinated, except for a hybrid strategy tested as part of our sensitivity analysis, where 20% of the general population was vaccinated versus 80% of HCWs (Table 2). For the ring vaccination strategy, we calculated the number of index cases that would arise after the reactive vaccination campaign had started and assumed that 90 vaccine regimens would be needed to vaccinate a ring of individuals around each index case based on estimates from ring vaccination campaigns during recent Ebola and cholera outbreaks (Ali et al. 2016; Henao-Restrepo et al. 2017). For the ring vaccination strategy, we only estimated the number of vaccine regimens that would be required and did not attempt to estimate the impact of vaccination on cases averted, because our model was designed to simulate a single vaccine campaign and not the periodic deployment as required by a ring vaccination strategy.

Once a vaccination campaign was completed and the delay between vaccination and protective immunity had elapsed, vaccination in the general population removed spillover cases with a probability equal to vaccination coverage in the general population multiplied by per-exposure protection (PEP). Vaccination of the general population also removed patient-to-HCW nosocomial cases with probability equal to vaccination coverage in HCWs multiplied by PEP. Vaccination of HCWs had no impact on spillover cases, but it removed nosocomial cases with probability equal to vaccination coverage in HCWs multiplied by PEP. PEP depended on whether a sufficient amount of time since vaccination had elapsed and, in the event of a two-dose vaccine, whether an individual had received one dose or two doses at the time of exposure (Table 2). Cases downstream in a transmission chain from a case averted by vaccination were also averted.

### Vaccine demand calculation

To quantify the number of regimens required to meet the demands of a given outbreak response strategy, we estimated the number of healthcare workers and overall population associated with each catchment area where an outbreak occurred. The overall population per catchment area was estimated based on WorldPop data from 2015 (Tatem 2017). For healthcare workers, we took the national-level numbers of healthcare workers and distributed them proportional to the population associated with each catchment area (Ref (WHO 2021)).

### Graphical user interface

A generalized implementation of the model is provided as a graphical user interface (GUI) at http://eidvaccinedemand.crc.nd.edu. In the generalized implementation, a few adjustments were made to allow for more flexible application of the model and to make computing time more acceptable for an interactive web tool. First, annual spillovers are drawn from a negative binomial distribution and then distributed across the catchment areas with a multinomial distribution proportional to the probability that spillovers occur in these catchment areas.

Second, the population in the catchment areas were defined by a negative binomial distribution so that specific geographies did not need to be reproduced. The default parameters for the GUI of each pathogen were obtained by fitting the corresponding distribution function to the estimated spillover and population data from this study. The source code for the GUI is provided at https://github.com/lerch-a/CEPI_VaccineCampaignGUI.

## Results

### Spillover cases and human-to-human transmission

The median annual number of spillover cases was 6 (95% prediction interval: 0-190) for Nipah, 114 (95% PrI: 48-266) for MERS, 185 (95% PrI: 8-13,134) for RVF, and 417 (95% PrI: 142-1,837) for Lassa fever (Figure 3A). Simulated variability in the annual number of spillover cases matched the cumulative distribution of observed spillover cases for each pathogen (SI Figures S1B-S4B). Spillover rates for each pathogen varied both seasonally (SI Figures S1A-S4A) and geographically (Figure 4A). Spillover cases of Lassa fever were concentrated in Sierra Leone, Liberia, and Nigeria, although a few spillover cases occurred in other western African countries. Spillover of RVF to humans was widespread in South Africa, Madagascar, eastern Africa and the Arabian Peninsula, with frequent spillover cases occurring in several western and northern Africa countries as well. The majority of MERS spillover cases occurred in Saudi Arabia, and the majority of Nipah spillover cases occurred in Bangladesh, with additional spillover events in India and Malaysia.

**Figure 3.**
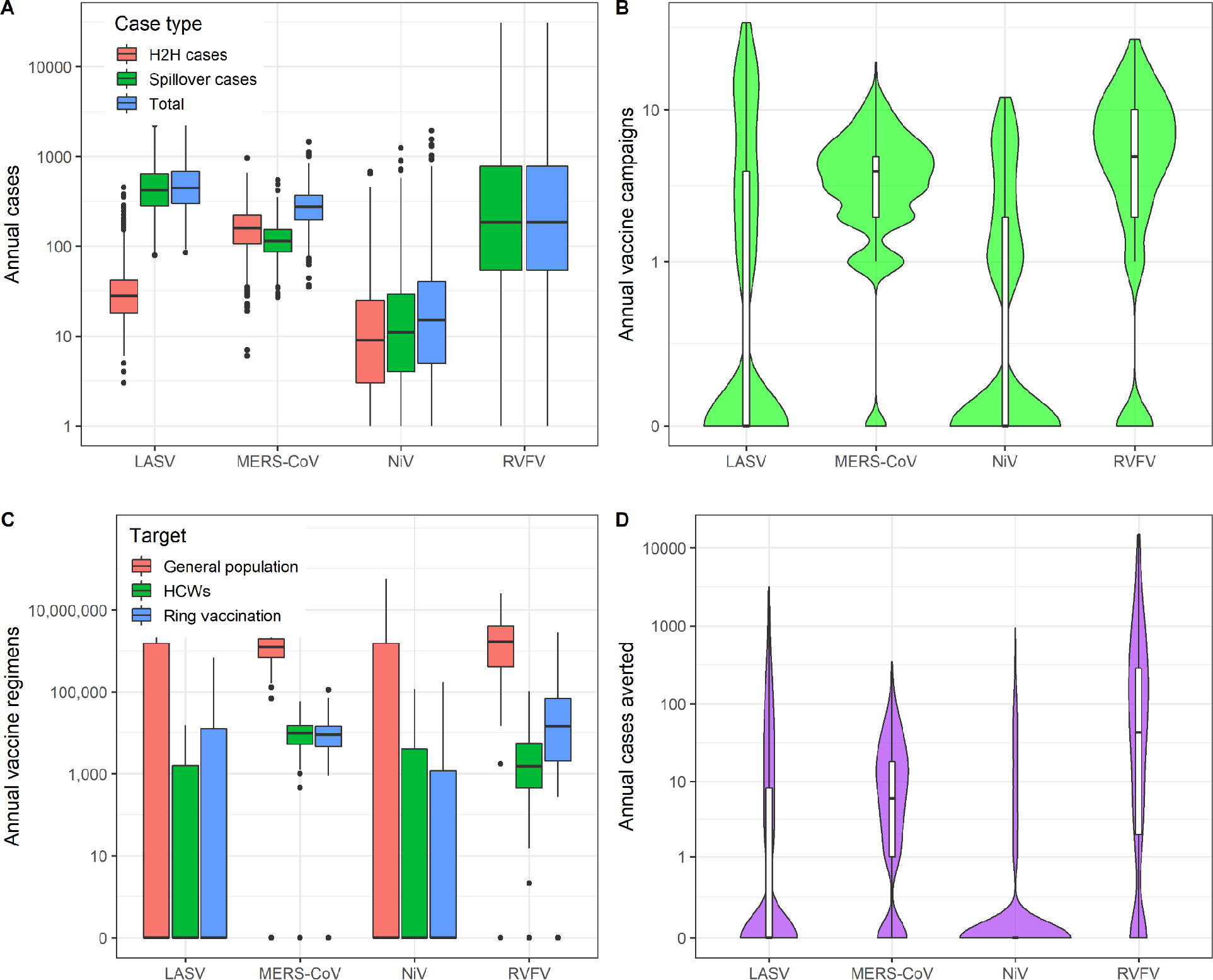
Simulated annual cases and reactive vaccination impacts. (A) Annual number of spillover, human-to-human (H2H), and total cases for each pathogen across the entire study region (in the absence of vaccination). (B) Violin plot (including box plot representing the median, IQR, and 95% CI) of the annual number of vaccine campaigns triggered due to the outbreak threshold being exceeded across 1,000 simulations for each pathogen. (C) Number of vaccine regimens required per year for reactive vaccination under our baseline scenario under three alternative assumptions about the target of vaccination campaigns. (D) Violin plot (including box plot representing the median, IQR, and 95% CI) of annual number of cases averted by reactive vaccination campaigns across 1,000 simulations for each pathogen. All y-axes are log10 scaled.

**Figure 4.**
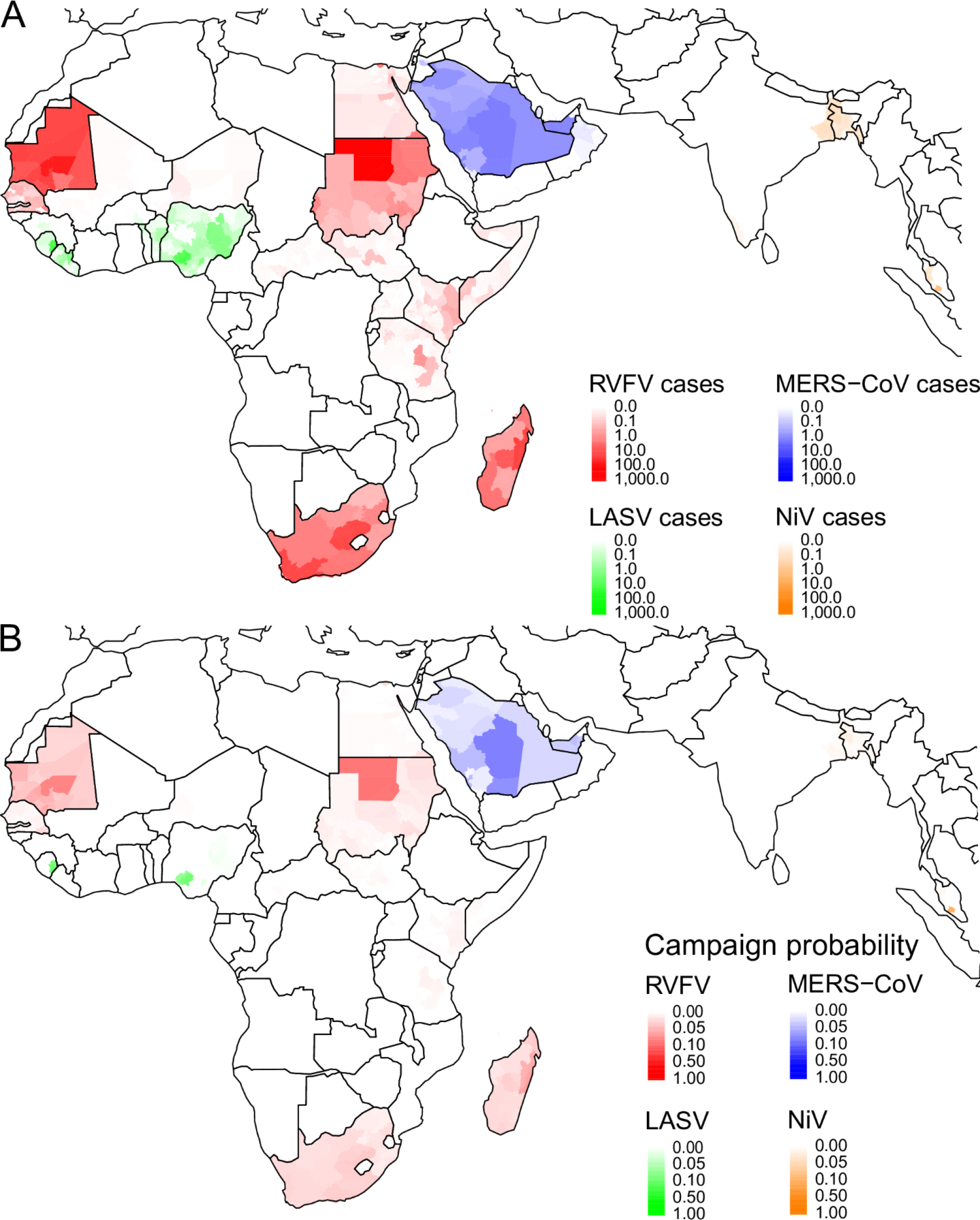
Geographic distribution of predicted spillover cases and reactive vaccination campaigns. (A) Geographic distribution at the 2^nd^ administrative level (adm2) of the expected annual number of spillover cases for each pathogen. (B) The annual probability that a campaign will be triggered in each adm2 catchment area based on 1,000 simulations.

The number of cases arising from human-to-human transmission depended on both the spillover rate and R0 (Figure 3A). Under our default parameter assumptions, there was no human-to-human RVFV transmission, but in the absence of vaccination the median annual number of human-to-human cases following spillover was 2 (95% PrI: 0-82) for Nipah, 29 (95% PrI: 11-143) for Lassa fever, and 161 (95% PrI: 46-407) for MERS (see Figure 5 for an example of the transmission chains for one catchment area).

**Figure 5.**
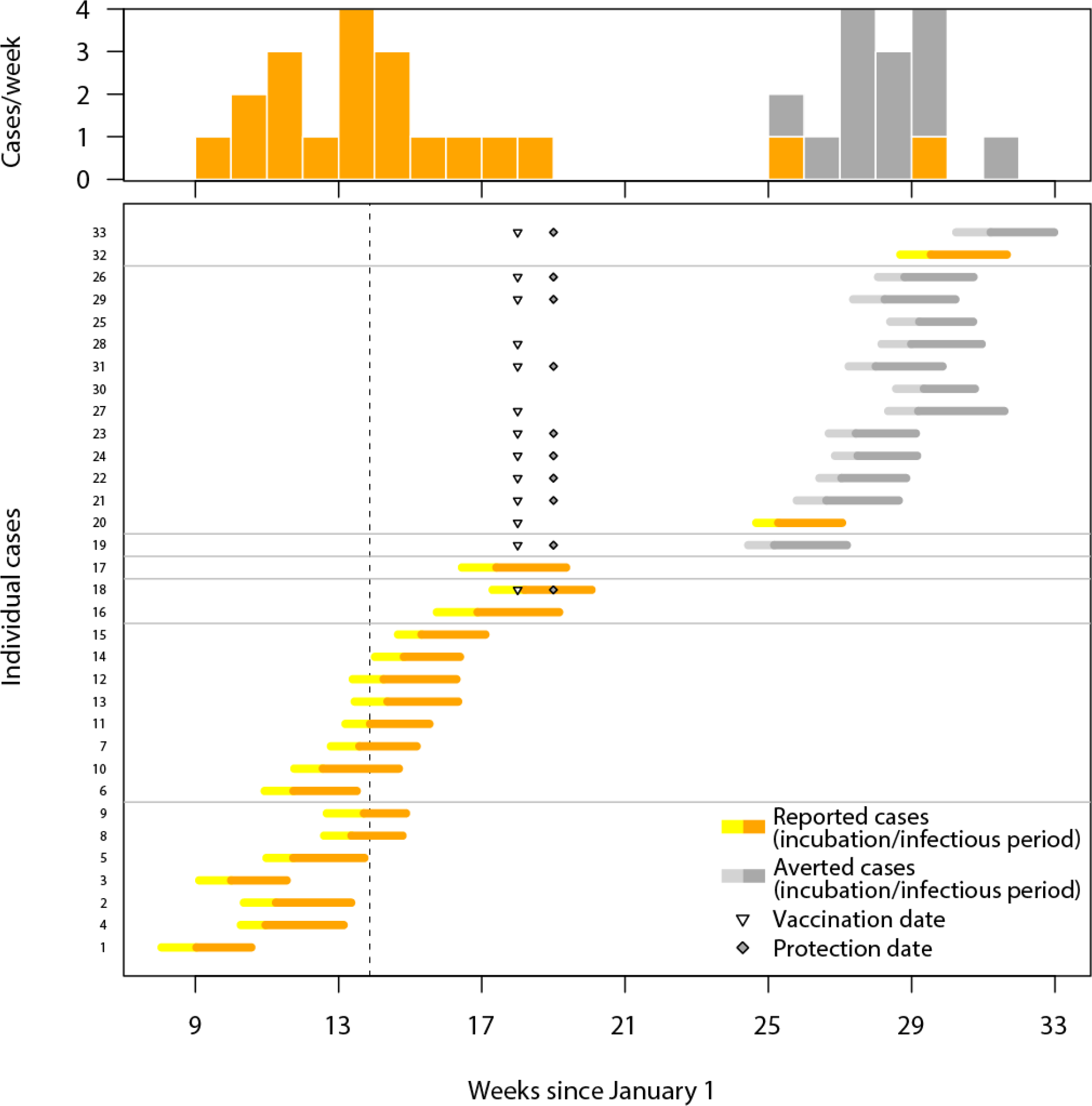
Timing of spillover and nosocomial cases in a single realization of one catchment area from the MERS-CoV outbreak model. (Bottom) Individual cases are visualized as thick horizontal lines, with observed cases in yellow/orange and averted cases in gray (yellow and light gray indicate incubation time, orange and dark gray indicate infectious time). Unrelated transmission trees are separated by thin horizontal gray lines. The dashed vertical line indicates the date the outbreak threshold was reached. Triangles indicate vaccination date and diamonds indicate protection date. (Top) Number of observed (orange) and averted (gray) cases per week.

### Estimates of vaccine demand

In our analysis, a median of 0 (95% PrI: 0-8) Nipah reactive vaccination campaigns were triggered annually, compared to 4 (95% PrI: 0-11) MERS campaigns, 5 (95% PrI: 0-20) RVF campaigns, and 0 (95% PrI: 0-20) Lassa fever campaigns (Figure 3B). The locations of reactive vaccination campaigns broadly followed the geographic distribution of spillovers for each pathogen, although Lassa fever spillovers in Guinea, Benin, Togo, and western Nigeria were rarely reported frequently enough to trigger a response in our simulations (Figure 4B). The number of reactive vaccination campaigns that were triggered, and the timing of those campaigns, was strongly influenced by the seasonal pattern of pathogen spillover (SI Figures S1-S4).

For all four pathogens, there was a wide range in the number of vaccine regimens required in a typical year due to the dependence of vaccine demand on the spatiotemporal clustering of spillover cases required to trigger an outbreak response. The largest annual vaccine demand was for RVFV, with a median of 1,191,741 (95% PrI: 0-8,480,275) vaccine regimens required to target the general population under our baseline outbreak response scenario (Figure 3C). The median number of vaccine regimens for MERS-CoV was 870,045 (95% PrI: 0-2,843,407). The median number of vaccine regimens needed for NiV and LASV was zero, implying that an outbreak response was triggered less than 50% of the time. However, the mean number of vaccine regimens was 673,167 (95% PrI: 0-3,629,052) for LASV and 1,450,177 (95% PrI: 0-12,240,814) for NiV (Figure 3C). The number of vaccine regimens required to conduct a ring vaccination strategy or to cover healthcare workers as a part of an outbreak response was typically several orders of magnitude (between 1/25 and 1/700) lower than the number required to cover the general population (Figure 3C). The median number of MERS-CoV vaccine regimens required to cover healthcare workers was 6,786 (95% PrI: 0-22,086). A median of 1,540 (95% PrI: 0-62,320) vaccine regimens were needed among healthcare/veterinary workers for RVFV outbreak response, 0 (mean: 1,144; 95% PrI: 0-6,485) were required for LASV, and 0 (mean: 2,330; 95% PrI: 0-15,833) for NiV. The median number of vaccine regimens required for ring vaccination was 4,860 (95% PrI: 0-21,429) for MERS-CoV, 12,150 (95% PrI: 0-1,175,758) for RVFV, 0 (mean: 13,774; 95% PrI: 0-108,056) for LASV, and 0 (mean: 2,605; 95% PrI: 0-21,641) for NiV.

### Impact of outbreak response

The estimated impact of reactive vaccination as an outbreak response tool was generally low for all four pathogens. Vaccinating 70% of the general population in response to an outbreak with a single-dose vaccine prevented an annual median of 43 (95% PrI: 0-5,853) RVF cases, 6 (95% PrI: 0-83) MERS cases, 0 (95% PrI: 0-90) Nipah cases, and 0 (95% PrI: 0-357) cases of Lassa fever (Figure 3D). These vaccine impacts correspond to 0.69 (95% PrI: 0-2.92) cases averted per 100,000 vaccine regimens administered for MERS, 3.61 (95% PrI: 0-69.02) for RVF, 0 (95% PrI: 0-9.84) for Lassa fever, and 0 (95% PrI: 0-0.74) for Nipah. Vaccinating only healthcare workers typically had a smaller total impact than vaccinating the general population at the same coverage level, because there was no protection against spillover in the general population, but a larger per-regimen impact due to the lower number of regimens required. Vaccinating 70% of HCWs prevented an annual median of 4 (95% PrI: 0-77) MERS cases, corresponding to 58.9 (95% PrI: 0-348.6) cases averted per 100,000 vaccine regimens in HCWs. Vaccinating HCWs averted 0 (95% PrI: 0-46) Lassa fever cases and 0 (95% PrI: 0-48) Nipah cases, corresponding to 0 (95% PrI: 0-710.4) and (95% PrI: 0-303.5) cases averted per 100,000 HCW vaccine regimens respectively (we did not explore vaccinating HCWs against RVFV due to the lack of any documented nosocomial transmission).

### Sensitivity analysis

The number of total cases increased with higher R0 values for each pathogen, with the largest sensitivity observed for MERS-CoV, because its higher value of R0 was close to one (Figure S16). There was also a large increase in the number of vaccine regimens required to vaccinate either the general population or HCWs for MERS-CoV at the higher R0 value, but the impact of R0 on the required number of vaccine regimens was minimal for the other pathogens (Figures S17-S18). As a result, there were minimal differences in the impact of vaccination under higher or lower R0 values for LASV, NiV, or RVFV (Figures S19-S22). Vaccination averted both a greater magnitude and a higher fraction of MERS cases as R0 increased (Figures S19-S20). In addition, the number of MERS cases averted per vaccine regimen administered to the general population or to HCWs also increased as R0 increased (Figures S21-S22).

Lowering the outbreak threshold (from 10 to 5 cases within a 28 day window for MERS-CoV and LASV, and from 3 to 1 cases for NiV and RVFV) increased both the number of vaccine regimens needed for outbreak response and the number of cases averted. With the lower outbreak threshold, the projected demand for MERS-CoV vaccine regimens was 2,351,059 (95% PrI: 492,028-5,872,847), a 170% increase, while the median number of cases averted was 19 (95% PrI: 0-162), a 217% increase compared to the baseline. The required number of vaccine regimens for RVFV increased to 4,793,351 (95% PrI: 659,297-14,157,197), a 302% increase, while the median number of RVF cases averted was 66 (95% PrI: 0-6,066), a 53% increase. The median number of vaccine regimens for LASV increased from 0 to 756,273 (95% PrI: 0-6,644,995), and the median number of Lassa fever cases averted increased from 0 to 15 (95% PrI: 0-534). The median number of vaccine regimens for NiV increased from 0 to 3,501,587 (95% PrI: 0-54,814,275), but the median number of cases averted remained 0 (95% PrI: 0-119). When the outbreak threshold was increased to 5 cases for RVF, the required number of vaccine regimens decreased by 50% to 594,894 (95% PrI: 0-7,493,183). The number of RVF cases averted via vaccination decreased to 26 (95% PrI: 0-5,735), which was 41% fewer cases averted compared with an outbreak threshold of 3 cases.

Decreasing the time delay between the outbreak threshold being reached and the start of the vaccination campaign tended to increase the number of cases averted, while increasing the delay reduced the number of cases averted (Figure 6). For MERS-CoV, reducing the time delay from 28 to 7 days increased the median number of cases averted from 6 (95% PrI: 0-83) to 14 (95% PrI: 0-112), while increasing the delay to 120 days reduced the number of cases averted to 0 (95% PrI: 0-38).

**Figure 6.**
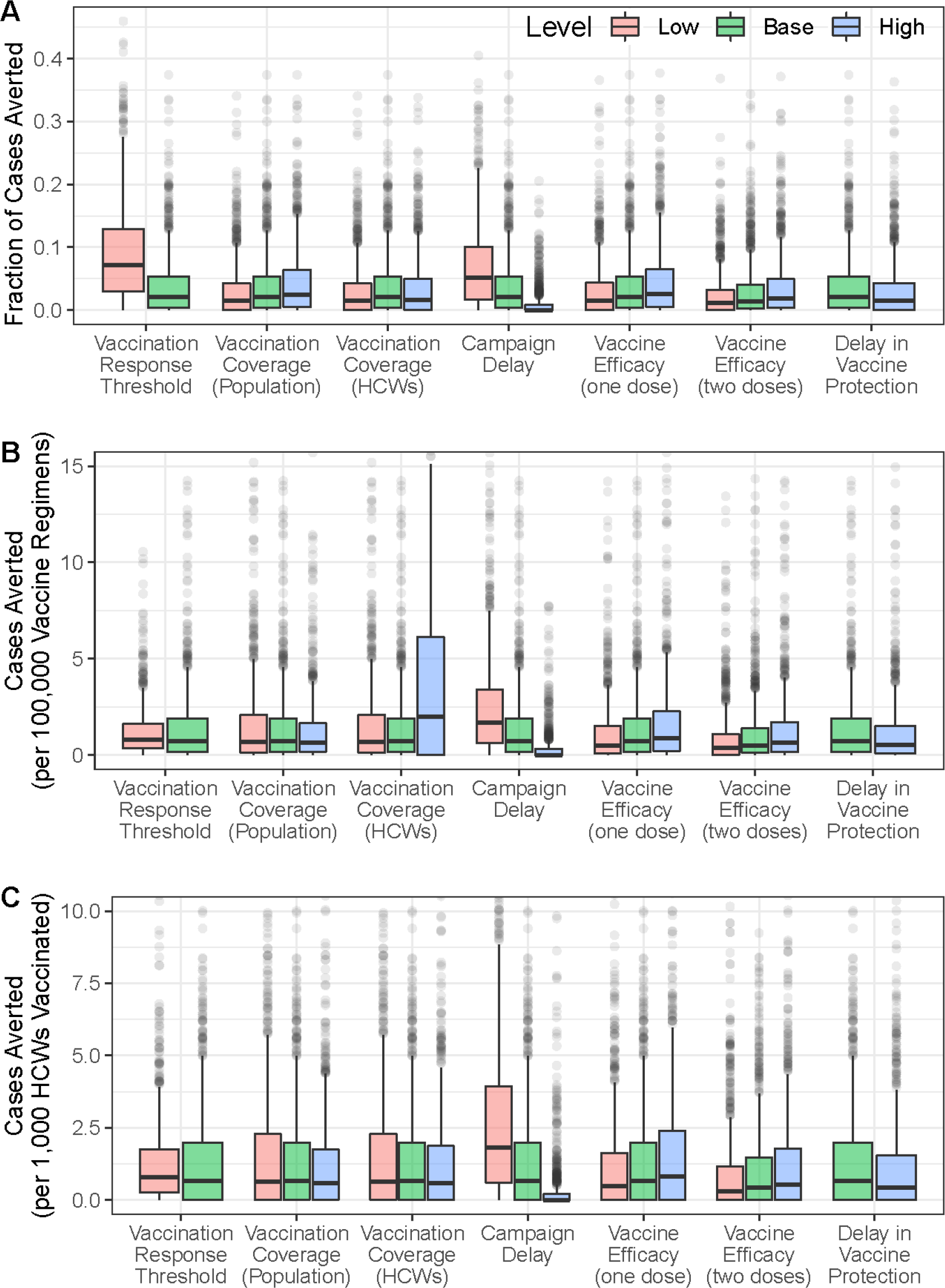
Vaccine impact sensitivity analysis for MERS-CoV. Sensitivity of vaccination impact for MERS-CoV to variation in different campaign parameters expressed as (A) fraction of cases averted, (B) cases averted per 100,000 vaccinated in the general population, and (C) cases averted per 1,000 health care workers (HCWs) vaccinated.

Increasing or decreasing the percentage of the population that was targeted during reactive vaccination campaigns also led to corresponding increases or decreases in the number of cases averted (Figure 6). For example, if only 50%, rather than 70%, of the population was vaccinated for MERS-CoV, the median number of cases averted declined from 6 (95% PrI: 0-to 4 (95% PrI: 0-72). In contrast, if vaccination coverage was increased to 90%, then 7 (95% PrI: 0-93) MERS cases were averted. The number of MERS cases averted per 100,000 vaccine regimens administered decreased from 0.69 (95% PrI: 0-2.92) at 70% coverage, to 0.64 (95% PrI: 0-3.55) at 50% coverage, and 0.63 (95% PrI: 0-2.54) at 90% coverage. The sensitivity of the impact of outbreak response to other campaign parameters considered in our model, including per-exposure protection, time to protection following vaccination, vaccination coverage levels in HCWs, and one-dose vs. two-dose vaccines are provided in Figure 6. The sensitivity analyses for the other pathogens (NiV, LASV, and RVFV) and for different catchment levels are provided in the Supplement (SI Text). In general, the number of cases averted were highest when the spatial scale for vaccine response (catchment area) was the first administrative level, but the per-regimen vaccination impact was higher for the smaller catchment areas (second administrative level or hospital-based catchment areas), because fewer vaccine regimens were required per campaign in those areas (Figures S33-S34).

## Discussion

### Model performance

Our spillover simulation model estimates closely matched the average annual reported number of spillover cases for each pathogen, as well as the observed interannual variability in the number of spillover cases that have occurred in the past few decades. The simulation results also captured the geographic distribution and seasonality of spillover cases for each pathogen. The magnitude, spatial distribution, and timing of spillover rates are the main determinants of how frequently an outbreak response threshold will be triggered and therefore the size of the vaccine stockpile needed for outbreak response. Although these patterns could shift to some degree in the future, our model represents what we know about them presently. In addition to influencing stockpile size, these three factors (the magnitude, spatial distribution, and timing of spillover rates) are also relevant for logistical considerations such as the geographic location(s) of the stockpile and the necessary stockpile replenishment rate (Yen et al. 2015).

### Stockpile estimates

The estimated number of vaccine regimens needed to reach vaccination coverage targets in the general population varied considerably across the four pathogens examined. For both LASV and NiV, the median was zero, indicating that reactive vaccination campaigns would not be triggered more than 50% of the time. In contrast, the median numbers of vaccine regimens needed for MERS-CoV and RVFV were 870,000 and 1,190,000, respectively. However, the 95% prediction intervals for all four pathogens were wide due to spatial and temporal heterogeneity in spillover rates and overdispersion in outbreak sizes resulting from human-to-human transmission. For all four pathogens, the vaccine regimens needed to target HCWs were several orders of magnitude lower than needed to target the general population.

These results indicate that the size of the vaccine stockpile needed to meet annual reactive vaccination demands will depend on the pathogen’s epidemiology, the vaccine coverage strategy, and the specific demands of a sustainable manufacturing strategy. In addition to the median or mean annual vaccine demand, our estimates also provide an estimate of the inter-annual variability in vaccine demand and the potential magnitude of vaccine demand in low-frequency, but high-demand years. For example, the 75th or 90th percentile of our estimates correspond to the level of demand experienced once every four or ten years, on average. The desired size of a vaccine stockpile will likely depend not only on the average annual vaccine demand, but also on the stockpile capacity needed to adequately handle the unpredictability in the timing, frequency, geography, and magnitude of outbreaks. These questions will depend on sustainable vaccine manufacturing capacity, the geographic distribution of both this manufacturing capacity and the stockpile, and vaccine shelf life. A graphical user interface is available at http://eidvaccinedemand.crc.nd.edu to facilitate interactive exploration of these dependencies.

Our vaccine demand estimates indicate that the biggest determinant of the size of the reactive vaccine stockpile needs was the vaccination strategy: targeting the general population, only HCWs, or ring vaccination. For pathogens that primarily cause nosocomial outbreaks (e.g., LASV), vaccinating HCWs can protect high-risk individuals. In our analysis, this strategy had a larger impact in terms of cases averted per vaccine regimen than vaccinating the general population. The impact of vaccinating HCWs will be highest when spillovers are highly spatially clustered because vaccination campaigns are more likely to be triggered in high-spillover catchment areas, thereby protecting HCWs against nosocomial transmission in areas where vaccination has already occurred earlier in the transmission season but where the spillover risk may remain high. A ring vaccination strategy would also require significantly fewer regimens than a general vaccination strategy. We estimated that the vaccine demand under a ring vaccination strategy would be similar to the demand under a HCW-vaccination strategy for LASV, NiV, and MERS-CoV, and moderately higher than the HCW-vaccination strategy for RVFV. Another strategy to reduce the number of vaccine regimens needed per reactive campaign that we did not consider in our analysis would be to target high-risk sub-populations instead of the whole population of a catchment area. In the case of RVFV, this would be animal workers like butchers, veterinarians, and farmers who are at highest risk of infection (Wilson et al. 1994; Nyakarahuka et al. 2018; Msimang et al. 2019). For MERS-CoV, camel workers have a higher risk of infection than the general population (Dudas et al. 2018). For LASV, rural populations within a catchment area are assumed to have a higher risk than urban populations (but see Chika-Igwenyi et al. 2021, where >50% of patients in one outbreak were urban residents). For NiV, rural populations and people drinking raw date palm sap could be targeted for vaccination (Rahman et al. 2012; Islam et al. 2016).

In addition to providing an estimate of vaccine stockpile size, our modeling approach also provides an estimate of where the stockpile will most frequently need to be deployed. An understanding of the geographical distribution of vaccine demand is critical for sustainable manufacturing and timely response to outbreaks (Grais et al. 2008; Azman and Lessler 2015; Wells et al. 2019). Knowledge of vaccine needs by geographic area is essential so that the stockpile(s) can be strategically positioned for rapid deployment following the triggering of an outbreak response. Vaccine demand in a given area will be a function of the probability of an outbreak response being triggered and the size of the target population. Because we used a sliding time window for the outbreak threshold, the probability of a reactive vaccination campaign being triggered will also depend on the seasonality of spillover. Spillover cases that are highly seasonal will be more likely to trigger a response than spillovers that occur sporadically throughout the year. Highly seasonal spillover rates also increase the importance of rapid deployment of reactive vaccination campaigns, because the shorter duration of the transmission season increases the likelihood that any delays would cause campaigns to occur only after seasonal spillover transmission has declined.

The size of the outbreak-response catchment areas (our baseline catchment area at the 2nd administrative level vs. 1st administrative units or individual hospitals within each 1st administrative unit) also had a large impact on the frequency and timing of outbreak response. First-level administrative catchment areas triggered more outbreak responses and also have larger population sizes, and would therefore require a larger vaccine stockpile. However, this result assumes that the outbreak threshold (number of cases needed to trigger a reactive vaccination campaign) is the same regardless of the size of the catchment area. Adjusting the threshold size based on the geographic extent or population size of the catchment areas would alter the stockpile requirements and could be one approach to aligning expected stockpile demands with manufacturing capacity. The expected number of regimens needed for adm1 catchment areas might also be an overestimate if only certain regions in an adm1 are at risk.

Therefore, another approach that could balance the advantage of expanded adm1 catchment surveillance areas against the larger stockpile requirements would be to monitor spillover cases at the adm1 level, but limit reactive vaccination to the adm2 regions within the adm1 catchment area where spillover cases were observed.

### Vaccination impact

Our results indicate that reactive vaccination strategies for preventing the transmission of zoonotic pathogens with R0<1 tend to have limited impacts. For each of the four pathogens we considered, reactive vaccination of the general population averted fewer than 100 cases per year on average and required more than 10,000 vaccine regimens per case averted. The largest impact (as measured by total cases averted or fraction of cases averted) was achieved for RVFV, which was the only pathogen where >5% of total cases were averted via reactive vaccination under our default assumptions. On a cases-averted per regimen basis, vaccinating HCWs was more effective than vaccinating the general population for each of the pathogens with at least some human-to-human transmission in nosocomial settings (LASV, MERS-CoV, and NiV), suggesting that targeting this group may be a viable strategy for reducing the spread of zoonotic pathogens that are capable of nosocomial transmission.

Under our baseline reactive vaccination scenario, vaccination averted a higher proportion of RVF cases than cases of the other three diseases, even though we assumed that there was no human-to-human RVFV transmission. The higher impact of reactive vaccination for RVFV was the result of two factors. First, our default threshold to trigger an RVFV vaccination campaign was three cases (compared to 10 cases within a 28-day window for LASV or MERS-CoV), which led to more RVFV campaigns being triggered than for the other diseases. Second, RVFV spillovers are highly clustered in space and time, so additional spillover cases were often concentrated in catchment areas where previous spillovers during the transmission season had already triggered a reactive vaccination campaign. Although the lower threshold led to more vaccine regimens being required for RVFV than for the other pathogens, the per regimen impact of reactive vaccination was still highest for RVFV. These results highlight the importance of understanding the underlying epidemiology of zoonotic pathogens when assessing the feasibility of a reactive vaccination strategy. The spatial and temporal heterogeneity in spillover patterns will be a primary factor determining the potential impact of reactive vaccination for pathogens where cases primarily occur via zoonotic spillover rather than human-to-human transmission. With a sensitive case threshold for triggering a vaccination campaign, and a relatively quick response time (28 days), our results indicate that ∼25% of RVF cases could be averted. However, if the response time is slower (120 days), fewer than 5% of RVF cases would be averted via reactive vaccination. This highlights the importance of rapid response and vaccine deployment to the success of reactive campaigns when spillover is seasonal.

After RVFV, the impact of vaccination was modestly higher for the pathogen (MERS-CoV) with the highest R0 (baseline R0=0.58), indicating that rapid deployment of a reactive vaccination campaign can avert a fraction of cases for pathogens capable of at least some sustained human-to-human transmission. However, even for MERS-CoV, fewer than 10% of annual cases were averted by reactive vaccination, even under our most optimistic scenario with a minimal delay. This was partly because a significant fraction of cases were spillover cases in geographic areas where no vaccination campaign was triggered, and partially because reactive vaccination often did not occur rapidly enough to avert a significant proportion of cases resulting from secondary human-to-human transmission. The one scenario where reactive vaccination had a large impact on MERS-CoV transmission was with a higher R0 value of 0.99. In this case, 84.0% (95% PrI: 10.7-97.5%) of MERS cases could be averted under our baseline reactive vaccination scenario, compared to only 2.1% (95% PrI: 0-18.2%) of cases averted with the default R0=0.58. This result highlights the increased potential impactof a reactive vaccination strategy as R0 approaches or exceeds one and self-sustaining human-to-human transmission chains that lead to larger outbreaks become more likely.

### Reactive vs. prophylactic vaccination

Delays between the triggering of the outbreak threshold and vaccine administration limit the impact of reactive vaccination. In most simulated outbreaks, the outbreak died out before the vaccination was administered due to the low R0. In light of this, prophylactic immunization of HCWs or people at high risk could have a larger impact than reactive vaccination. However, a potentially important aspect that was not considered in our study was the impact that reactive vaccination campaigns in one year had for protection in subsequent year(s). Depending on the duration of vaccine-derived immunity, the number of cases averted in subsequent years could be substantial, particularly if the geographic clustering of spillovers is fairly consistent from year to year. For example, in the past few years, some catchment areas in Nigeria have experienced outbreaks of Lassa fever multiple years in a row (Siddle et al. 2018; Roberts 2018). As an extension of our work, the number of averted cases in the years following a reactive vaccination campaign could be estimated based on the spillover rate, the probability of an outbreak, and the durability of vaccine-derived immunity.

### Limitations

We have attempted to estimate vaccine stockpile needs and identify the most important determinants of success for reactive vaccination of zoonotic emerging pathogens by modeling several vaccination strategies and exploring the sensitivity of our results to different aspects of pathogen natural history and vaccine deployment. However, there are some limitations to our approach that could affect these estimates. We briefly mention the main limitations here and include an expanded discussion of these limitations in the SI Text.

First, there is a relatively poor understanding of the epidemiology of most emerging zoonotic pathogens, and data that could be used to try and elucidate the most important aspects of their epidemiology is limited (Grange et al. 2021). In this study, we collated epidemiological data and parameter estimates from a variety of published sources and also consulted pathogen-specific experts, but, inevitably, our approach was limited by current knowledge. Second, because the modeling framework is intended to be applicable for a range of emerging zoonotic pathogens, it cannot incorporate all of the specific epidemiological details that might affect vaccine demand or impact for a particular pathogen. Our focus was on the key aspects of epidemiology and outbreak response that influence sustainable manufacturing needs, vaccine stockpile requirements, and the impact of outbreak response. Third, we only considered reported cases when estimating pathogen spillover rates, because undiagnosed or unreported infections would not trigger an outbreak response, which could bias the geographic distribution of vaccine demand away from areas with limited disease surveillance systems. This decision was made to ensure that our framework could be implemented with existing data only, and therefore could be applied to other pathogens in a straightforward manner.

Fourth, because the extent of community transmission for each of the study pathogens is poorly understood, we assumed that human-to-human transmission was limited to nosocomial settings, which could result in an underestimate of vaccine demand. However, our modeling framework could be used to explicitly represent community transmission dynamics, and for pathogens with R0 << 1, as was largely the case in this study, the limited size of the modeled transmission chains would be similar in either a community or hospital setting since we did not restrict the potential number of contacts per index case. Fifth, we also assumed that all nosocomial transmission was from patients to HCWs or between HCWs, and that there was no patient-to-patient or HCW-to-patient transmission. Therefore, our estimates of the impact of vaccinating HCWs represents an upper-bound on the effectiveness of this strategy, as instances of patient-to-patient transmission would not be prevented via this strategy. Sixth, another simplifying assumption of our model is that cases in one catchment area do not lead to transmission or an outbreak outside of that catchment area. However, our model already implicitly incorporates the possibility of spread between catchment areas, and although our model does not predict spillover cases occurring outside of each pathogen’s currently documented geographic distribution, the reactive vaccination strategies we examined should also be applicable for responding to imported cases and their associated outbreaks. Finally, we did not consider any targeted vaccination strategies beyond ring vaccination or targeting healthcare workers to limit nosocomial outbreaks.

## Conclusion

To inform the development of sustainable vaccine manufacturing processes for emerging pathogens, we developed a modeling framework to estimate the necessary reactive vaccine stockpile size for emerging zoonotic pathogens. Our framework provides a flexible methodology for estimating vaccine stockpile needs for outbreak response, and for exploring the impact of epidemiology and vaccination strategies on outcomes that have important logistical implications for sustainable vaccine manufacturing, such as the geographic distribution of demand or the required stockpile replenishment rate. However, our model showed that the impact of reactive vaccination for the four pathogens that we explored was minimal, preventing fewer than 10% of human cases under most scenarios with their current epidemiology. However, all these pathogens are closely monitored for their outbreak potential, and control measures are needed. Targeting populations at higher risk of infection, such as HCWs, had a higher per-regimen impact than population-wide vaccination in outbreak control situations. Our results highlight the need for a more thorough epidemiological understanding of these, and other, emerging zoonotic pathogens. Improved pathogen surveillance and case detection are also essential for improving the model and our estimates of vaccine demand. Further work exploring additional scenarios, such as the possibility of targeting certain high-risk populations or the potential uses of vaccines for outbreak prevention rather than just outbreak response, is also needed to improve the potential impacts of vaccination.

## Data Availability

All data used in the present study were obtained from public sources and cited in the manuscript. The collated data, along with code to run our model, are provided at https://github.com/lerch-a/CEPI_VaccineCampaignGUI.

## Acknowledgements

We thank Jim Robinson of CEPI and several members of the CEPI Scientific Advisory committee for helping formulate the reactive vaccination strategies examined in this manuscript and for providing feedback on our modeling framework. We thank John Edmunds (LASV), Robert Sumaye (RVFV), Bart Haagmans (MERS-CoV), Emily Gurley (NiV), and Steve Luby (NiV) for serving as pathogen-specific experts and providing assistance with identifying key epidemiological features and data gaps for their respective pathogens. We thank Nico Vandaele for feedback on how epidemiological modeling can help inform the planning and development of sustainable vaccine manufacturing. We thank Shweta Bansal and Romain Garnier for discussions regarding the modeling of emerging infectious disease outbreaks and vaccination.

## Supplementary Information

Table S1. Overview of data references.

**Figure S1. Spillover and reactive vaccination patterns for Lassa fever virus (LASV).**

**Figure S2. Spillover and reactive vaccination patterns for Middle Eastern respiratory virus (MERS-CoV).**

**Figure S3. Spillover and reactive vaccination patterns for Nipah virus (NiV).**

**Figure S4. Spillover and reactive vaccination patterns for Rift Valley fever virus (RVFV). SI Text. Sensitivity analysis.**

**Figures S5-S34 in SI Text.**

## Supplementary Information Table of Contents

**Table S1.**
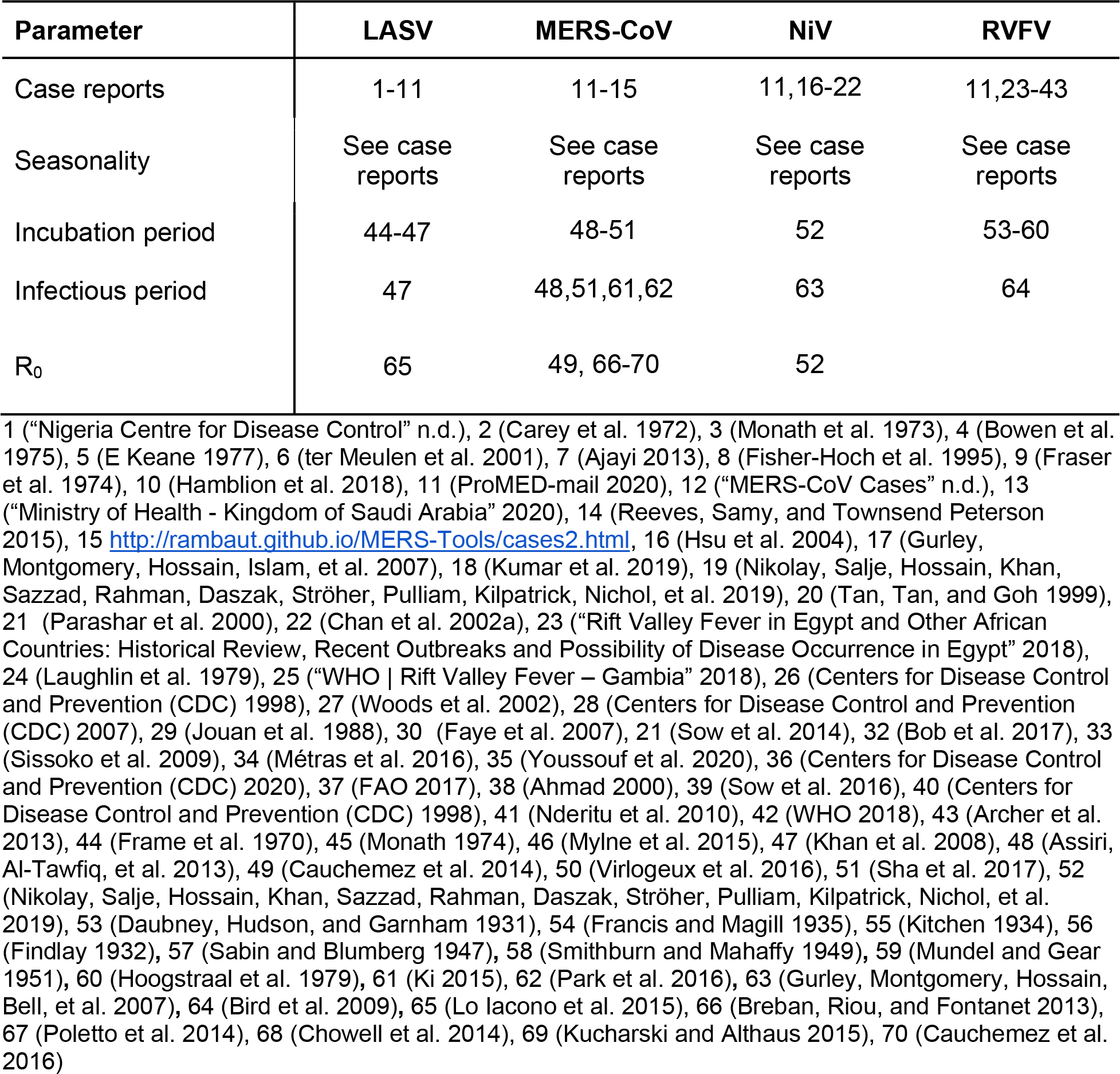
Overview of data references.

| Parameter | LASV | MERS-CoV | NiV | RVFV |
| --- | --- | --- | --- | --- |
| Case reports | 1-11 | 11-15 | 11,16-22 | 11,23-43 |
| Seasonality | See case reports | See case reports | See case reports | See case reports |
| Incubation period | 44-47 | 48-51 | 52 | 53-60 |
| Infectious period | 47 | 48,51,61,62 | 63 | 64 |
| R<sub>0</sub> | 65 | 49, 66-70 | 52 |  |

**Figure S1.**
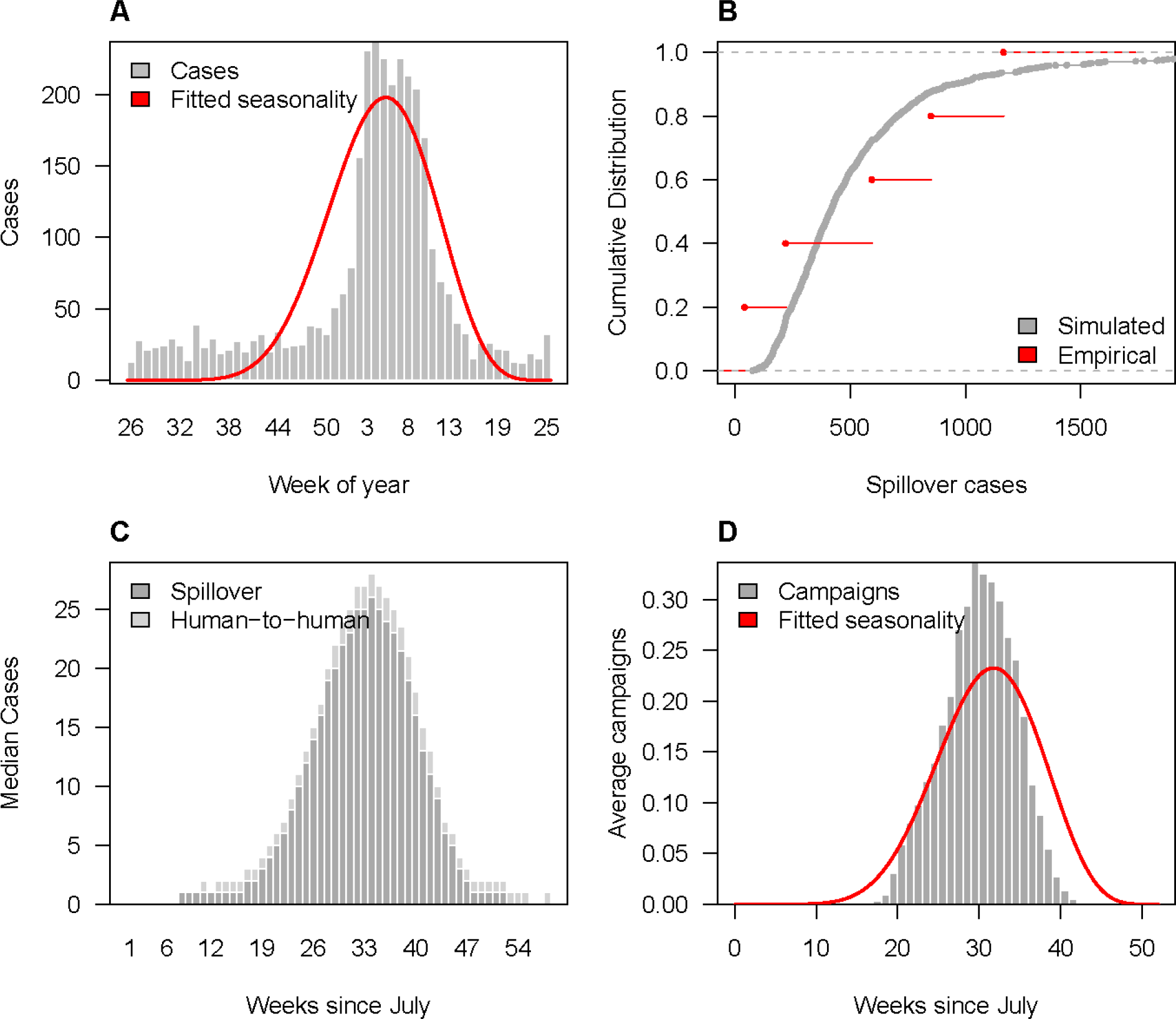
Spillover and reactive vaccination patterns for Lassa fever virus (LASV). (A) Observed weekly Lassa fever spillover cases (grey bars) and estimated seasonal spillover rate (red line). (B) Annual number of spillovers over the past 5 years (red) and cumulative distribution of simulated annual spillovers from 1000 replicates (grey). (C) Median weekly simulated spillover and human-to-human Lassa fever cases. (D) Average weekly number of reactive campaigns triggered via spillover detection compared to the estimated seasonal spillover rate (red line).

**Figure S2.**
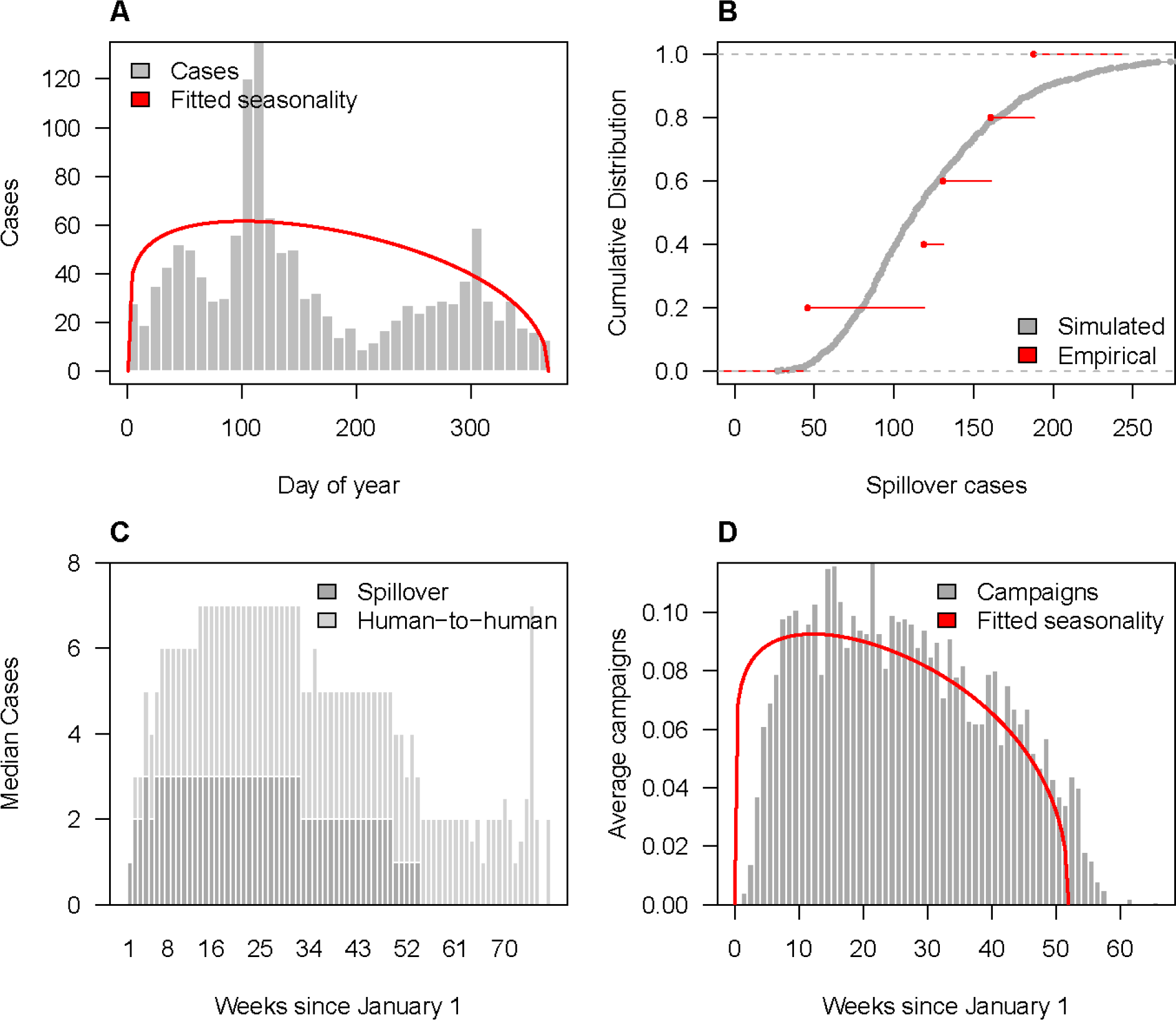
Spillover and reactive vaccination patterns for Middle Eastern respiratory virus (MERS-CoV). (A) Observed weekly MERS spillover cases (grey bars) and estimated seasonal spillover rate (red line). (B) Annual number of spillovers over the past 5 years (red) and cumulative distribution of simulated annual spillovers from 1000 replicates (grey). (C) Median weekly simulated spillover and human-to-human MERS cases. (D) Average weekly number of reactive campaigns triggered via spillover detection compared to the estimated seasonal spillover rate (red line).

**Figure S3.**
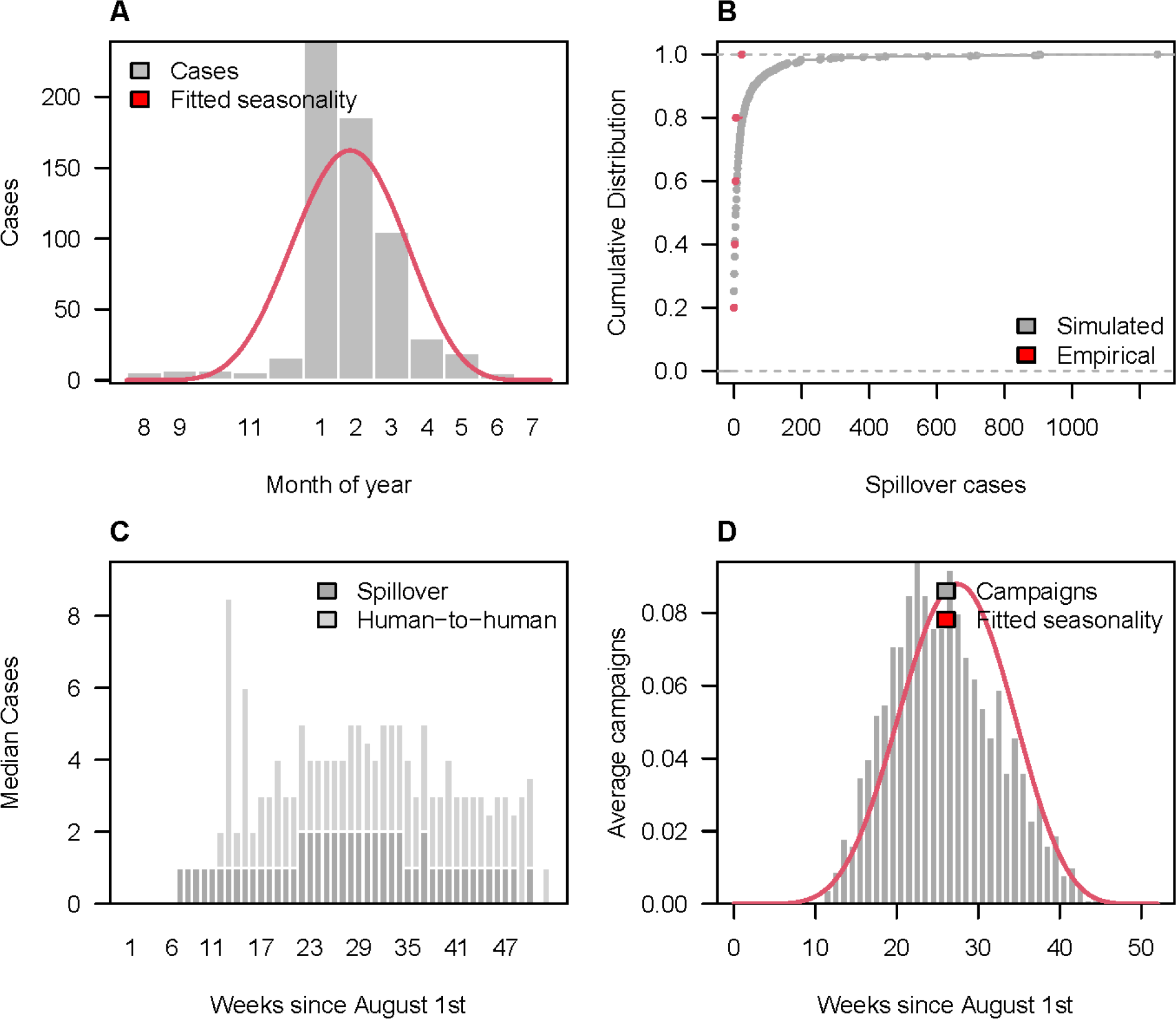
Spillover and reactive vaccination patterns for Nipah virus (NiV). (A) Observed weekly Nipah spillover cases (grey bars) and estimated seasonal spillover rate (red line). (B) Annual number of spillovers over the past 5 years (red) and cumulative distribution of simulated annual spillovers from 1000 replicates (grey). (C) Median weekly simulated spillover and human-to-human Nipah cases. (D) Average weekly number of reactive campaigns triggered via spillover detection compared to the estimated seasonal spillover rate (red line).

**Figure S4.**
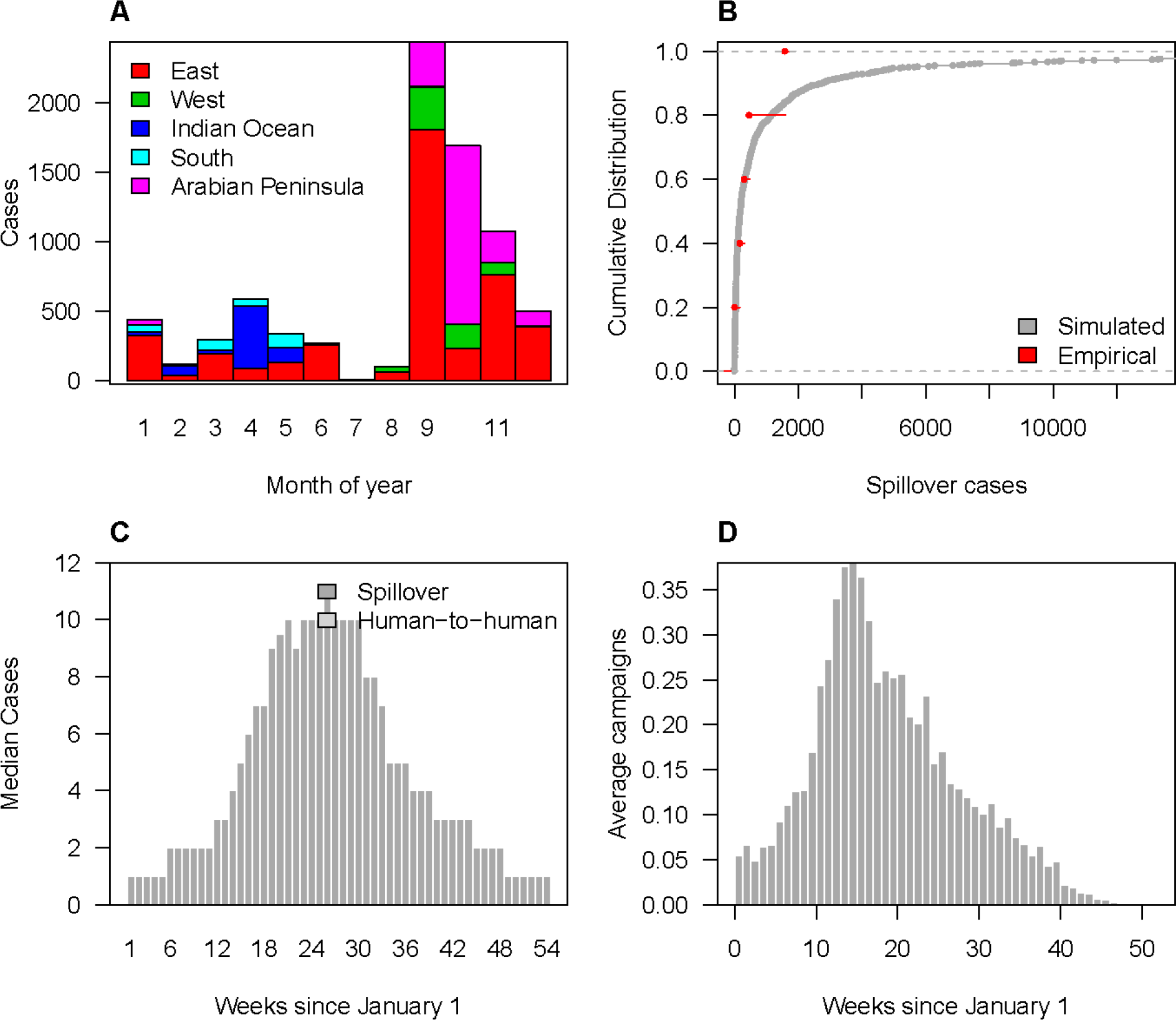
Spillover and reactive vaccination patterns for Rift Valley fever virus (RVFV). Observed monthly RVF spillover cases by region. (B) Annual number of spillovers over the past 5 years (red) and cumulative distribution of simulated annual spillovers from 1000 replicates (grey). (C) Median weekly simulated spillover and human-to-human RVF cases. (D) Average weekly number of reactive campaigns triggered via spillover detection. Fitted seasonality is not shown for RVFV as it was for the other pathogens because seasonality was fit separately for each region.

## SI Text

### 1. Sensitivity analysis: Analysis of reactive vaccination scenarios

In addition to our baseline reactive vaccination scenario, we examined the sensitivity of vaccination impact to varying different scenario parameters: the threshold number of cases needed to trigger a response, vaccination coverage in the general population, vaccination coverage of healthcare workers (HCWs), time from the threshold trigger to the start of vaccination, vaccine per exposure protection for one or two doses (PEP), and time delay from vaccination to protection (see Table 2 for default, low, and high parameter values). In addition, we examined the sensitivity of model results to the assumed or estimated value of R0 for each pathogen (SI Text section 2), and the impact of defining different catchment areas for vaccination (SI Text section 3). The results of the sensitivity analysis for MERS-CoV are summarized in the main text (Figure 6).

As expected, lowering the number of cases required to trigger a reactive vaccination campaign or increasing the percent of the population targeted for vaccination increased the number of vaccine regimens required for each pathogen (Figures S5-S8). These parameter values had a similar impact on the number of vaccine regimens needed to vaccinate HCWs (Figures S9-S12). Higher R0 values also increased the required number of vaccine regimens needed as higher human-to-human transmission increased the likelihood of the case threshold being exceeded. However, this impact was fairly small for LASV and RVFV due to their low R0 values (Figures S5,S8).

When vaccination impact was measured as the fraction of cases averted, the largest impact for each pathogen was achieved by lowering the threshold number of cases needed to trigger a reactive vaccination campaign (Figures 4A, S13A-15A). The second largest impact on the fraction of cases averted for each pathogen besides RVFV was achieved by decreasing the delay between a campaign being triggered and the start of vaccination (Figures 4A, S13A-S14A). For RVFV, the 2nd largest fraction of cases could be averted by increasing vaccination coverage in the general population or increasing vaccine PEP for a single dose vaccine (Figure S15A). Although lowering the response threshold maximized the fraction of cases averted for each pathogen, it did not maximize the number of cases averted per vaccine regimen administered, because lowering the threshold increased the number of vaccine campaigns and the required number of vaccine regimens (Figures S5-S8, 4B, S13B-S15B). For LASV, the largest per regimen impact was achieved by minimizing the delay prior to vaccination (Figure S13B). For NiV and RVFV, the largest per regimen impact was achieved by raising the response threshold (Figures S14B-S15B).

For each pathogen (besides RVFV, which is not associated with nosocomial transmission) vaccinating HCWs had a larger per regimen impact than vaccinating the general population (Figures 4C, S13C-S14C). Reducing the delay prior to vaccination and increasing vaccine PEP had the largest impact on the number of cases averted per HCW vaccinated (Figures 4C, S13C- S14C). The number of cases averted was slightly lower under our high-coverage of HCWs scenario because the high coverage of vaccination among HCWs was paired with low coverage among the general population (Figures 4A, S13A- S15A). However, this scenario would have achieved the highest number of cases averted per total number of vaccine regimens administered for LASV, MERS-CoV, and NiV.

### 2. Sensitivity analysis: Impact of R0

All R0 values and uncertainty ranges used in our analysis were either drawn from the literature or estimated from data (see Table 1 for parameter values and data sources). However, R0 estimates vary between studies and can also vary in space or time due to different environmental conditions or differences in human contact networks. Therefore, we also tested the sensitivity of our model results to lower and higher R0 values for each pathogen (for RVFV, the default R0=0, so only sensitivity to a higher value was examined). The number of total cases increased with R0 for each pathogen, with the largest sensitivity observed for MERS-CoV because the high estimate of R0 was close to 1 (Figure S16). There was also a large increase in the number of vaccine regimens required to vaccinate either the general population or HCWs for MERS-CoV at the higher R0 value, but the impact of R0 on the required number of vaccine regimens was minimal for the other pathogens (Figures S17-S18). As a result, there were minimal differences in the impact of vaccination under higher or lower R0 values for LASV, NiV, or RVFV (Figures S19-S22). Vaccination averted both a greater magnitude and a higher fraction of MERS cases as R0 increased (Figures S19-S20). In addition, the number of MERS cases averted per vaccine regimen administered to the general population or to HCWs also increased as R0 increased (Figures S21-S22). These results highlight the increasing potential effectiveness for reactive vaccination as a control strategy as R0 approaches 1 and larger outbreaks become more likely.

### 3. Model limitations

The goal of our analysis was to estimate vaccine stockpile needs and identify the most important determinants of success for reactive vaccination of zoonotic emerging pathogens. We modeled several different reactive vaccination strategies that are applicable to any zoonotic emerging pathogen, and tested this framework for four pathogens with differing epidemiologies. In addition, we explored the sensitivity of our results to different aspects of reactive vaccine deployment, such as the coverage level, deployment delays, and vaccine per exposure protection. However, there are some limitations to our approach that could affect these estimates.

First, we have a relatively poor understanding of the epidemiology of most emerging zoonotic pathogens, and data that could be used to try and elucidate the most important aspects of their epidemiologies is limited. Here we examined the impact of reactive vaccination for four pathogens with differing epidemiologies to try and capture how a range of epidemiological parameters (e.g., spillover rates, R0, etc.) affect vaccine stockpile requirements and the likely impact of vaccination. But there are still uncertainties surrounding the epidemiology of these pathogens that could affect the results of our analysis, such as the frequency of human-to-human transmission of MERS-CoV in community settings (Group and The WHO MERS-CoV Research Group 2013), or the route of NiV spillover to humans during recent outbreaks in India (Arunkumar et al. 2019). In addition, because no vaccines have been licensed for these pathogens yet, we had to make assumptions about key vaccine parameters (e.g., number of doses, time between vaccination and protection, and per exposure protection), based on the current vaccine target product profiles (TPPs) for each pathogen. We also had to make assumptions about the baseline reactive vaccination campaign parameter estimates such as campaign response time and duration (and best-case and worst-case scenarios for our sensitivity analysis). Assessing vaccine stockpile needs for newly emerged pathogens will involve even more uncertainty as epidemiological knowledge is critically limited immediately following emergence, as was demonstrated following the 2019 emergence of SARS-CoV-2 (Lee et al. 2020; Tindale et al. 2020). Our modeling approach can be applied to newly emerged zoonotic pathogens, but there will likely be a large amount of uncertainty regarding vaccine stockpile needs and where vaccination campaigns are most likely to occur.

A second, related, limitation, is that the modeling framework is intended to be applicable for a range of emerging zoonotic pathogens, and therefore cannot incorporate all of the specific epidemiological details that might affect vaccine demand or impact for a particular pathogen.

Third, we only considered reported cases when estimating pathogen spillover rates and human-to-human transmission because undiagnosed or unreported infections would not trigger an outbreak response. For several of the pathogens considered, however, the majority of infections--and even symptomatic cases--go unreported. A frequently cited study estimated that LASV infects 100,000-300,000 and kills 5,000 people annually (McCormick et al. 1987), and seroprevalence studies in several endemic areas indicate that spillover occurs much more frequently than reported (Kernéis et al. 2009; O’Hearn et al. 2016; Gibb et al. 2017).

Seroprevalence surveys for RVFV and MERS-CoV also indicate that these pathogens cause many unreported infections in at least some subpopulations (Müller et al. 2015; Munyua et al. 2021; Bron et al. 2021). Therefore, our estimate of reactive vaccination impact does not take into account the potential reduction in unobserved cases that would occur if at-risk populations were vaccinated. Improved surveillance could address this issue and would likely increase the frequency of reactive vaccination campaigns. This detection issue could also be partially addressed by adjusting the case threshold for outbreak response to account for the case detection probability, and then also adjusting vaccination impact to account for undetected infections.

Next, because the extent of community transmission for each of the study pathogens is poorly understood, we assumed that human-to-human transmission was limited to nosocomial settings. Although this could result in an underestimate of vaccine demand, our model simulations are consistent with epidemiological patterns observed to date (Figures S1-S4). We also assumed that all nosocomial transmission involved transmission from patient to HCWs or between HCWs and that there was no patient-to-patient or HCW-to-patient transmission.

Therefore our estimates of the impact of vaccinating HCWs represents the upper-bound on the effectiveness of this strategy as instances of patient-to-patient transmission would not be prevented via this strategy.

Another simplifying assumption of our model is that cases in one catchment area do not lead to transmission or an outbreak outside of that catchment area. However, imported cases of these pathogens have been reported. A MERS-CoV outbreak in South Korea derived from a spillover event in the Middle East (Park et al. 2016), a NiV outbreak in Singapore derived from a spillover event in Malaysia (Chan et al. 2002b), and cases of Lassa fever have been imported to Europe (Overbosch et al. 2020). These types of events have been rare, and none of these documented events resulted in an outbreak larger than the range of those that we simulated. Furthermore, documented outbreaks involving pathogen spread to neighbouring catchment areas are included in our datasets, and as such are to some extent captured in the current analysis. For example, our datasets include Lassa fever cases in Benin derived from an outbreak in the neighbouring adm2 located in Nigeria and RVFV outbreaks within multiple catchment areas of Tanzania that likely resulted from the movement of livestock (ProMED-mail 2020; Bron et al. 2021). Therefore, our model already implicitly incorporates the possibility of spread between-catchment areas, and although our model does not predict spillover cases occurring outside of each pathogen’s currently documented geographic distribution, the reactive vaccination strategies we examined should also be applicable for responding to imported cases and their associated outbreaks.

Another limitation is that we had to make several simplifying assumptions regarding the implementation of the reactive vaccination campaigns. One such simplification was assuming that all vaccine doses (per regimen) were administered on the same day. This is likely an unrealistic assumption for mass vaccination campaigns, particularly those that cover large geographic areas. Relaxing this assumption would reduce the public health impact of reactive vaccination in the same manner that delays in the start of the vaccination campaign did in our analysis. Another simplification is that we did not consider any targeted vaccination strategies besides targeting healthcare workers to limit nosocomial outbreaks or ring vaccination around index cases. For the ring vaccination we calculated the number of index cases that would trigger a ring vaccination response, but we did not model the impact of this response. Besides these two strategies, there might be other targeted vaccination approaches that would require a smaller vaccine stockpile than targeting the general population while still producing a substantial public health impact. For example, in the case of RVFV, a potential vaccination strategy might include targeting high-risk groups such as veterinarians, butchers, and livestock holders. One reason we did not consider this strategy is because of the coarseness or absence of the data available on these professions. While veterinarians only constitute a small proportion of the at-risk population, their higher risk of acquiring infections could increase the impact of a campaign that targeted them for vaccination. This could also increase the safety of those that are often at the frontline of an outbreak response. Similar targeted strategies might be envisioned for camel workers in areas where MERS-CoV is endemic in livestock, or individuals who collect or consume date palm sap in India and Bangladesh (Dudas et al. 2018; Islam et al. 2016).

Finally, spillover cases were distributed over catchment areas representing 2nd administrative districts (or hospitals within the 1st administrative units in our sensitivity analysis), irrespective of the urban/rural nature of the catchment area. This may result in an overestimation of the population at risk and thus the number of regimens needed. Simulating the spillover rates per 2nd administrative unit, instead of at the 1st administrative level, could improve the estimation of reactive vaccine demand. However, the adm2 location of spillover cases were not available most of the time.

---

**Figure S5.**
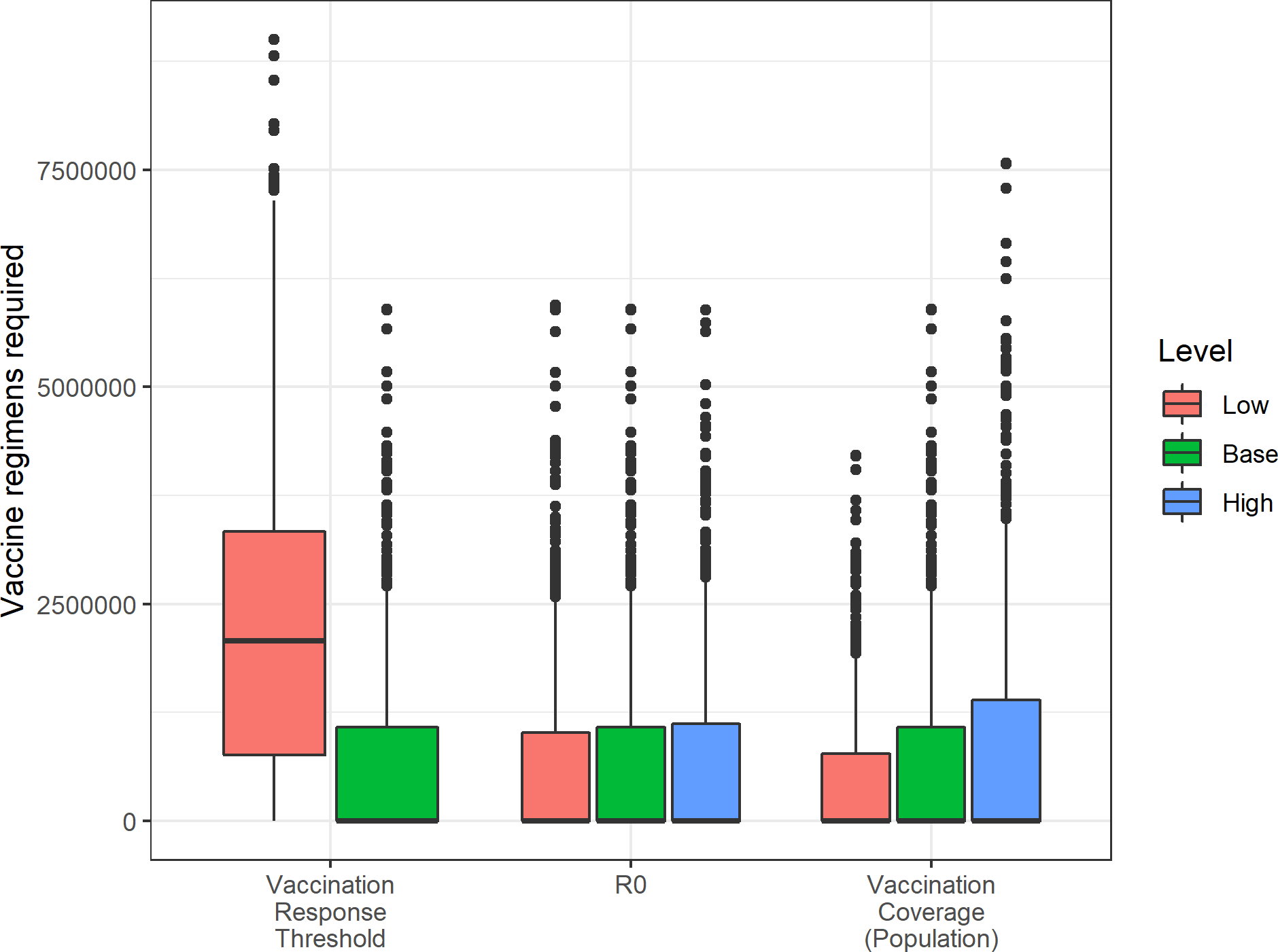
Vaccine regimens required for Lassa fever virus (LASV). The impact of varying several model parameters on the number of vaccine regimens required to meet reactive vaccination campaign targets. Base refers to the default scenario used in our main analysis. See Table 2 for specific parameter values.

**Figure S6.**
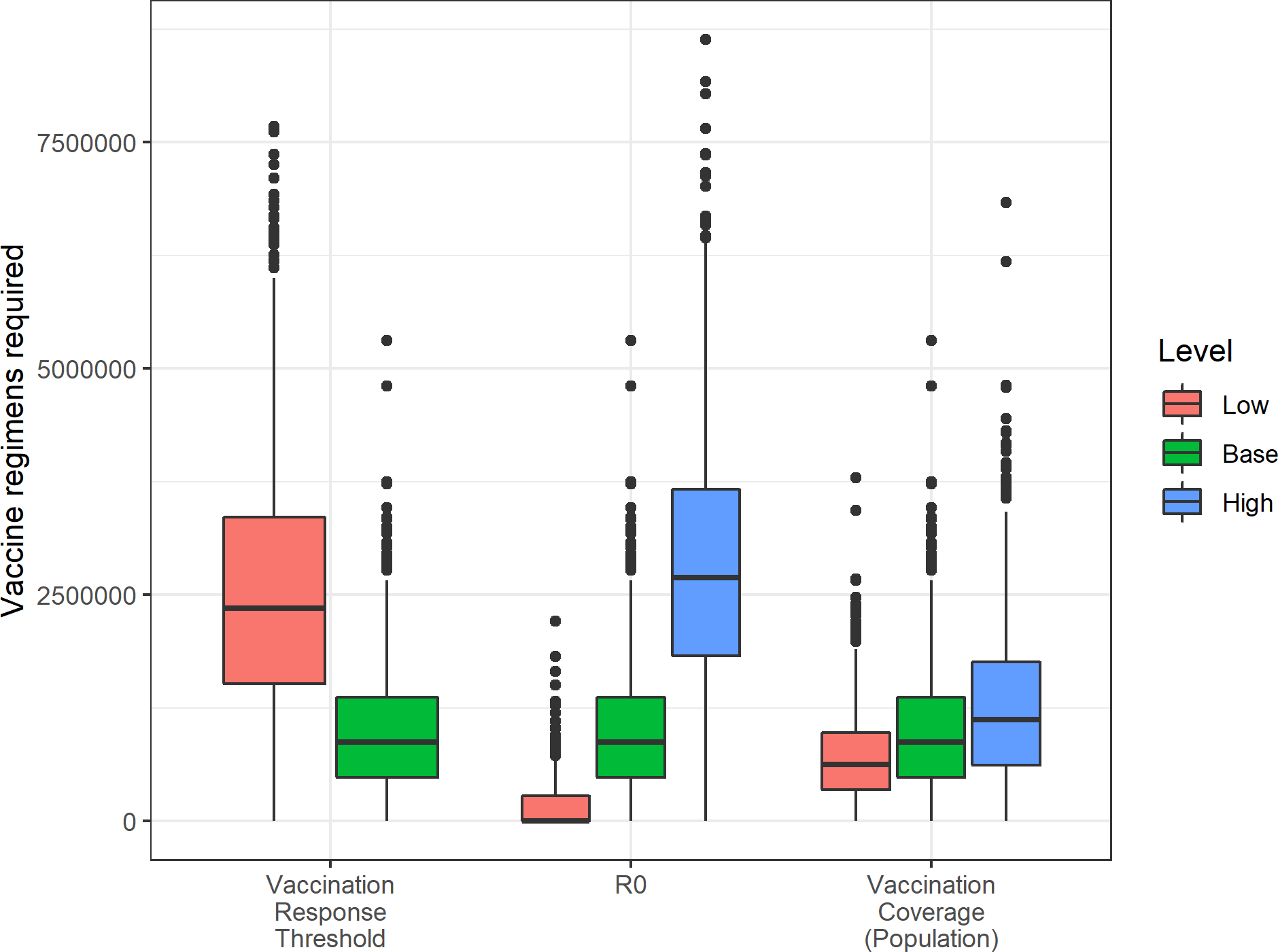
Vaccine regimens required for Middle Eastern respiratory virus (MERS-CoV). The impact of varying several model parameters on the number of vaccine regimens required to meet reactive vaccination campaign targets. Base refers to the default scenario used in our main analysis. See Table 2 for specific parameter values.

**Figure S7.**
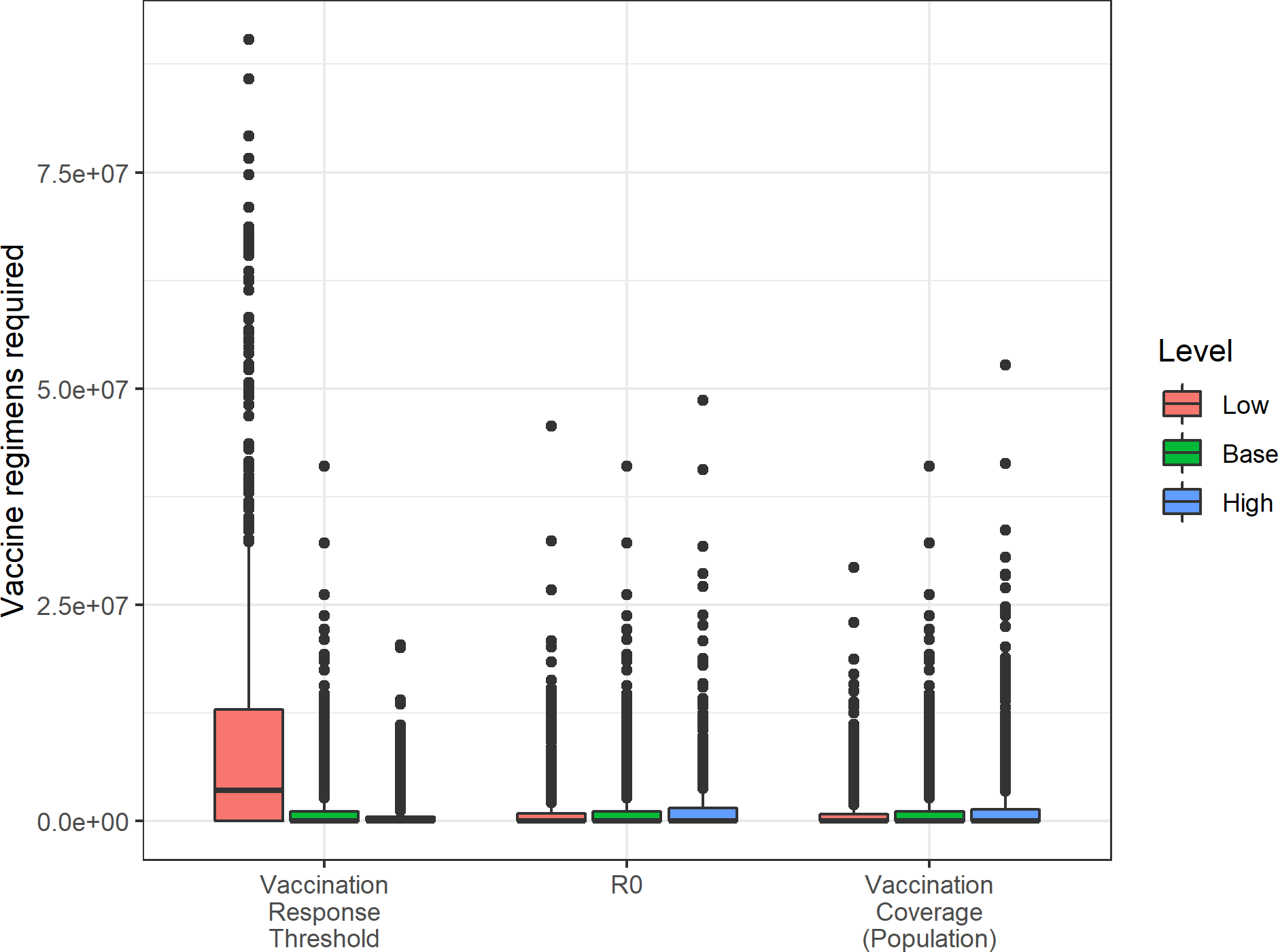
Vaccine regimens required for Nipah virus (NiV). The impact of varying several model parameters on the number of vaccine regimens required to meet reactive vaccination campaign targets. Base refers to the default scenario used in our main analysis. See Table 2 for specific parameter values.

**Figure S8.**
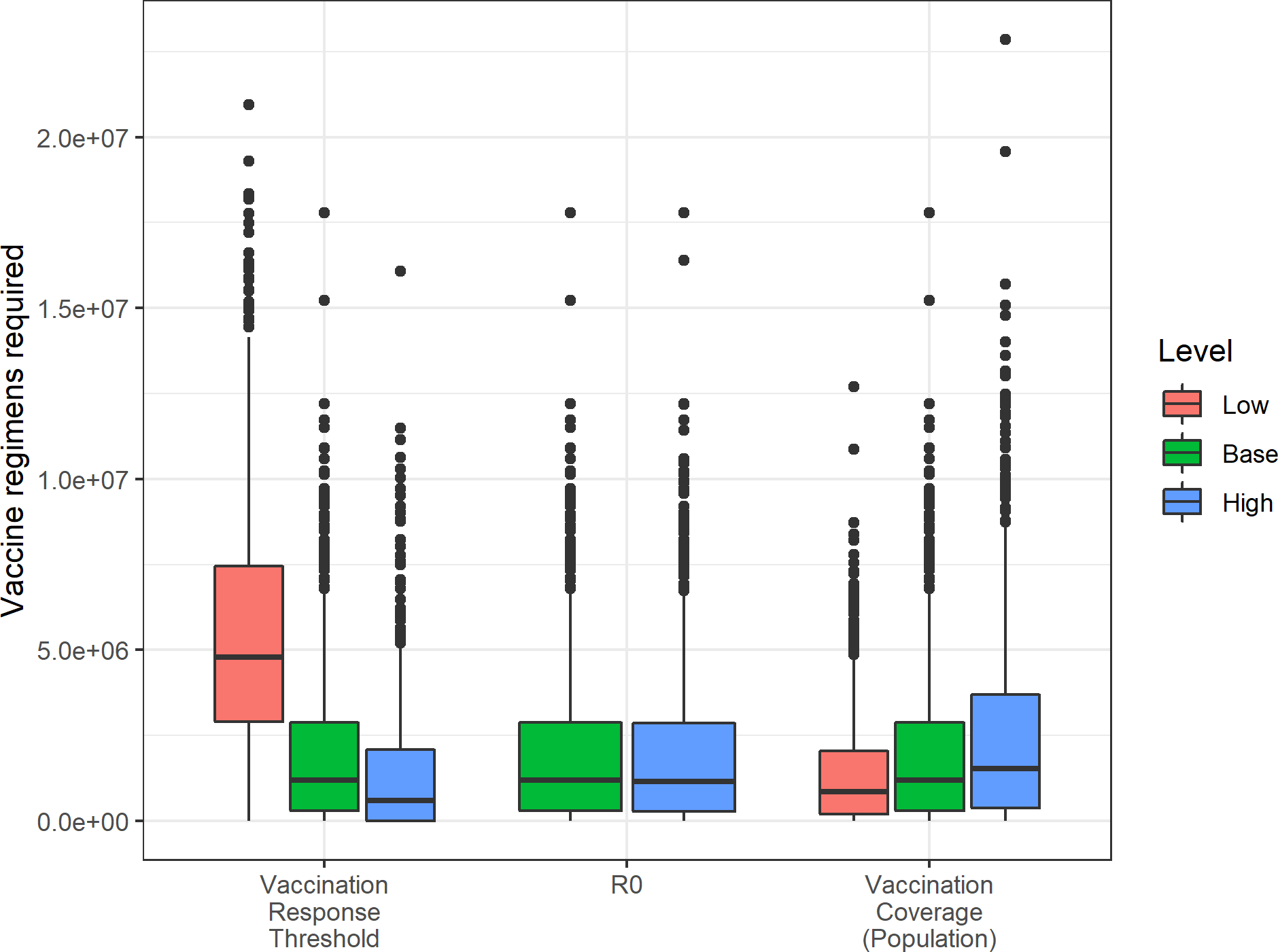
Vaccine regimens required for Rift Valley fever virus (RVFV). The impact of varying several model parameters on the number of vaccine regimens required to meet reactive vaccination campaign targets. Base refers to the default scenario used in our main analysis. See Table 2 for specific parameter values.

**Figure S9.**
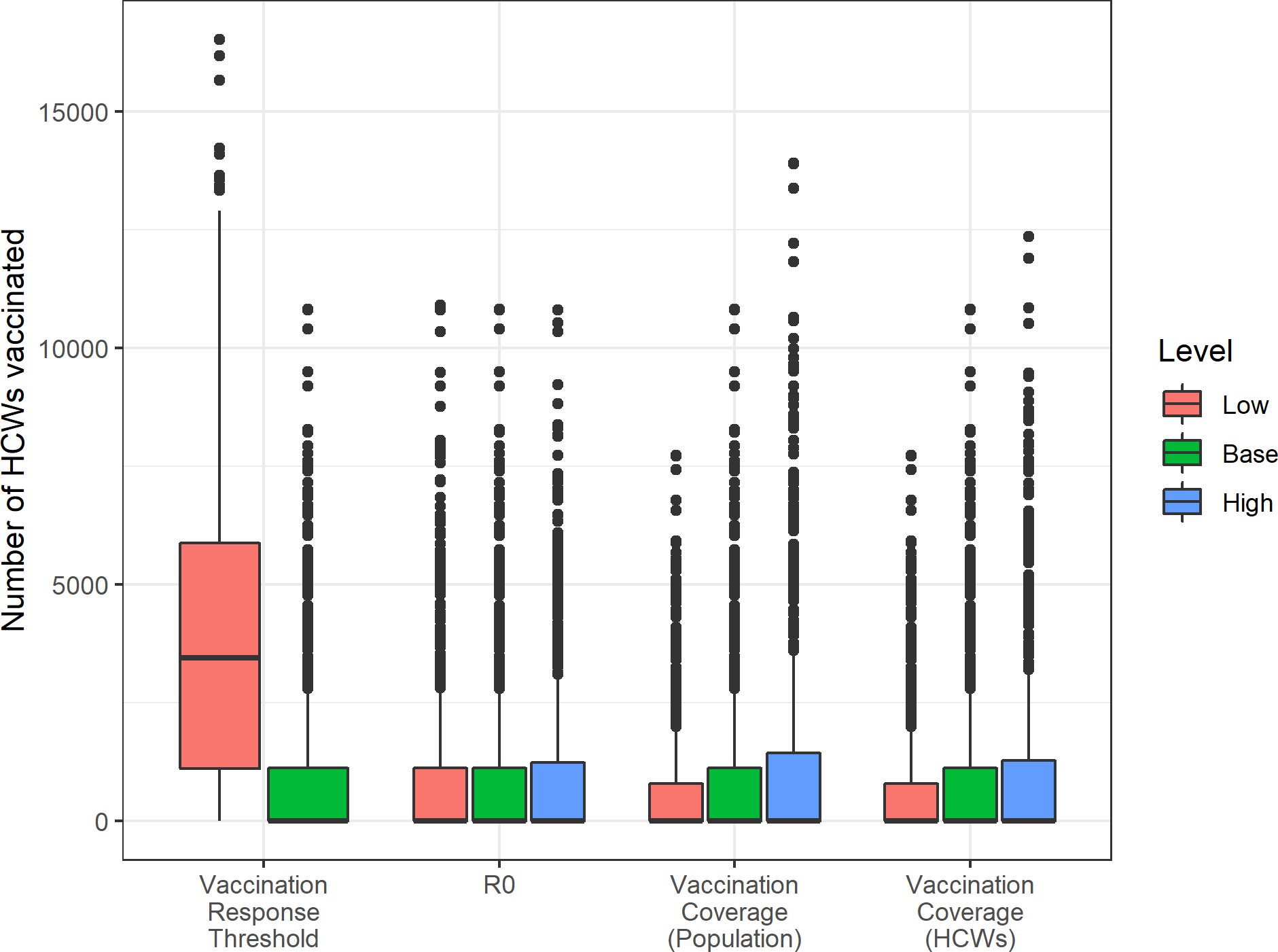
Vaccine regimens required to vaccinate healthcare workers for Lassa fever virus (LASV). The impact of varying several model parameters on the number of vaccine regimens required to meet reactive vaccination campaign targets among healthcare workers (HCWs). Base refers to the default scenario used in our main analysis. See Table 2 for specific parameter values.

**Figure S10.**
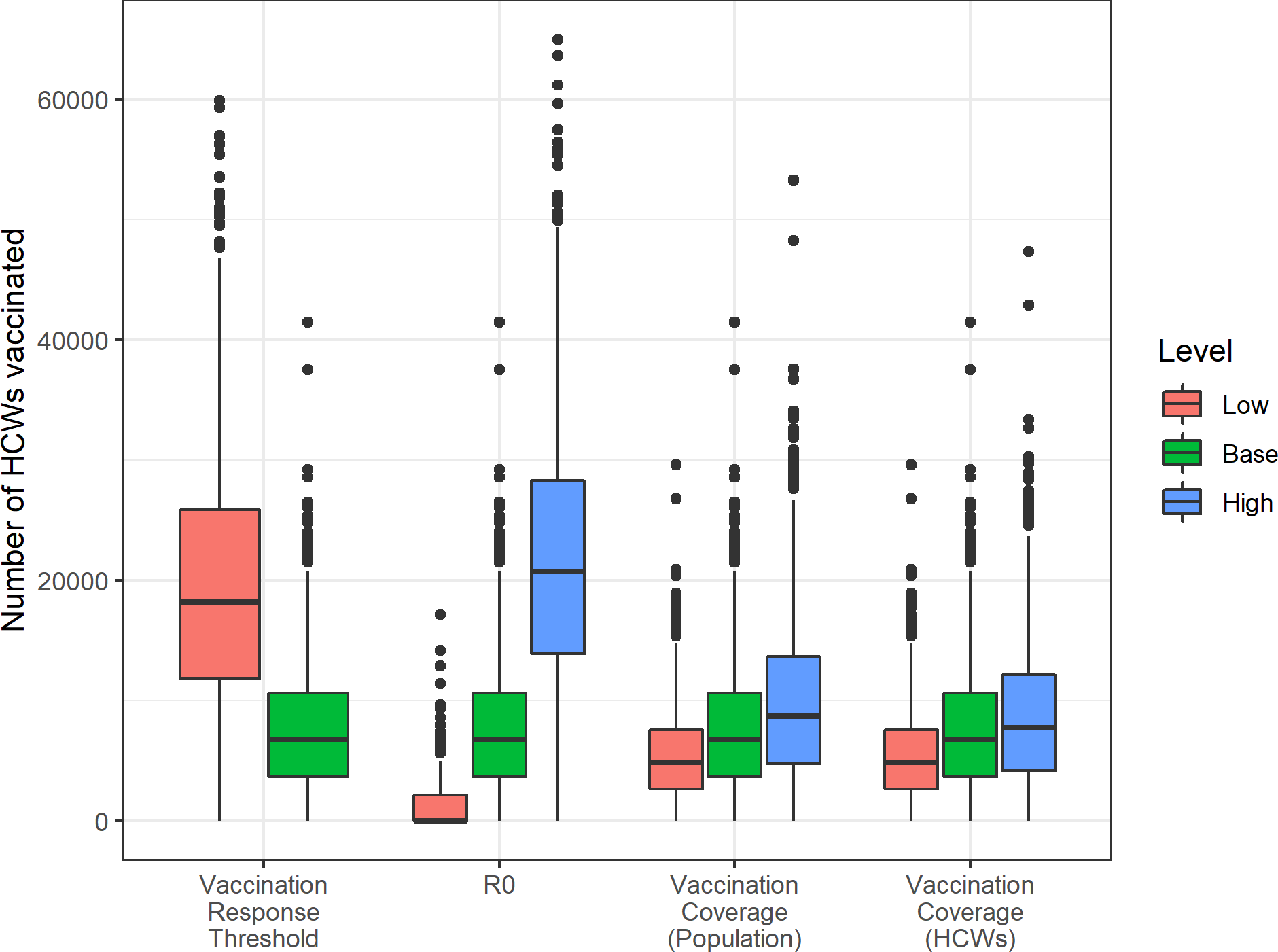
Vaccine regimens required to vaccinate healthcare workers for Middle Eastern respiratory virus (MERS-CoV). The impact of varying several model parameters on the number of vaccine regimens required to meet reactive vaccination campaign targets among healthcare workers (HCWs). Base refers to the default scenario used in our main analysis. See Table 2 for specific parameter values.

**Figure S11.**
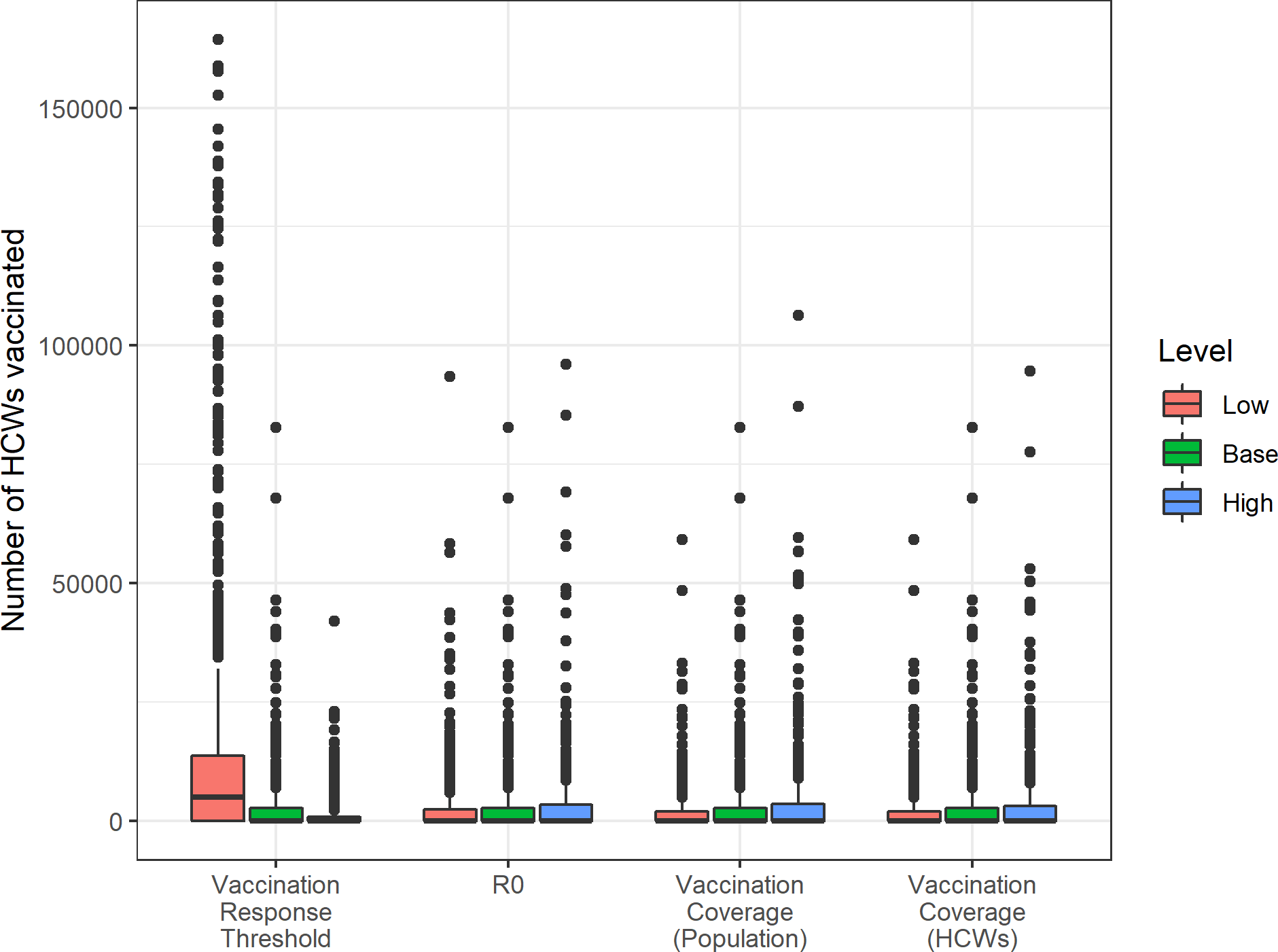
Vaccine regimens required to vaccinate healthcare workers for Nipah virus (NiV). The impact of varying several model parameters on the number of vaccine regimens required to meet reactive vaccination campaign targets among healthcare workers (HCWs). Base refers to the default scenario used in our main analysis. See Table 2 for specific parameter values.

**Figure S12.**
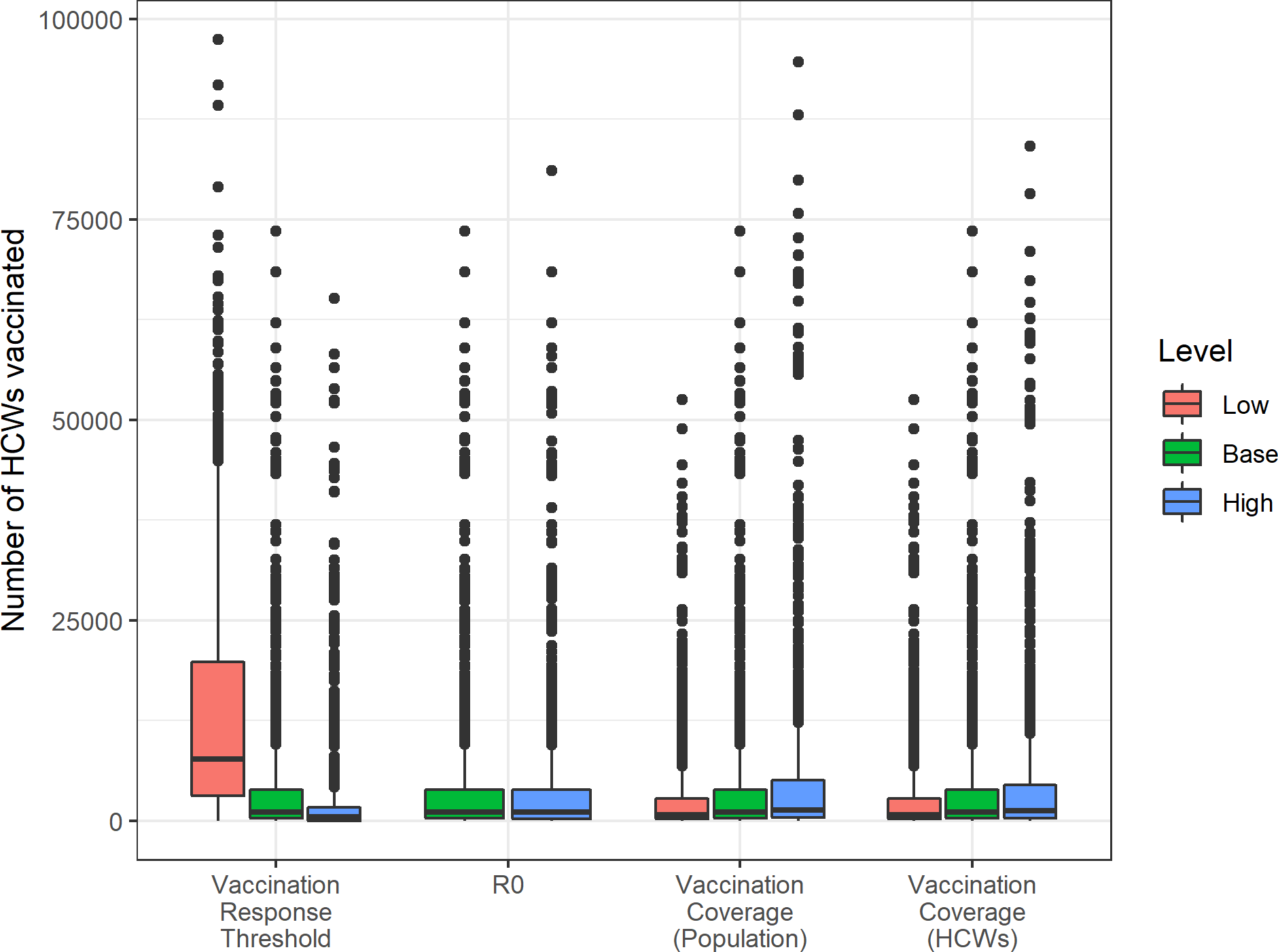
Vaccine regimens required to vaccinate veterinarians for Rift Valley fever virus (RVFV). The impact of varying several model parameters on the number of vaccine regimens required to meet reactive vaccination campaign targets among veterinarians (HCWs). Base refers to the default scenario used in our main analysis. See Table 2 for specific parameter values.

**Figure S13.**
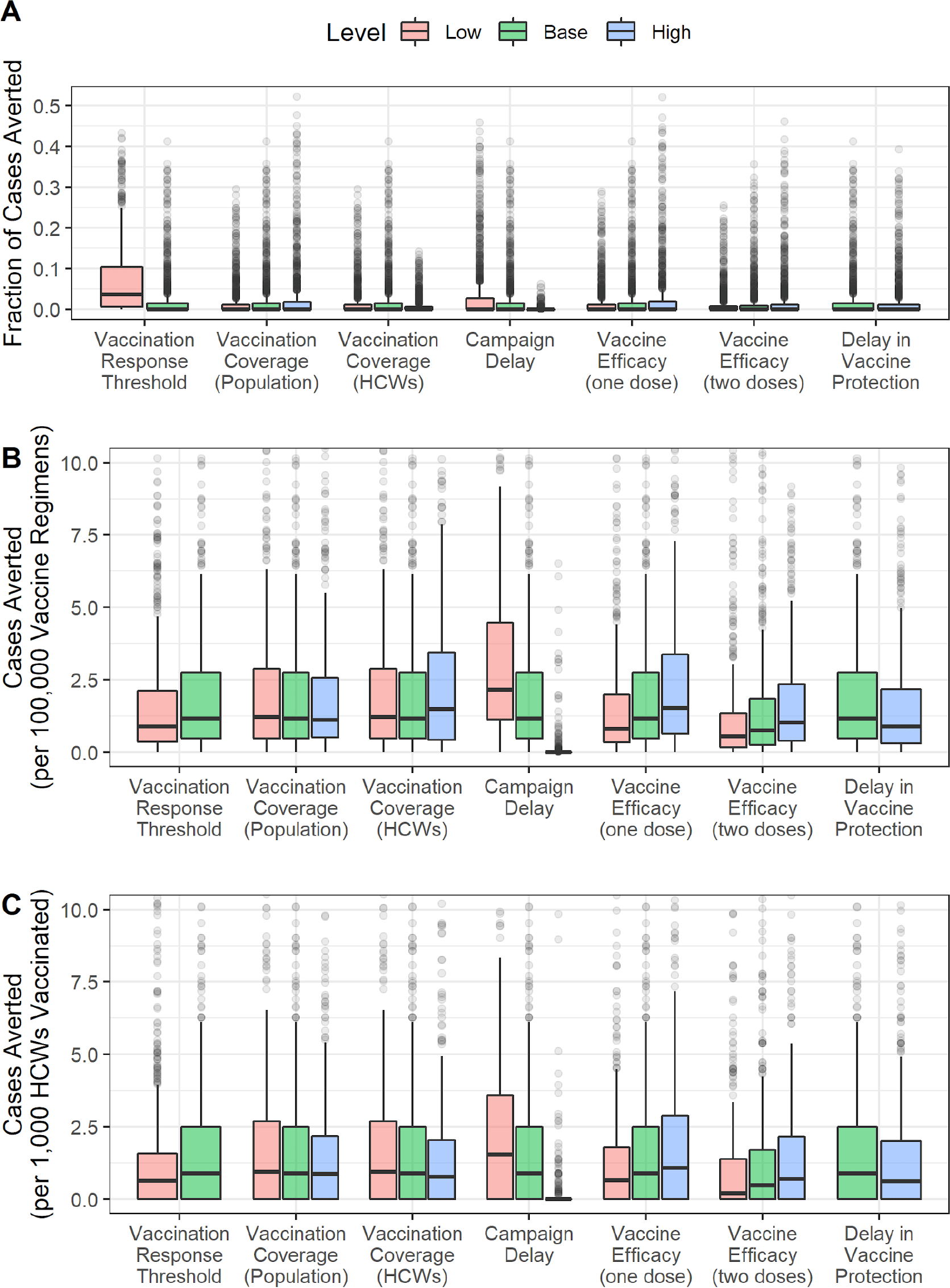
Vaccination impact sensitivity analysis for LASV. Sensitivity of vaccination impact for LASV to variation in different campaign parameters expressed as (A) fraction of cases averted, (B) cases averted per 100,000 vaccinated in the general population, and (C) cases averted per 1,000 health care workers (HCWs) vaccinated.

**Figure S14.**
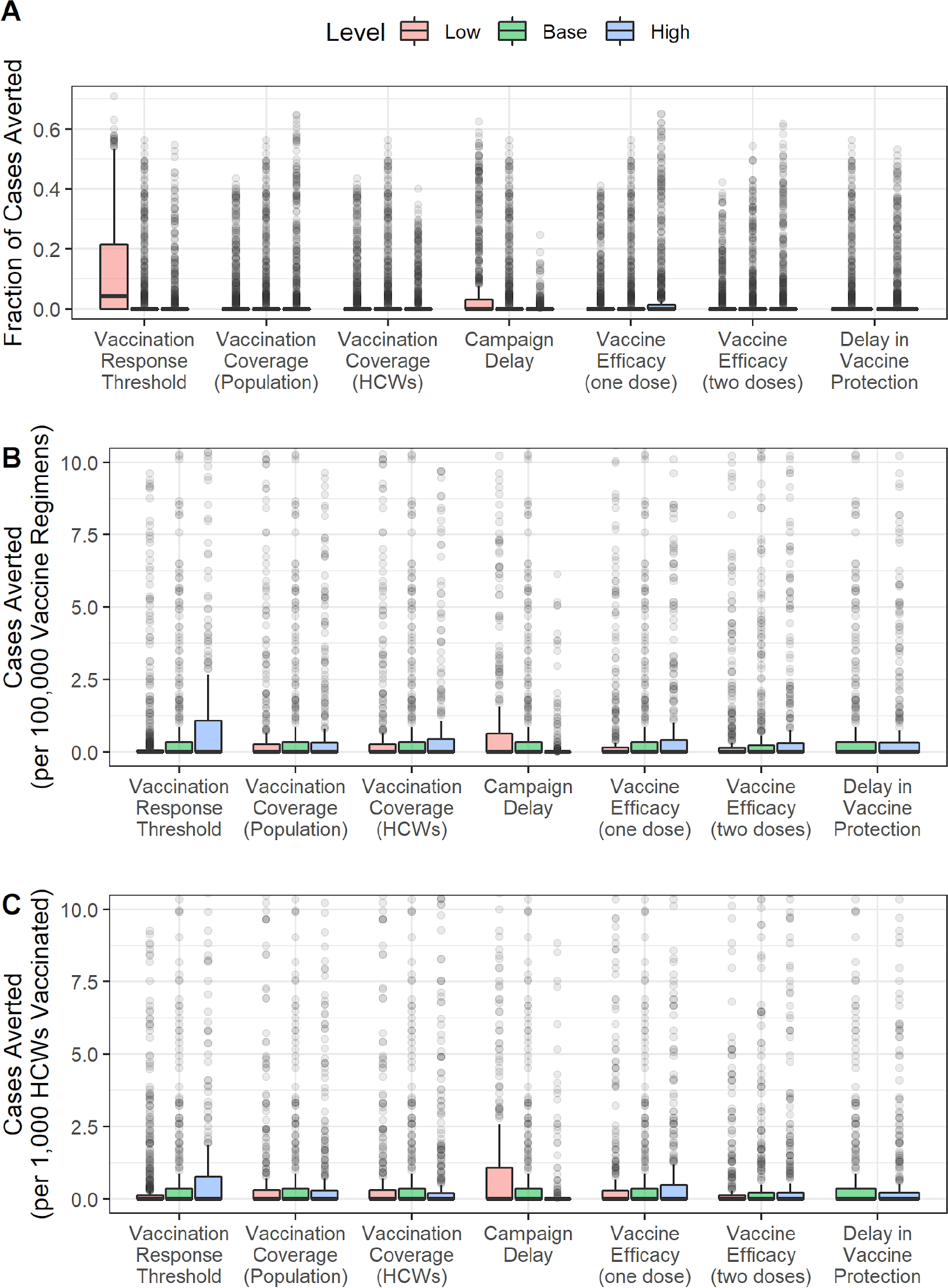
Vaccination impact sensitivity analysis for NiV. Sensitivity of vaccination impact for NiV to variation in different campaign parameters expressed as (A) fraction of cases averted, (A) cases averted per 100,000 vaccinated in the general population, and (C) cases averted per 1,000 health care workers (HCWs) vaccinated.

**Figure S15.**
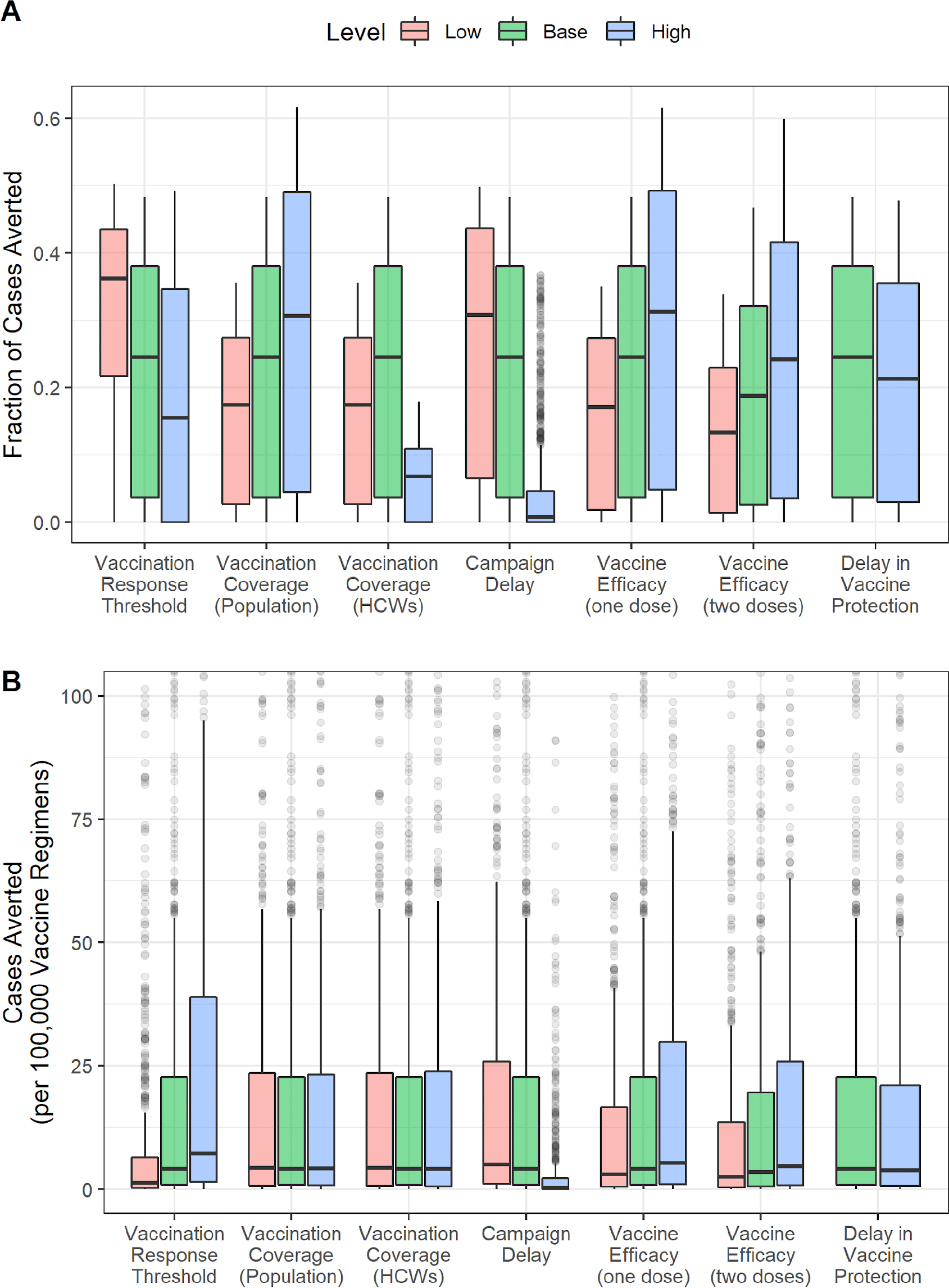
Vaccination impact sensitivity analysis for RVFV. Sensitivity of vaccination impact for RVFV to variation in different campaign parameters expressed as (A) fraction of cases averted, (B) cases averted per 100,000 vaccinated in the general population, and (C) cases averted per 1,000 health care workers (HCWs) vaccinated.

**Figure S16.**
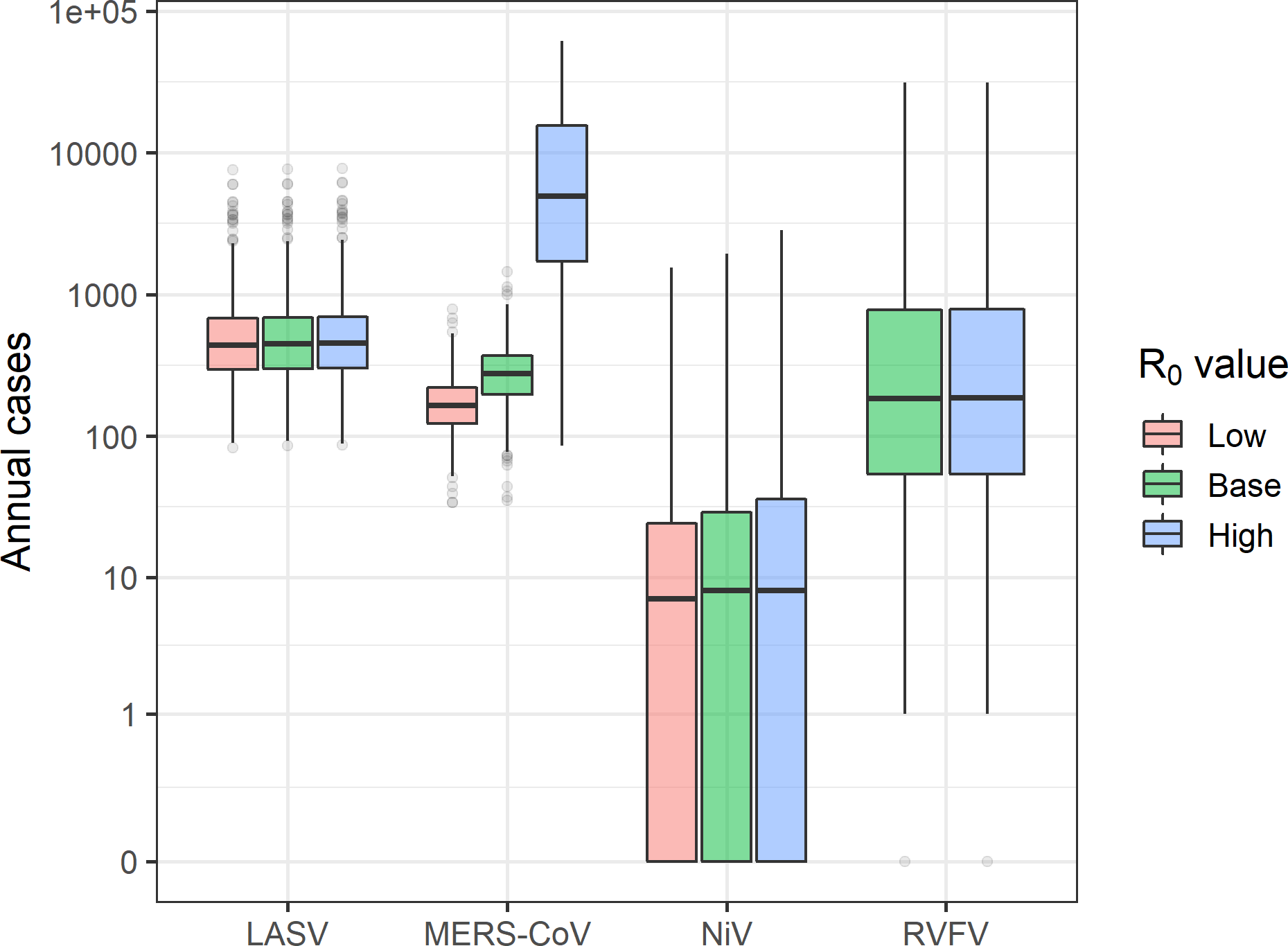
Number of cases under different R0 assumptions.

**Figure S17.**
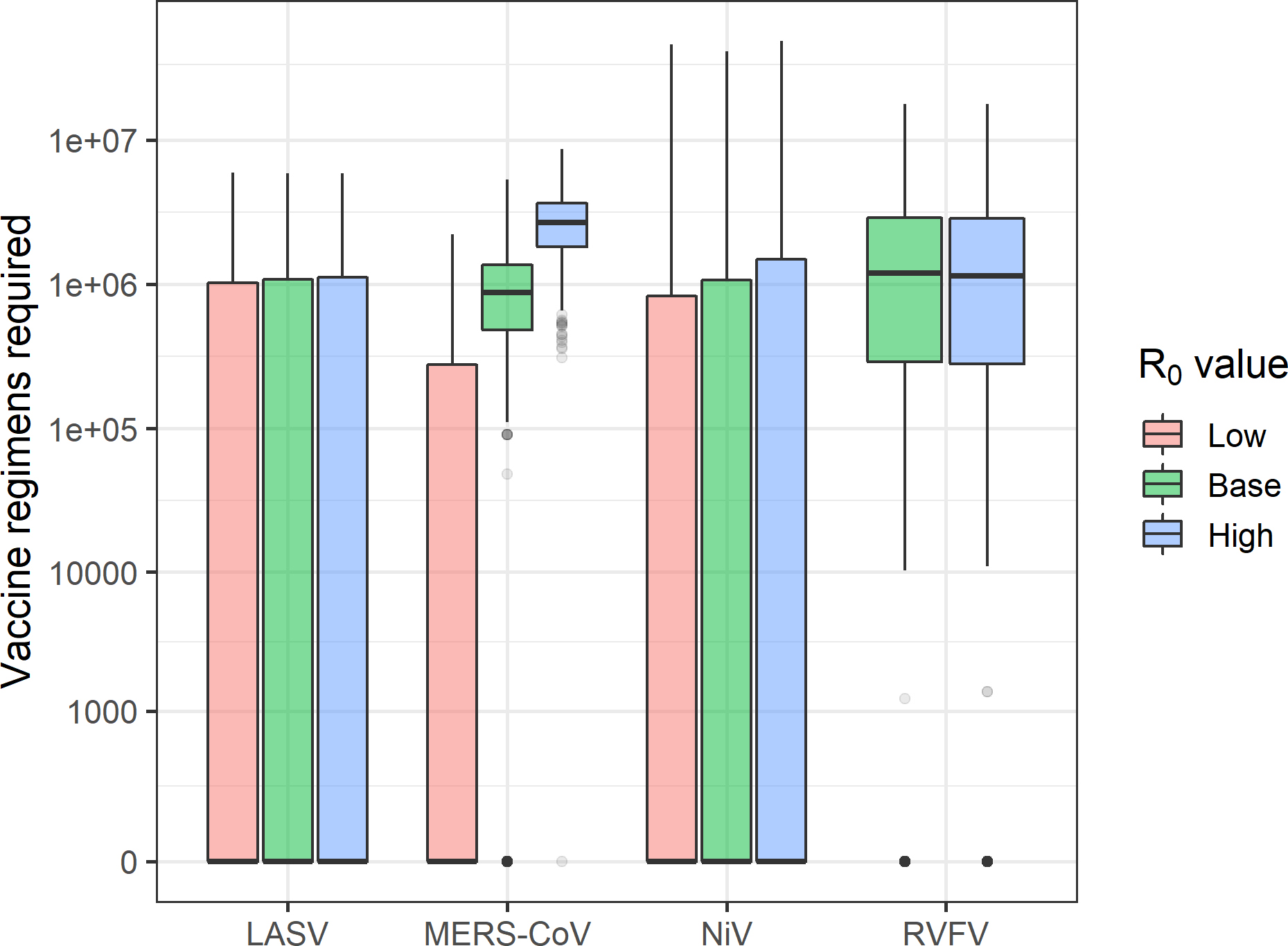
Number of vaccine regimens required under different R0 assumptions.

**Figure S18.**
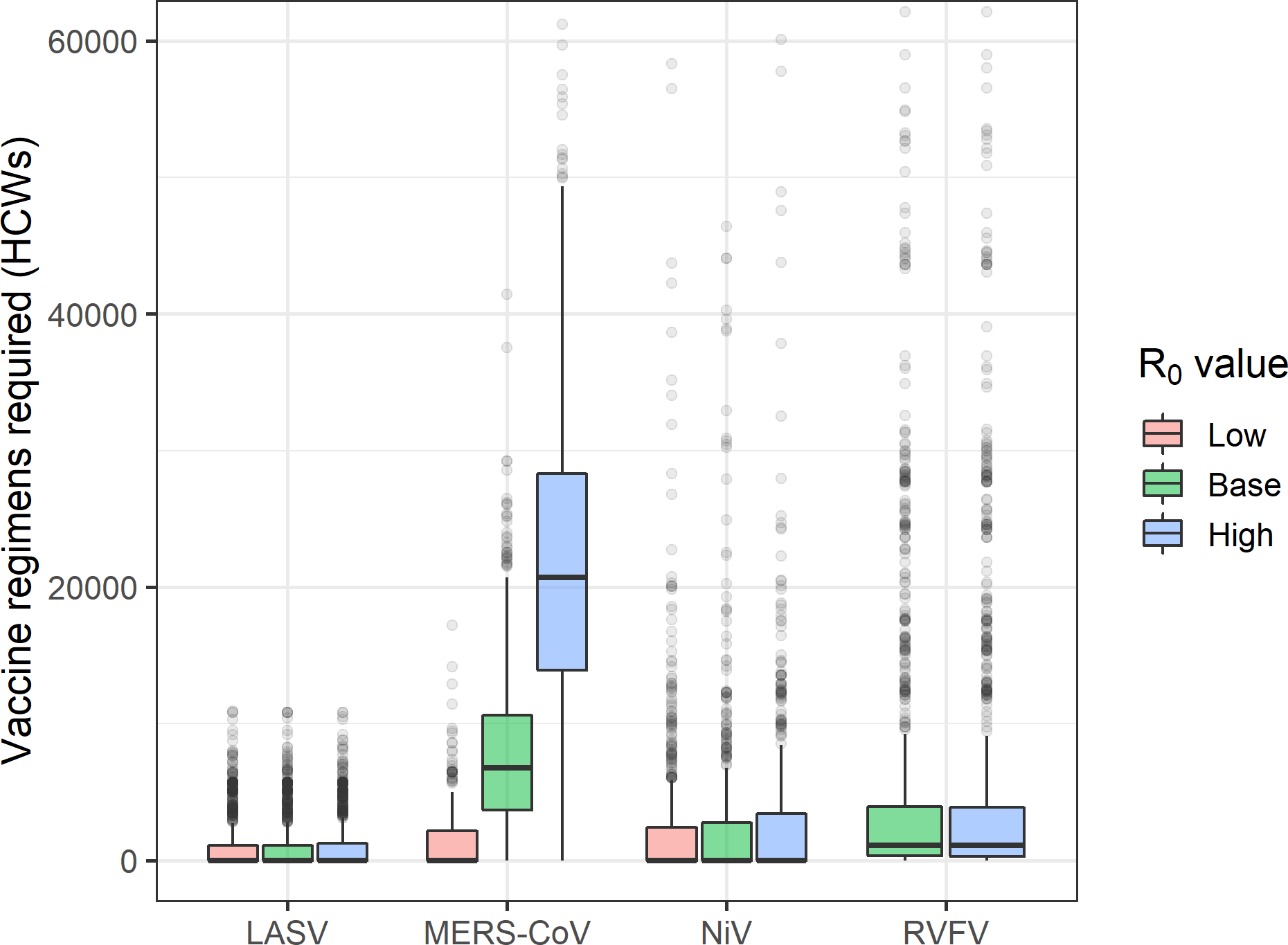
Number of vaccine regimens required for healthcare workers (HCWs) under different R0 assumptions.

**Figure S19.**
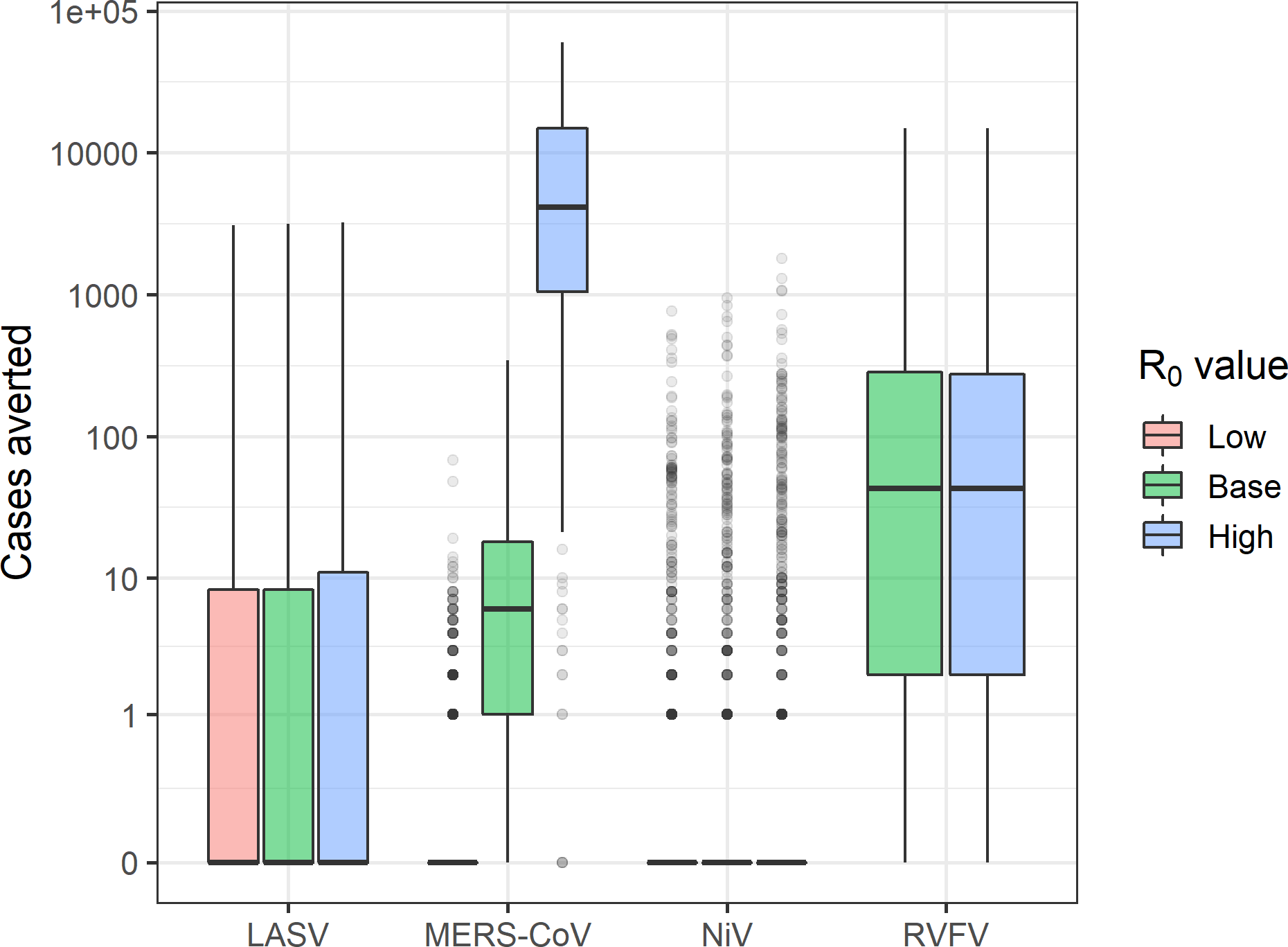
Number of cases averted by vaccinating the general population under different R0 assumptions.

**Figure S20.**
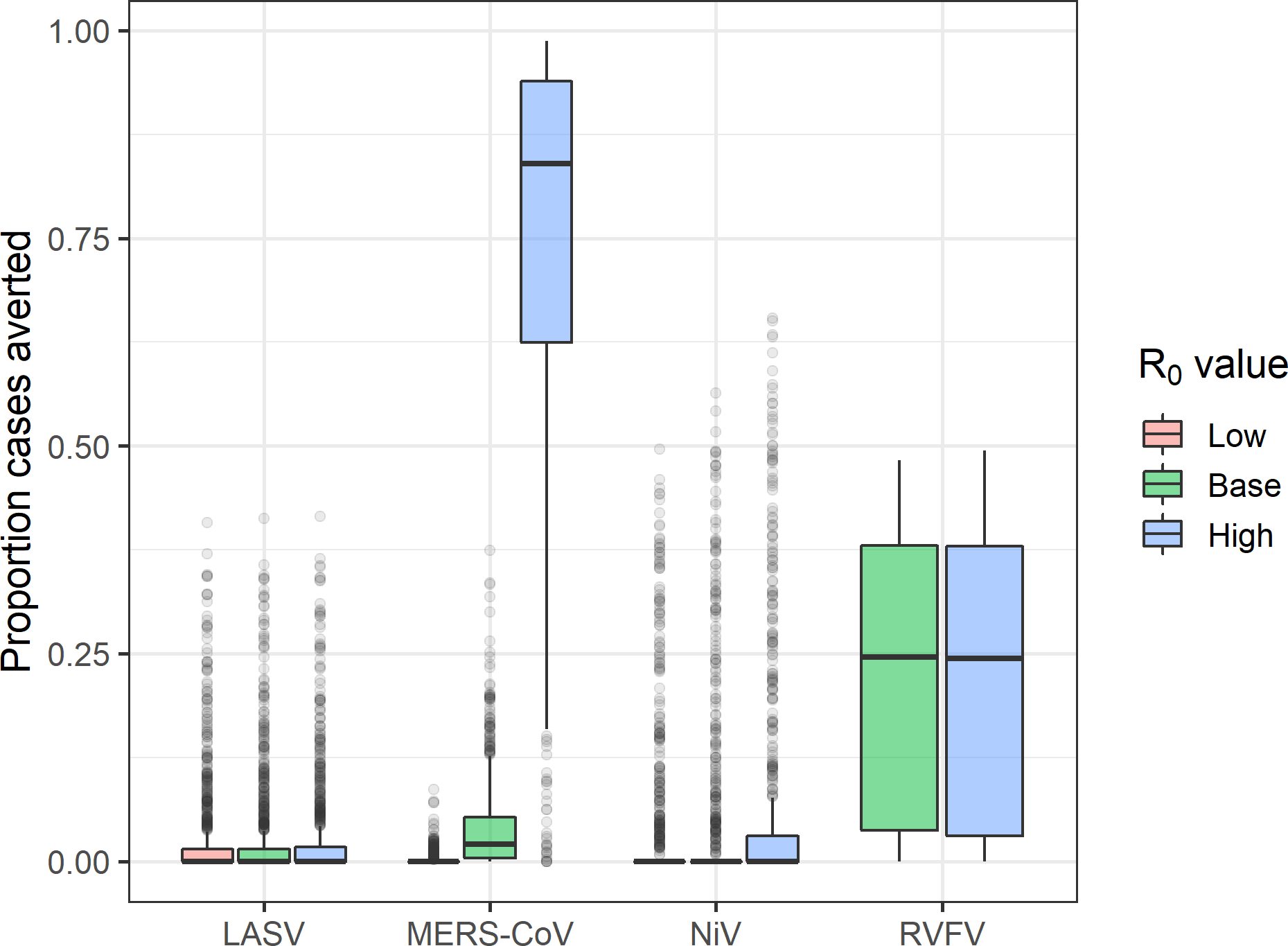
Fraction of cases averted by vaccinating the general population under different R0 assumptions.

**Figure S21.**
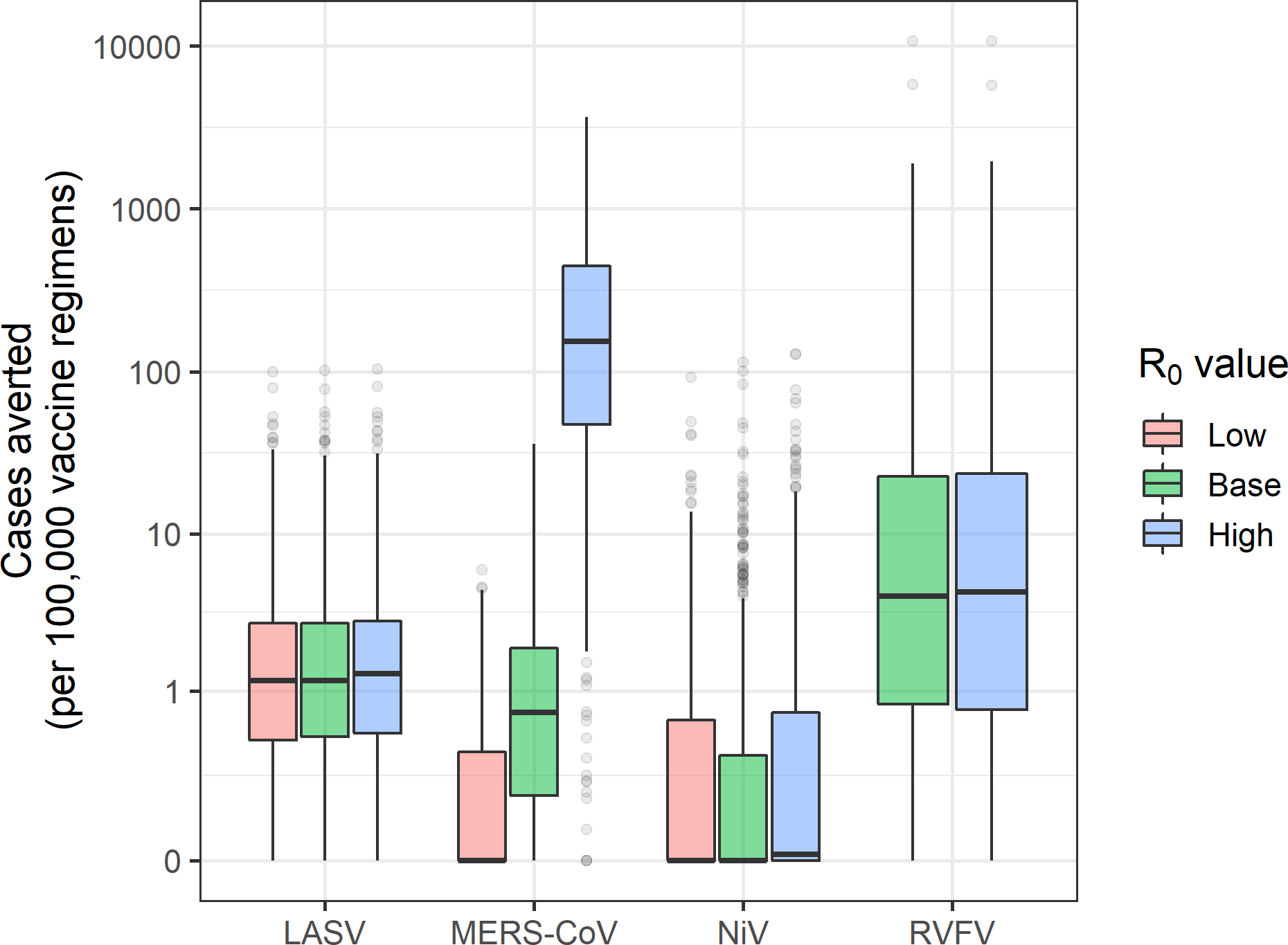
Number of cases averted per vaccine regimen administered to the general population under different R0 assumptions.

**Figure S22.**
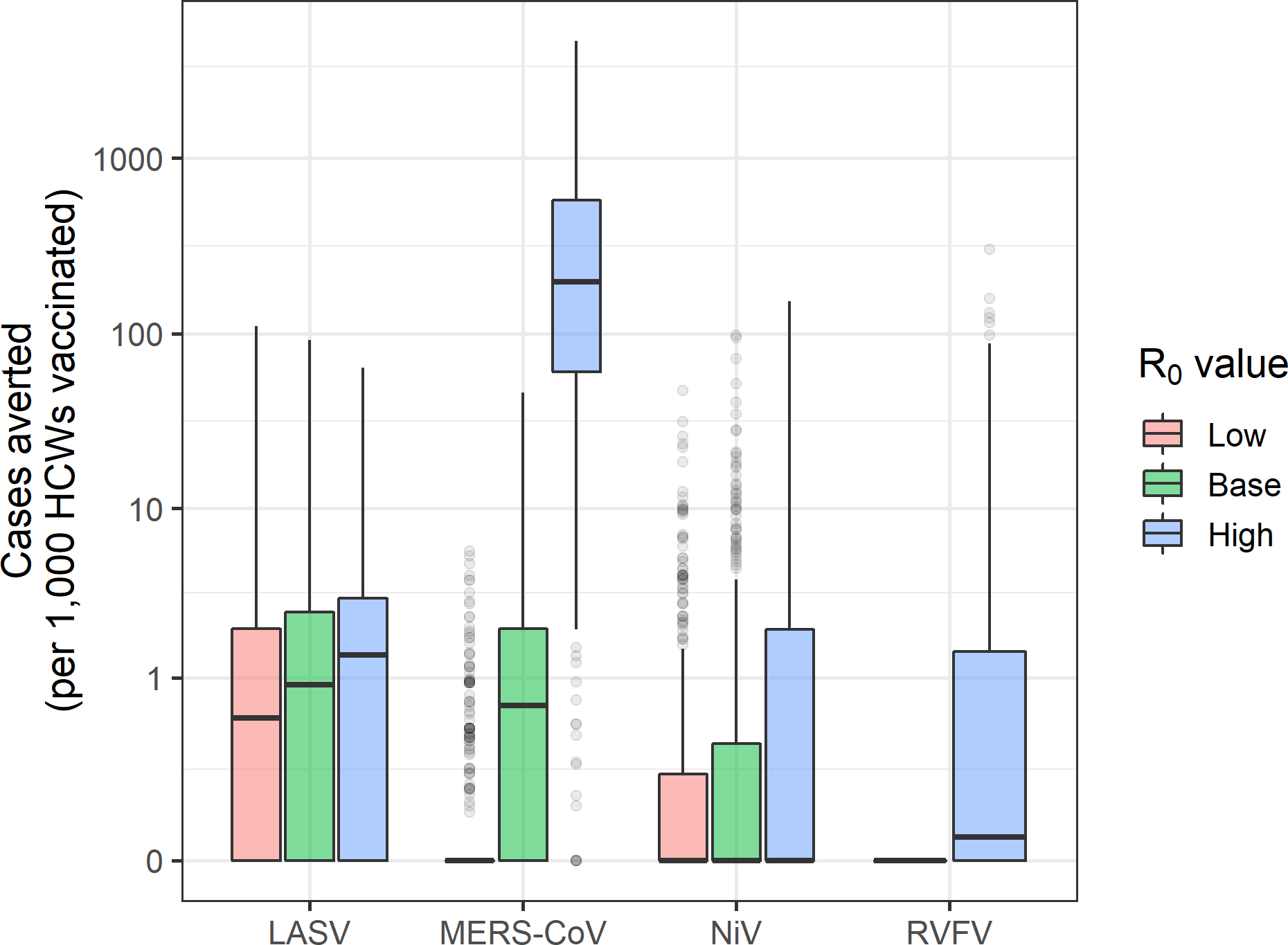
Number of cases averted per vaccine regimen administered to healthcare workers (HCWs) under different R0 assumptions.

## 3. Analysis of different spillover and vaccination catchment areas

In our analysis we estimated spillover rates for each pathogen at the 1st administrative level (adm1). We then accounted for spatial clustering of cases below the adm1 level by associating each simulated case with a catchment area. In the main analysis, catchment areas were defined as the 2nd administrative units (adm2) within each adm1. For countries with no 2nd administrative level, hospitals within an adm1 were treated as catchment areas (hospitals within 10 km were combined into a single catchment area). This catchment area definition produced 1570 catchment areas for LASV, 767 for MERS-CoV, 5076 for NiV and 2126 for RVFV. Here we consider two alternative catchment area definitions: (1) treating all adm1 units as unique catchment areas, and (2) treating all hospitals within adm1 units as unique catchment areas (with hospitals <10km apart combined into a single catchment area). The adm1 catchment area definition resulted in 214 catchment areas for LASV, 82 for MERS-CoV, 375 for NiV and 343 for RVFV. The adm1 hospitals catchment area definition produced 1749 catchment areas for LASV, 3138 for MERS-CoV, 10799 for NiV and 4722 for RVFV. Therefore the adm1 catchment areas are larger than the adm2 catchment areas, while the adm1 hospitals catchment areas are generally smaller than the adm2 catchment areas. Because spillover rates were estimated at the adm1 level, spillovers within an adm1 unit were allocated via a multinomial distribution to all hospital catchment areas within that adm1 unit, with a probability of 1/(# of catchment areas within adm1).

The number of spillover cases remained the same under the different catchment area definitions, but the frequency, timing, and location of reactive vaccination campaigns were shifted. Reactive vaccination campaigns tended to be triggered sooner during the transmission season for adm1 catchment areas because these catchment areas covered a broader area and larger population (Figures S23-S26). There were only minor differences in the timing of vaccination campaigns between the adm2 and adm1 hospitals catchment areas (Figures S1-S4, S27-S30). The geographic distribution of spillovers was less clustered for the larger adm1 catchment areas, but more finely distributed and clustered for the adm1 hospital catchment areas, particularly in countries like South Africa and Madagascar with large 1st-level administrative regions (Figures 2A, S31A-S32A). A similar geographic pattern was observed for the location of reactive vaccination campaigns (Figures 2B, S31B-S32B).

The total number of human-to-human cases did not differ by catchment area definition for any of the pathogens (Figures 1A, S33A-S34A). The median number of reactive vaccination campaigns was lower using adm1 catchment areas than adm2 catchment areas for MERS-CoV (3; 95% PrI: 1-6 vs. 4; 95% PrI: 0-11) and RVFV (3; 95% PrI:0-6 vs. 5; 95% PrI: 0-20) (Figures 1B, S33B). In contrast, the median number of reactive vaccination campaigns were higher using adm1 catchment areas for LASV (3; 95% PrI: 1-6 vs. 0; 95% PrI: 0-20) and NiV (1; 95% PrI: 0-3 vs. 0; 95% PrI: 0-8). The frequency of years with no reactive vaccination campaigns was lower using adm1 catchment areas compared to adm2 or adm1 hospital catchment areas for all four pathogens. Both the median number of reactive vaccination campaigns and the frequency of years with no campaigns were similar using adm1 hospital catchment areas compared to the baseline adm2 catchment areas, although the frequency of years with no campaigns was slightly higher for the adm1 hospital catchment areas for all four pathogens (Figures 1B, 34B).

The number of vaccine regimens required to cover the general population were significantly higher using adm1 catchment areas versus adm2 or adm1 hospital catchment areas for (Figures 1C, S33C-34C). For MERS-CoV, the median required number of regimens increased from 286,259 (95% PrI: 0-855,099) for adm1 hospital catchment areas, to 1,242,922 (95% PrI: 0-4,062,010) for adm2 catchment areas, and 14,303,325 (95% PrI: 1,416,969) for adm1 catchment areas. For LASV, the median required number of regimens increased from 0 (95% PrI: 0-4,452,966) for adm1 hospital catchment areas and 0 (95% PrI: 0-5,184,360) for adm2 catchment areas, to 11,603,802 (95% PrI: 2,953,628-25,277,594) for adm1 catchment areas. The number of vaccine regimens needed to vaccinate healthcare workers (HCWs) was also lowest for adm1 hospital catchment areas and highest for adm1 catchment areas (Figures 1C, S33C-S34C).

The total number of cases averted via vaccination for each pathogen was also lowest using adm1 hospital catchment areas and highest using adm1 catchment areas (Figures 1D, S33D-S34D). For MERS, the median number of cases averted increased from 2 (95% PrI: 0-60) for adm1 hospital catchment areas, to 6 (95% PrI: 0-83) for adm2 catchment areas, and 77 (95% PrI: 0-342) for adm1 catchment areas. For Lassa fever, the median number of cases averted increased from 0 (95% PrI: 0-306) for adm1 hospital catchment areas and 0 (95% PrI: 0-357) for adm2 catchment areas, to 101 (95% PrI: 3-771) for adm1 catchment areas. For RVF, the median number of cases averted increased from 29 (95% PrI: 0-3,525) for adm1 hospital catchment areas, to 43 (95% PrI: 0-5,826) for adm2 catchment areas, and 66 (95% PrI: 0-2,451) for adm1 catchment areas. For Nipah, the median number of cases averted was 0 for each catchment area definition, but the mean was highest for adm1 catchment areas. The total number of cases averted via vaccination of HCWs was also lowest using adm1 hospital catchment areas and highest using adm1 catchment areas (Figures 1D, S33D-S34D). For example, for MERS, the median number of nosocomial cases averted increased from 1 (95% PrI: 0-60) for adm1 hospital catchment areas, to 4 (95% PrI: 0-77) for adm2 catchment areas, and 55 (95% PrI: 0-259) for adm1 catchment areas.

Although the number of cases averted via reactive vaccination was highest using adm1 catchment areas, the number of cases averted per vaccine regimen administered was not necessarily the highest under this scenario because the number of regimens required was also higher using adm1 catchment areas. For MERS, the highest per regimen impact was achieved using adm1 hospital catchment areas where a median of 0.75 (95% PrI: 0-18.10) cases were averted per 100,000 vaccine regimens administered. In comparison, a median of 0.58 (95% PrI: 0.02-2.58) cases were averted per 100,000 vaccine regimens administered in adm1 hospital catchment areas, and 0.49 (95% PrI: 0-5.21) cases were averted per 100,000 vaccine regimens administered in adm2 catchment areas. The highest per regimen impact for RVF was also achieved using adm1 hospital catchment areas, with a median of 3.18 cases averted per 100,000 vaccine regimens administered versus 2.86 (95% PrI: 0-349.78) using adm2 catchment areas or 1.69 (95% PrI: 0-68.42) using adm1 catchment areas. For Lassa fever the per regimen impact was relatively consistent across different catchment areas, and for Nipah the median impact per 100,000 vaccine regimens administered was 0 for adm2 or adm1 hospital catchment areas and 0.01 (95% PrI: 0-11.17) for adm1 catchment areas. For MERS and Lassa fever, the largest impact of vaccinating HCWs as measured on a per-regimen-administered basis, was also achieved using adm1 hospital catchment areas. The per-regimen impact of vaccinating HCWs was minimal for Nipah, just as it was for vaccinating the general population (although the estimated per-regimen impact of vaccinating HCWs was higher than the impact of vaccinating the general population).

---

**Figure S23.**
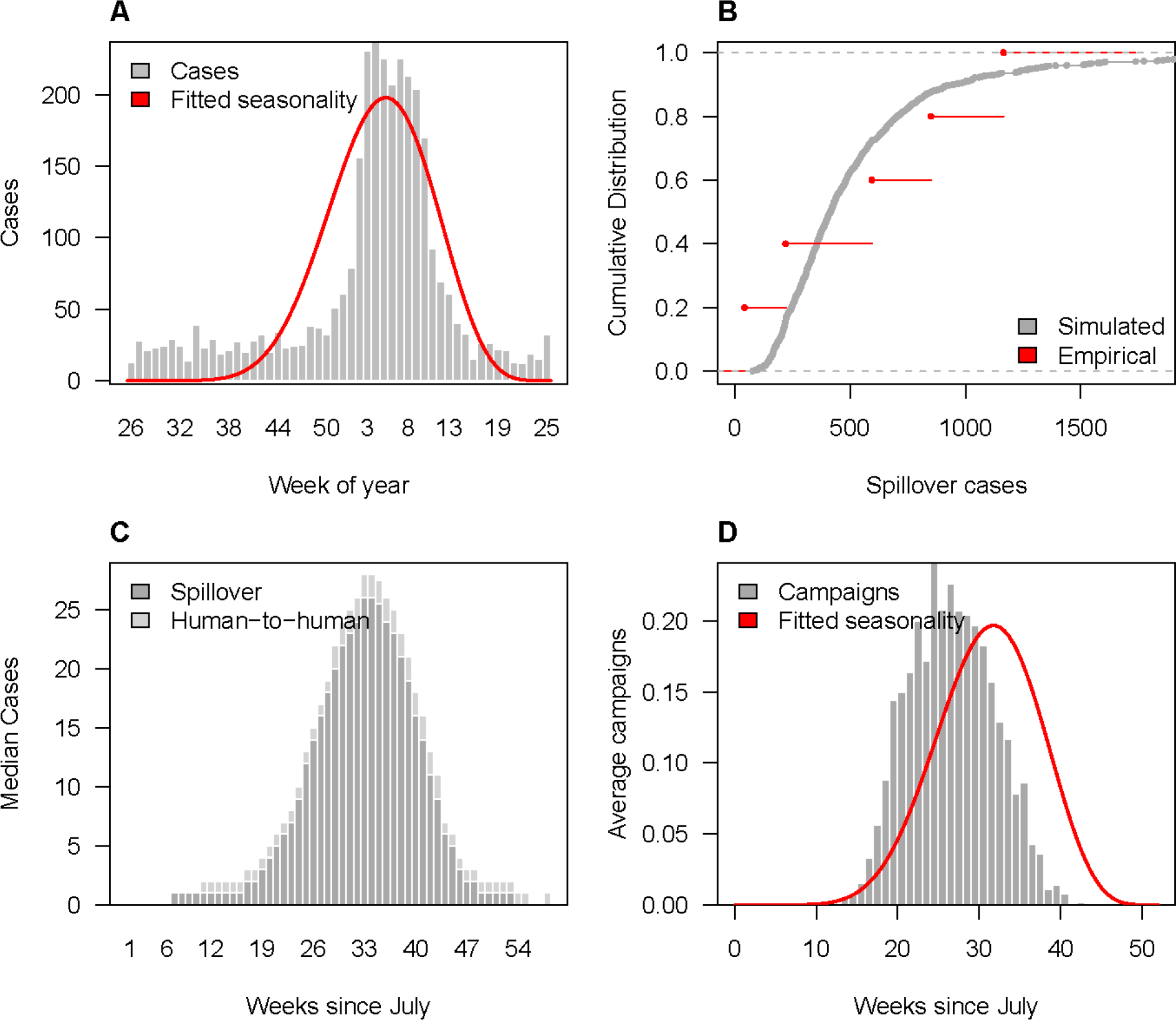
Spillover and reactive vaccination patterns for Lassa fever virus (LASV) within adm1 catchment areas. (A) Observed weekly Lassa fever spillover cases (grey bars) and estimated seasonal spillover rate (red line). (B) Annual number of spillovers over the past 5 years (red) and cumulative distribution of simulated annual spillovers from 1000 replicates (grey). (C) Median weekly simulated spillover and human-to-human Lassa fever cases. (D) Average weekly number of reactive campaigns triggered via spillover detection compared to the estimated seasonal spillover rate (red line).

**Figure S24.**
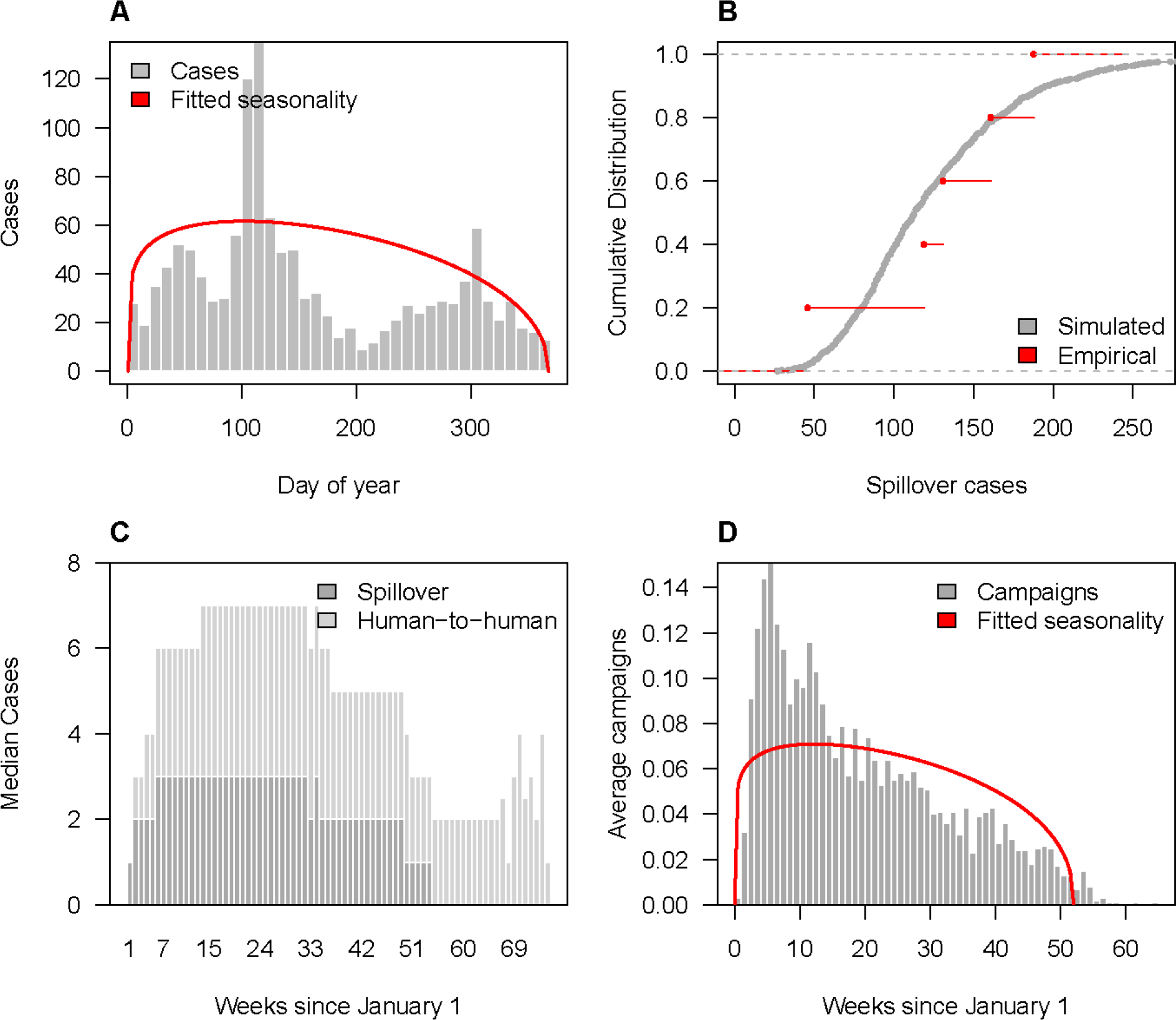
Spillover and reactive vaccination patterns for Middle Eastern respiratory virus (MERS-CoV) within adm1 catchment areas. (A) Observed weekly MERS spillover cases (grey bars) and estimated seasonal spillover rate (red line). (B) Annual number of spillovers over the past 5 years (red) and cumulative distribution of simulated annual spillovers from 1000 replicates (grey). (C) Median weekly simulated spillover and human-to-human MERS cases. (D) Average weekly number of reactive campaigns triggered via spillover detection compared to the estimated seasonal spillover rate (red line).

**Figure S25.**
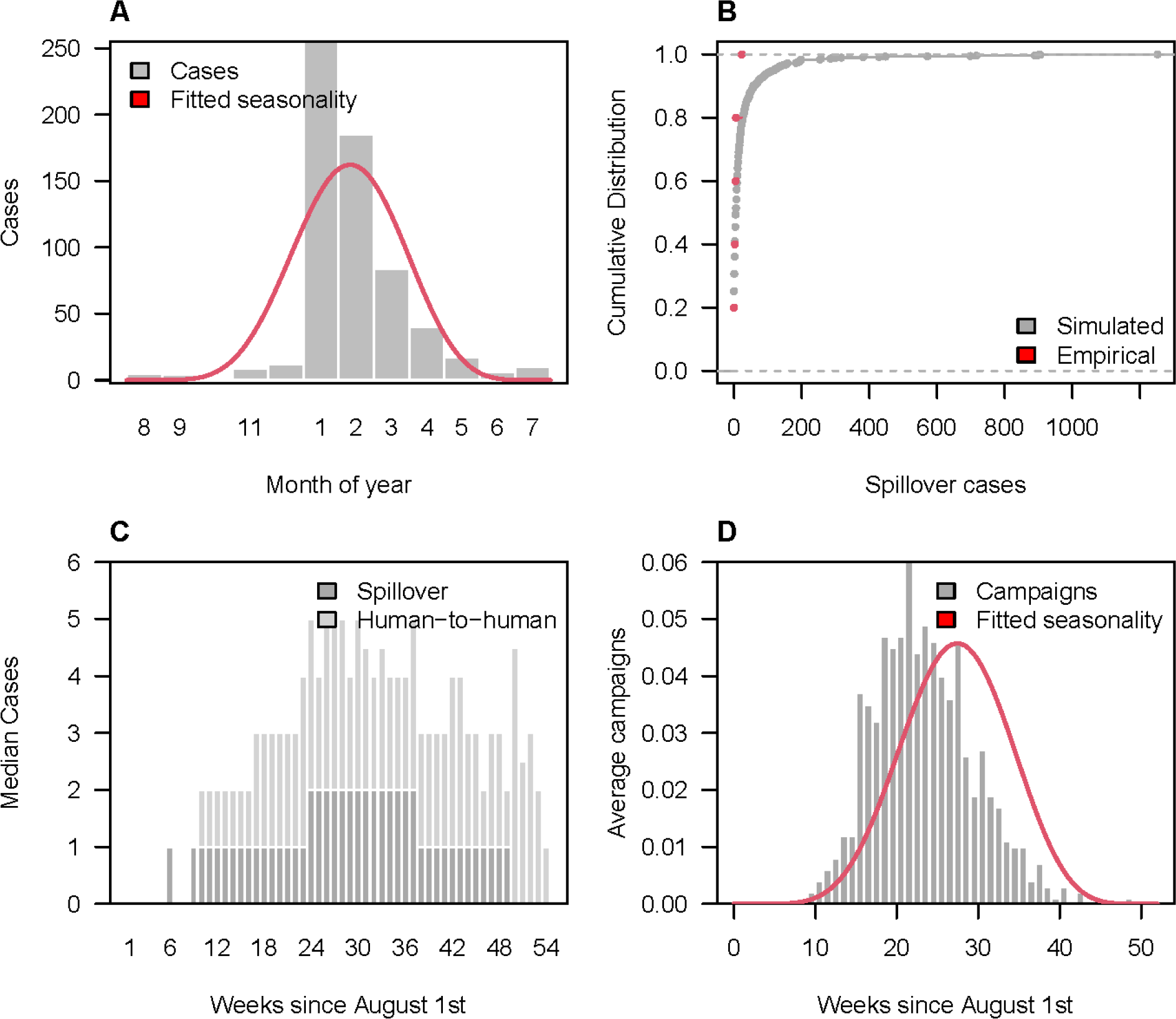
Spillover and reactive vaccination patterns for Nipah virus (NiV) within adm1 catchment areas. (A) Observed weekly Nipah spillover cases (grey bars) and estimated seasonal spillover rate (red line). (B) Annual number of spillovers over the past 5 years (red) and cumulative distribution of simulated annual spillovers from 1000 replicates (grey). (C) Median weekly simulated spillover and human-to-human Nipah cases. (D) Average weekly number of reactive campaigns triggered via spillover detection compared to the estimated seasonal spillover rate (red line).

**Figure S26.**
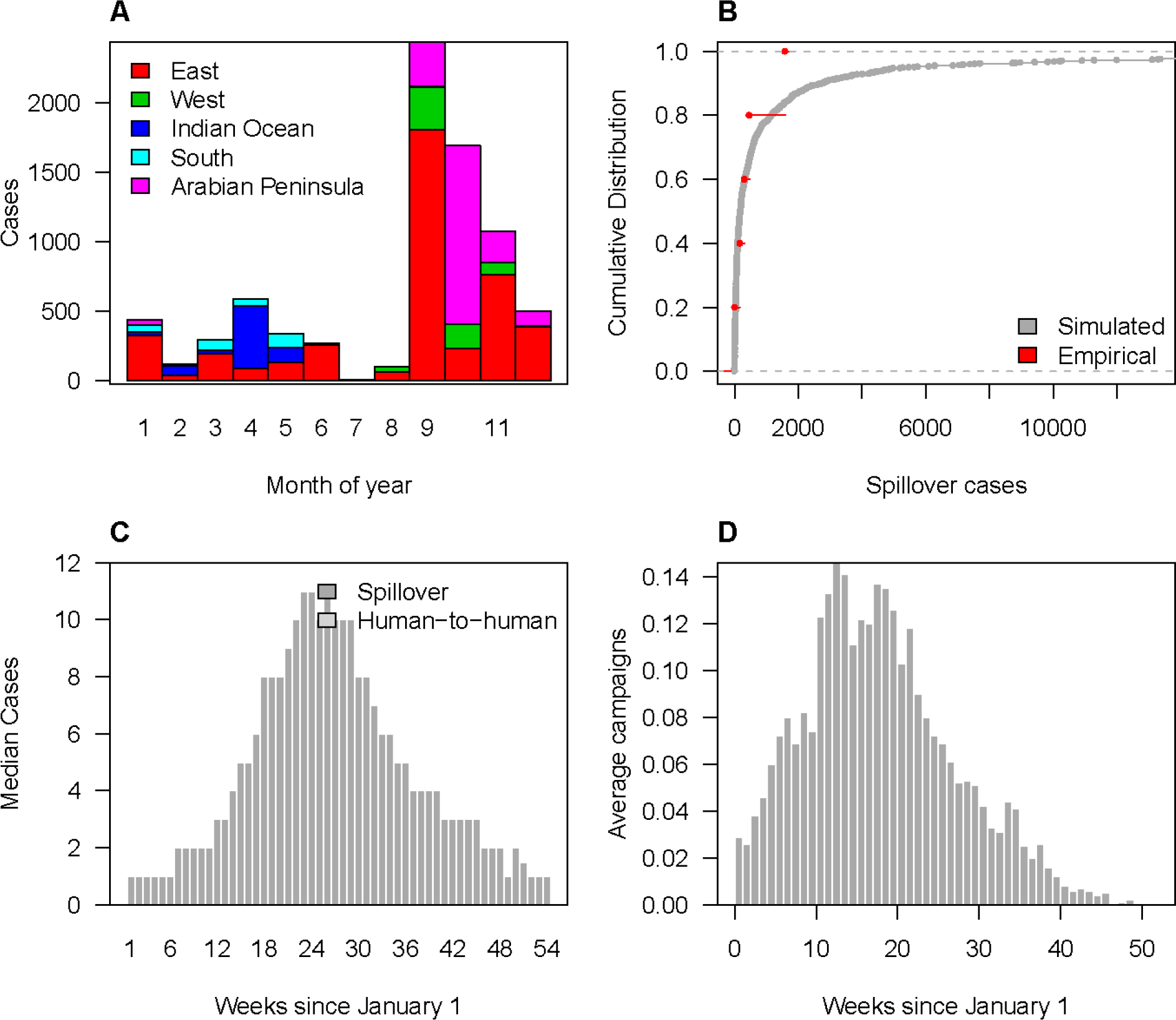
Spillover and reactive vaccination patterns for Rift Valley fever virus (RVFV) within adm1 catchment areas. (A) Observed monthly RVF spillover cases by region. (B) Annual number of spillovers over the past 5 years (red) and cumulative distribution of simulated annual spillovers from 1000 replicates (grey). (C) Median weekly simulated spillover and human-to-human RVF cases. (D) Average weekly number of reactive campaigns triggered via spillover detection compared to the estimated seasonal spillover rate (red line).

**Figure S27.**
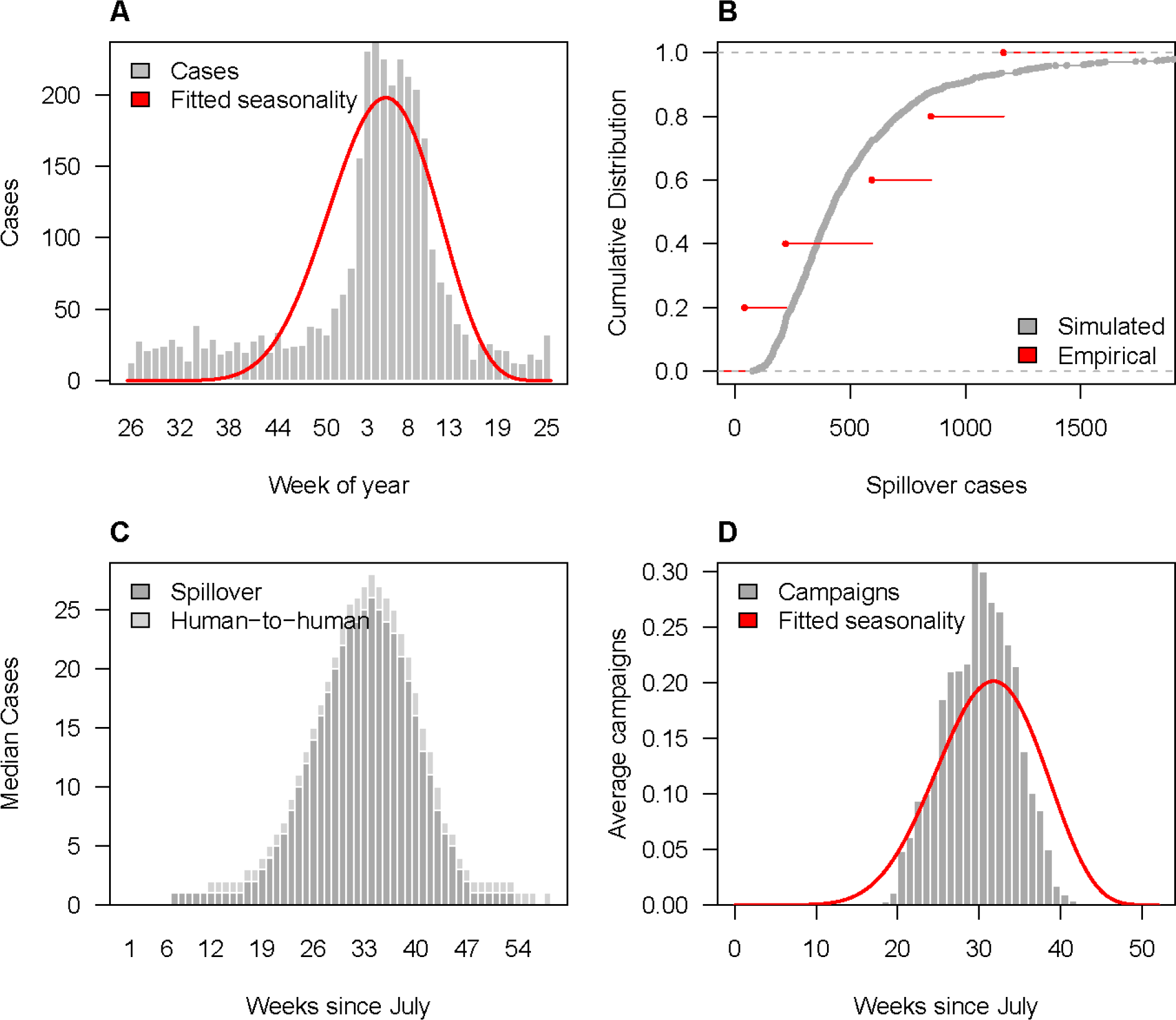
Spillover and reactive vaccination patterns for Lassa fever virus (LASV) within adm1 hospital catchment areas. (A) Observed weekly Lassa fever spillover cases (grey bars) and estimated seasonal spillover rate (red line). (B) Annual number of spillovers over the past 5 years (red) and cumulative distribution of simulated annual spillovers from 1000 replicates (grey). (C) Median weekly simulated spillover and human-to-human Lassa fever cases. (D) Average weekly number of reactive campaigns triggered via spillover detection compared to the estimated seasonal spillover rate (red line).

**Figure S28.**
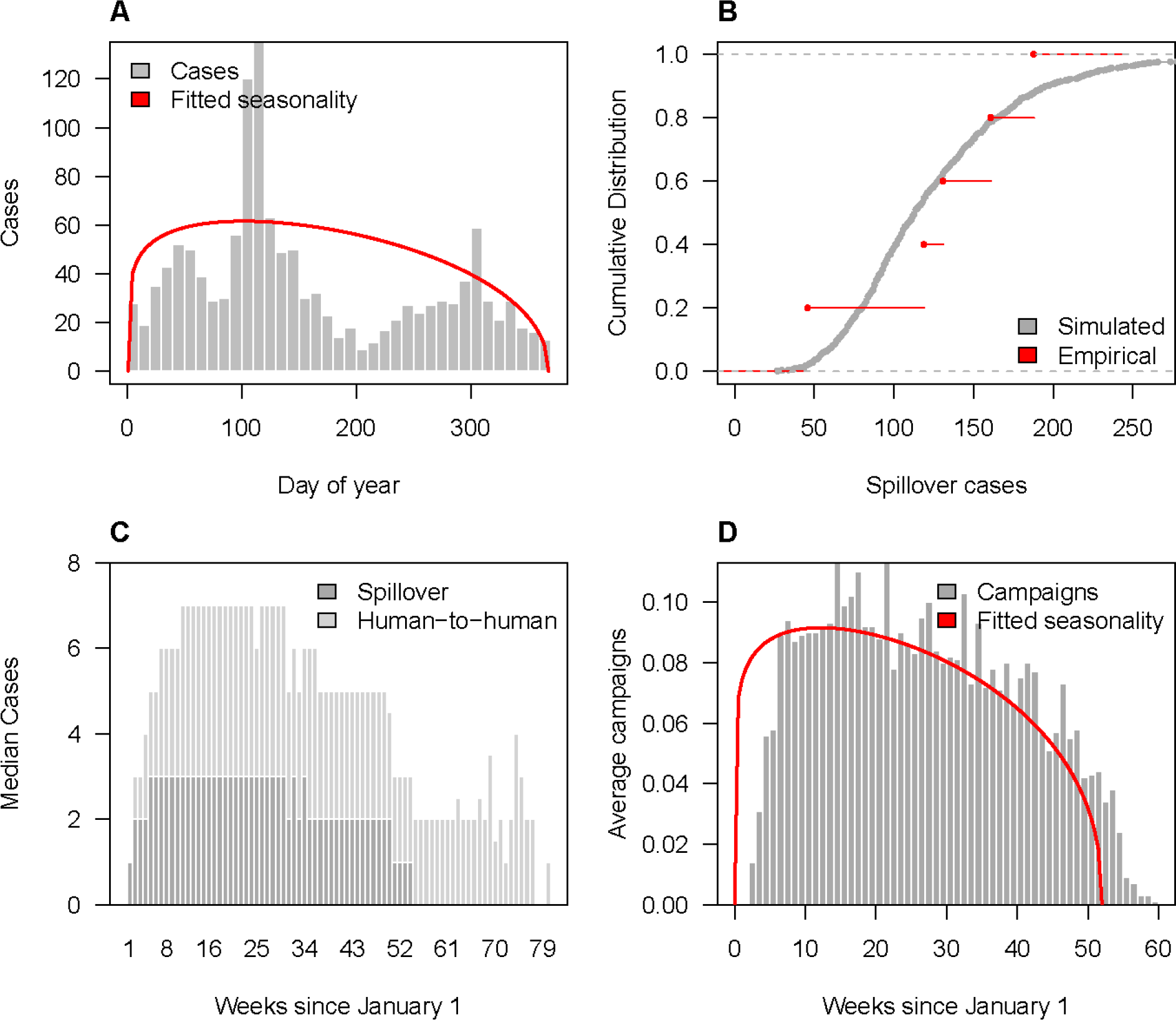
Spillover and reactive vaccination patterns for Middle Eastern respiratory virus (MERS-CoV) within adm1 hospital catchment areas. (A) Observed weekly MERS spillover cases (grey bars) and estimated seasonal spillover rate (red line). (B) Annual number of spillovers over the past 5 years (red) and cumulative distribution of simulated annual spillovers from 1000 replicates (grey). (C) Median weekly simulated spillover and human-to-human MERS cases. (D) Average weekly number of reactive campaigns triggered via spillover detection compared to the estimated seasonal spillover rate (red line).

**Figure S29.**
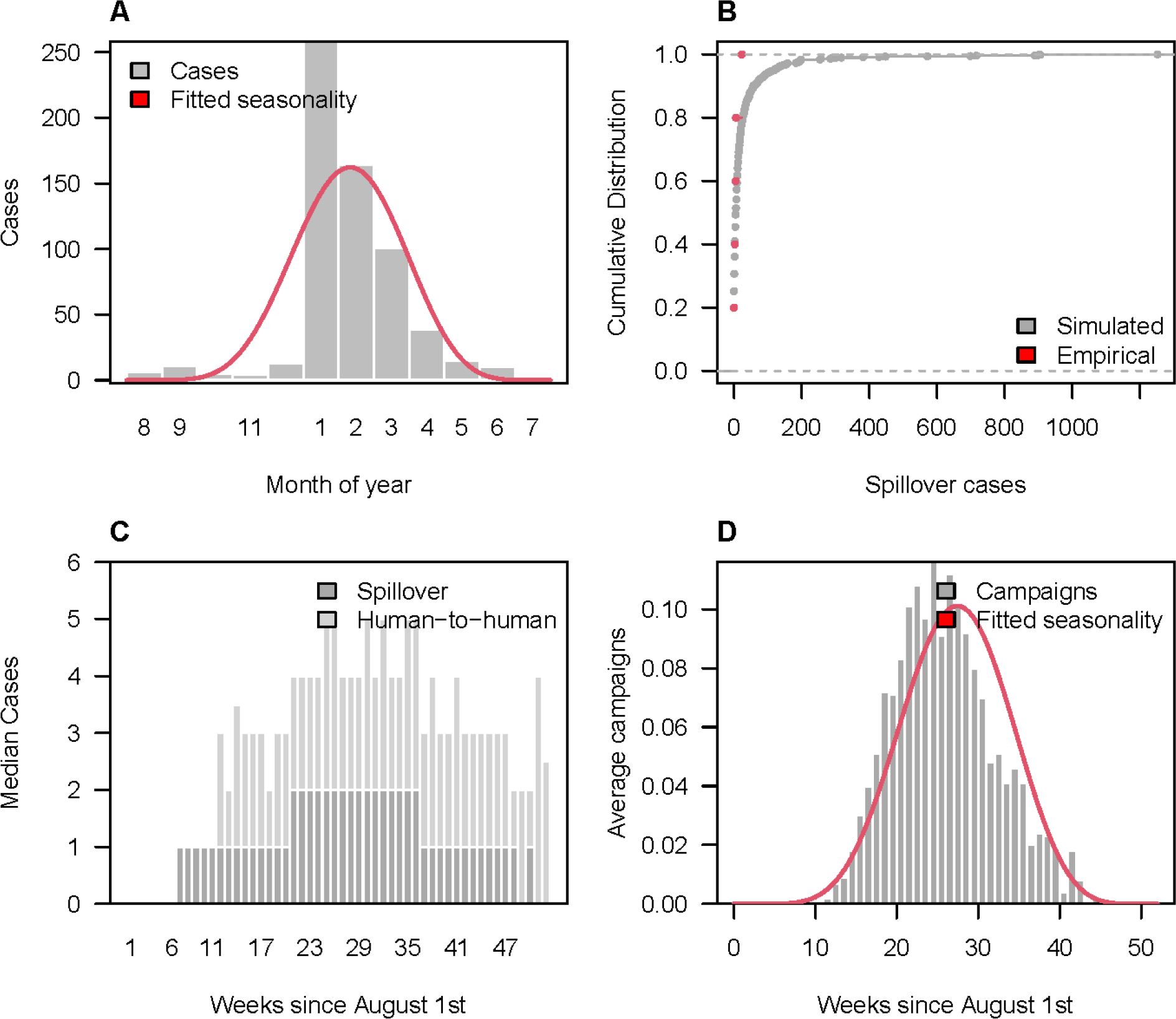
Spillover and reactive vaccination patterns for Nipah virus (NiV) within adm1 hospital catchment areas. (A) Observed weekly Nipah spillover cases (grey bars) and estimated seasonal spillover rate (red line). (B) Annual number of spillovers over the past 5 years (red) and cumulative distribution of simulated annual spillovers from 1000 replicates (grey). (C) Median weekly simulated spillover and human-to-human Nipah cases. (D) Average weekly number of reactive campaigns triggered via spillover detection compared to the estimated seasonal spillover rate (red line).

**Figure S30.**
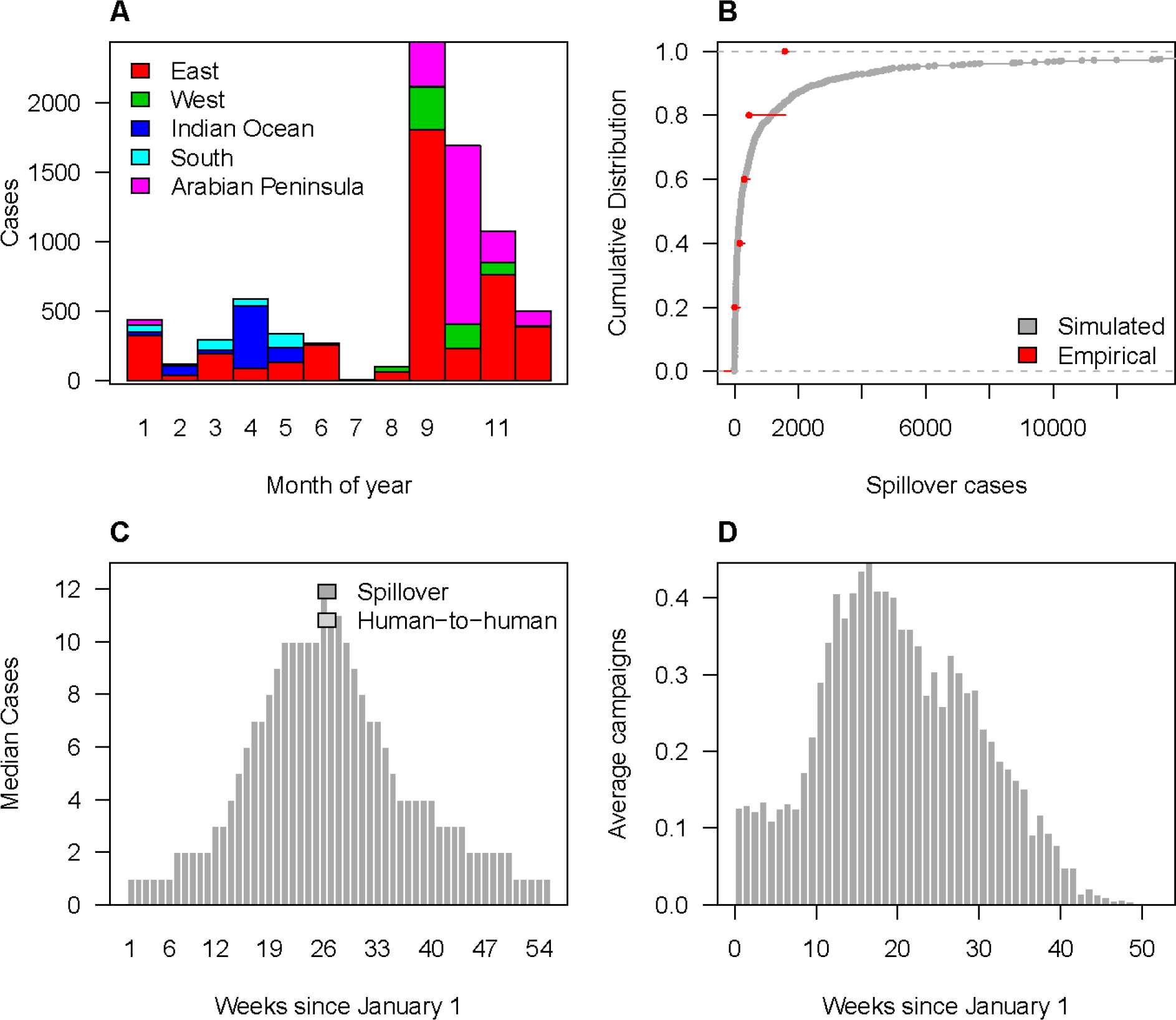
Spillover and reactive vaccination patterns for Rift Valley fever virus (RVFV) within adm1 hospital catchment areas. (A) Observed monthly RVF spillover cases by region. (B) Annual number of spillovers over the past 5 years (red) and cumulative distribution of simulated annual spillovers from 1000 replicates (grey). (C) Median weekly simulated spillover and human-to-human RVF cases. (D) Average weekly number of reactive campaigns triggered via spillover detection compared to the estimated seasonal spillover rate (red line).

**Figure S31.**
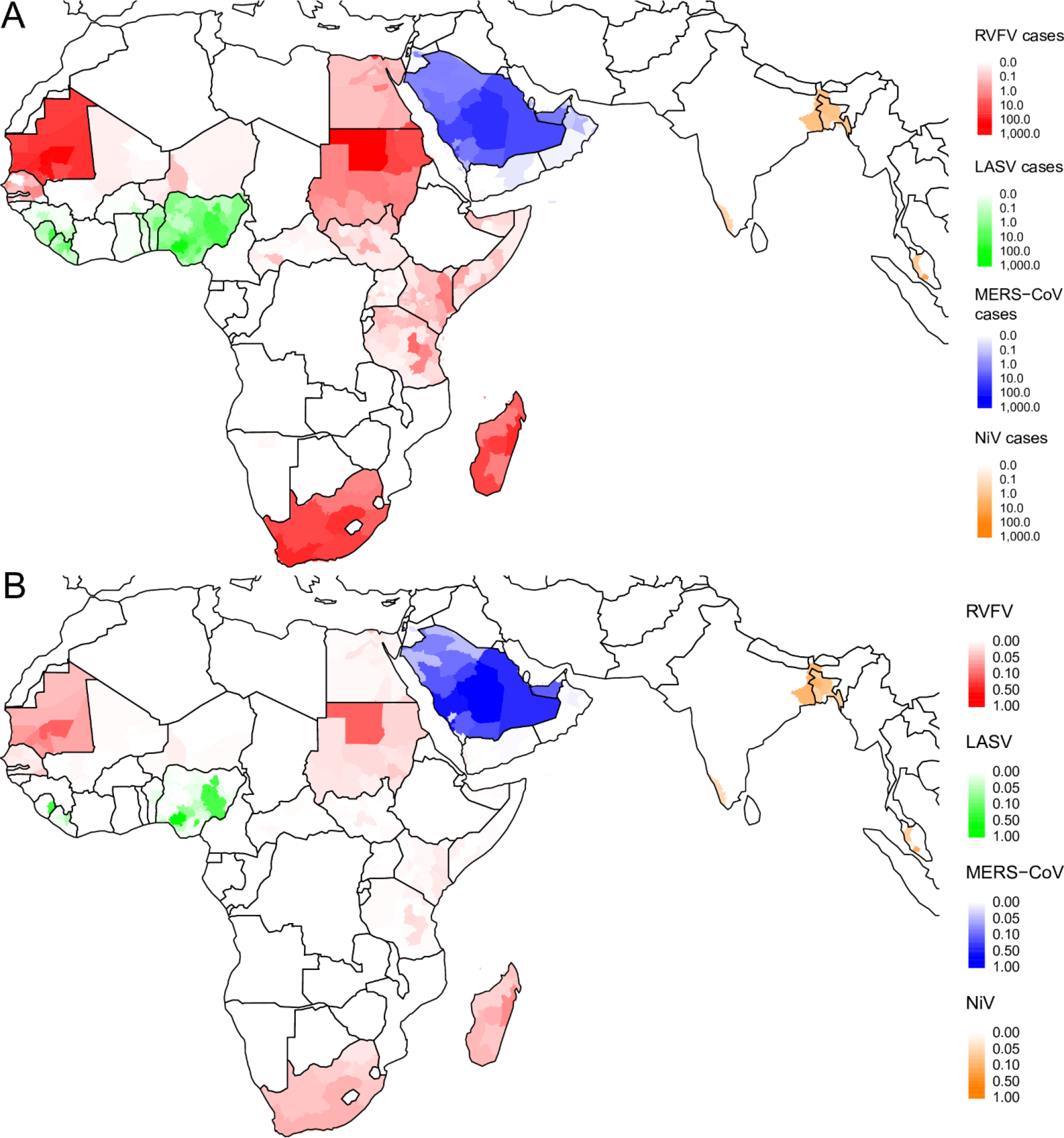
Geographic distribution of spillover cases and reactive vaccination campaigns for adm1 catchment areas. (A) Geographic distribution of the expected annual number of spillover cases for each pathogen. (B) Probability that a campaign will be triggered.

**Figure S32.**
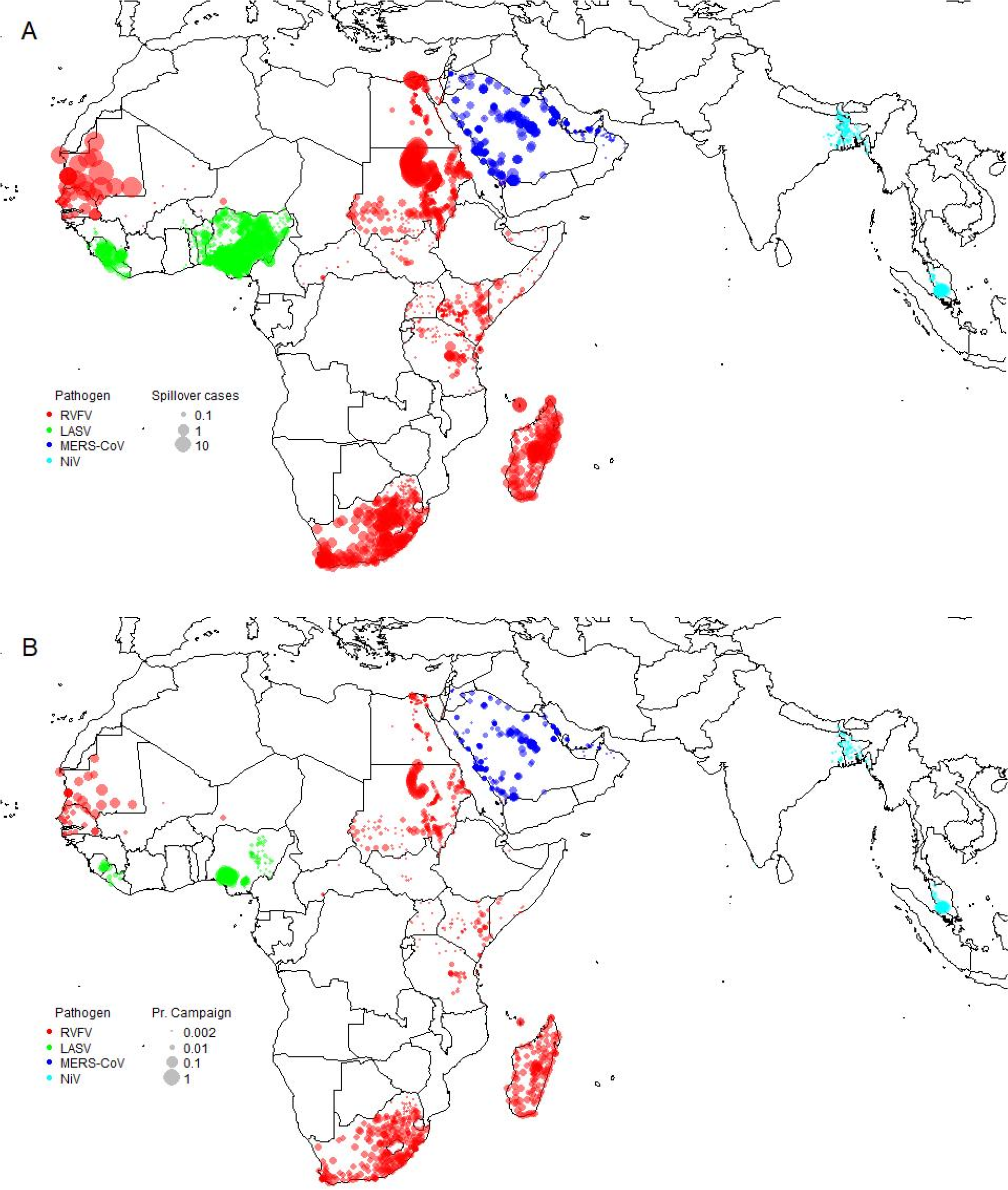
Geographic distribution of spillover cases and reactive vaccination campaigns for adm1 hospital catchment areas. (A) Geographic distribution of the expected annual number of spillover cases for each pathogen. (B) Proportion of time a campaign will be triggered.

**Figure S33.**
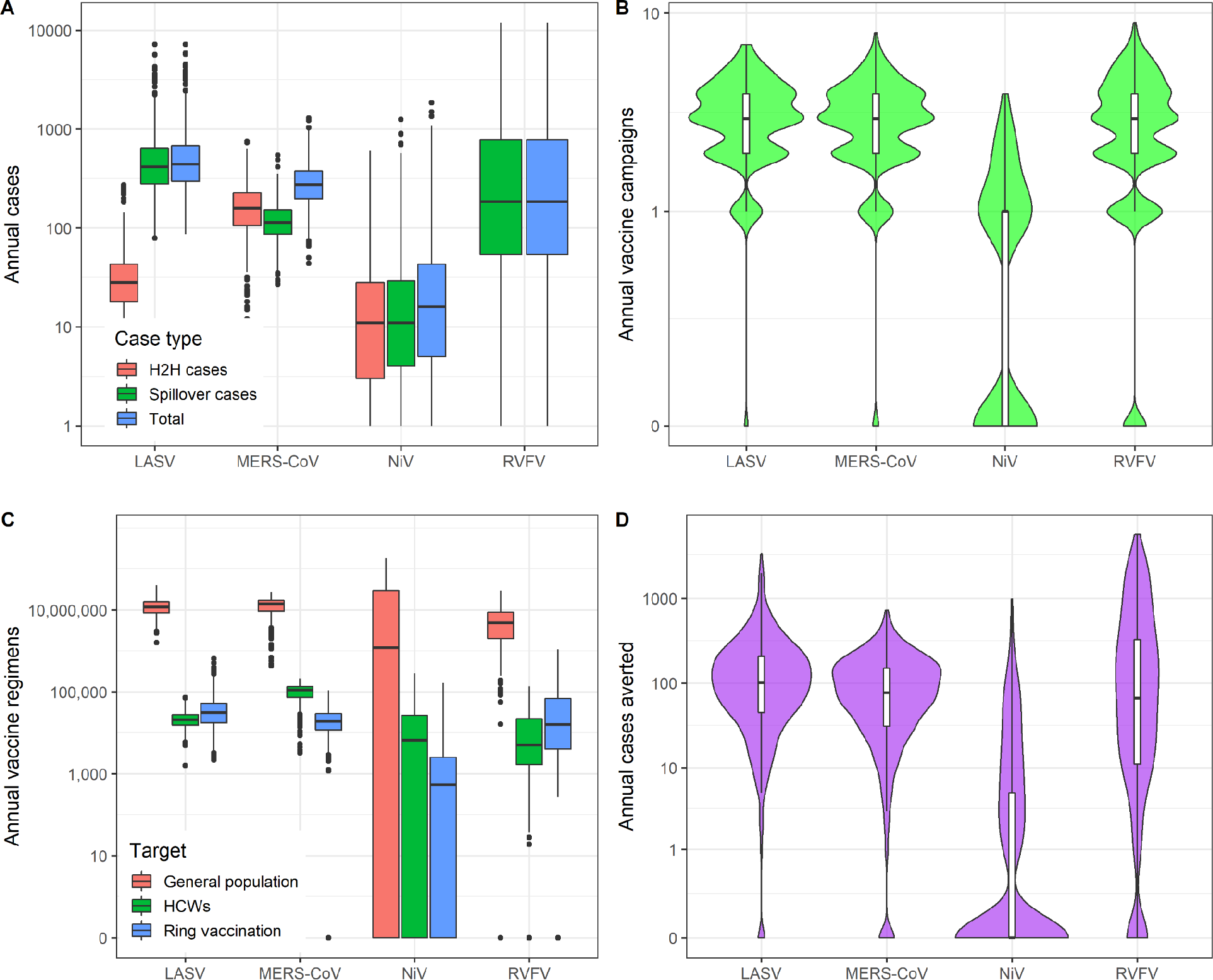
Annual cases and reactive vaccination impacts for adm1 catchment areas. (A) Annual number of spillover, human-to-human (H2H), and total cases for each pathogen across the entire study region. (B) Annual number of vaccine campaigns that will be triggered due to the outbreak threshold. (C) Number of vaccine regimens required per year for outbreak response when either the general population or healthcare workers (HCWs) only are targeted. (D) Annual number of cases averted via vaccination.

**Figure S34.**
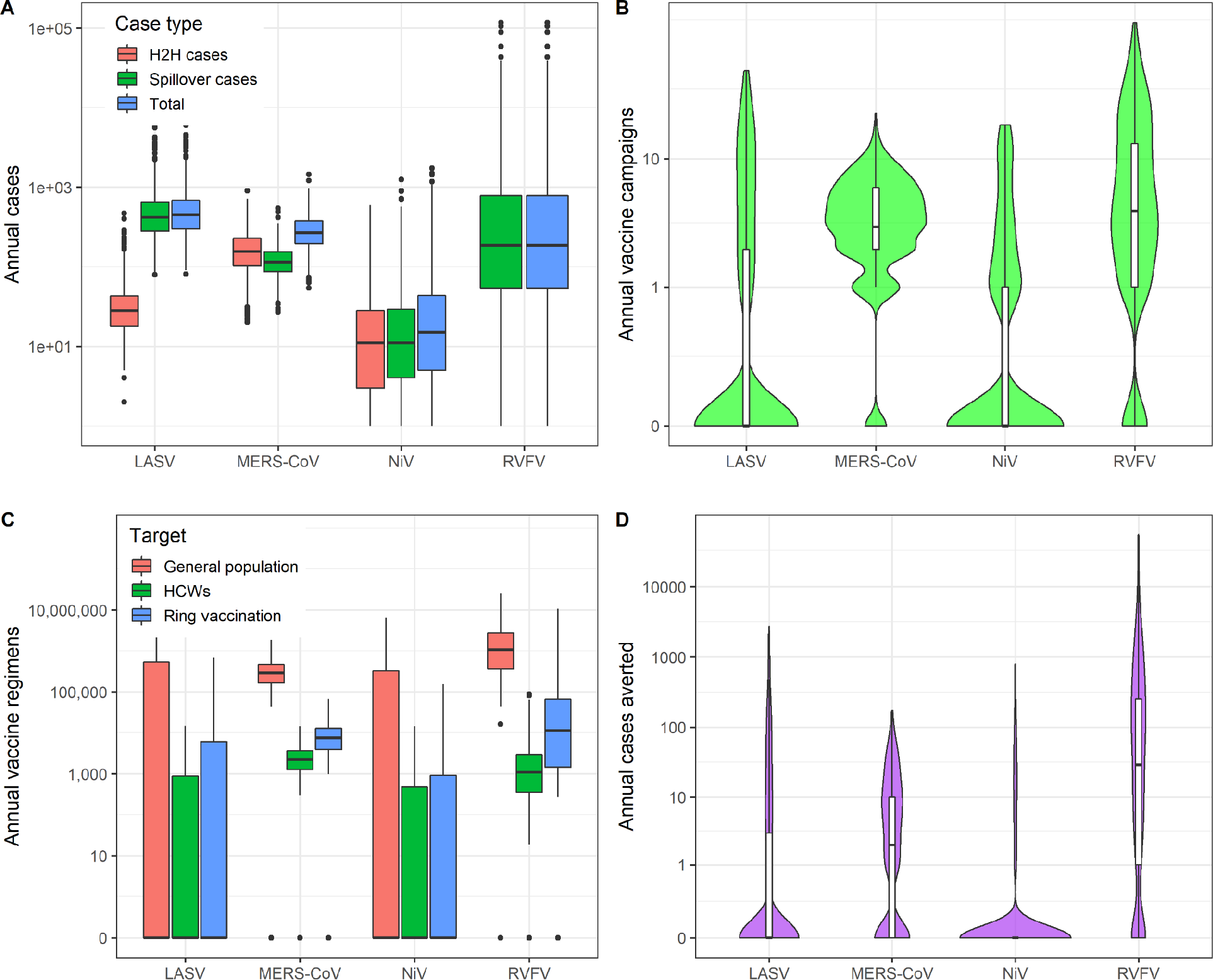
Annual cases and reactive vaccination impacts for adm1 hospital-based catchment areas. (A) Annual number of spillover, human-to-human (H2H), and total cases for each pathogen across the entire study region. (B) Annual number of vaccine campaigns that will be triggered due to the outbreak threshold. (C) Number of vaccine regimens required per year for outbreak response when either the general population or healthcare workers (HCWs) only are targeted. (D) Annual number of cases averted via vaccination.

